# Pervasive Influence of Hormonal Contraceptives on the Human Plasma Proteome in a Broad Population Study

**DOI:** 10.1101/2023.10.11.23296871

**Authors:** Nikola Dordevic, Clemens Dierks, Essi Hantikainen, Vadim Farztdinov, Fatma Amari, Vinicius Verri Hernandes, Alessandro De Grandi, Francisco S. Domingues, Michael Mülleder, Peter Paul Pramstaller, Johannes Rainer, Markus Ralser

## Abstract

**Background:** Plasma proteomics offers new avenues to explore non-genetic associations, such as biomarkers for lifestyle and environmental exposure in population studies. To date, most proteomic investigations in population studies have utilized affinity-reagent based technologies, which are ideal to quantify the low abundant fraction of the circulating proteome but may omit several of the abundant proteins that function in plasma.

**Methods:** Utilizing high throughput mass spectrometry, we quantified 148 highly abundant protein groups including immunoglobulins, coagulation factors, metabolic proteins, and components of the innate immune system, in the plasma of 3,632 participants from the Cooperative Health Research in South Tyrol (CHRIS) study. Using multiple regression analyses we then investigated associations with various factors including common medications.

**Results:** Beyond age and sex, the high abundant plasma proteome is predominantly influenced by hormonal contraceptives. For instance, Angiotensinogen (AGT) levels exhibit significant alteration with this treatment, suggesting that AGT levels could be a potential biomarker for contraceptive use. The effect of this drug class is more pronounced than other common medications or covariates. Furthermore, our analysis does not reveal any enduring signature associated with the use of these contraceptives.

**Conclusion:** In contrast to most used drugs, hormonal contraceptives exert a pronounced effect on the high abundant plasma proteome. Given its high prevalence among young female participants, the impact of hormonal contraceptives might be misconstrued as sex-or age-related effects on the plasma proteome. One should thus account for their use in any epidemiological or clinical plasma proteome study to prevent misleading results.

## Introduction

Human blood represents an easily obtainable and sensitive matrix for the assessment of health and disease in individuals or populations. The abundance of proteins, their respective isoforms, potential post-translational modifications and protein sequence variants provide a snapshot of the current physiological state of the circulatory system and all organs with which blood comes into contact (Anderson and Anderson, 2002; Deutsch et al., 2021; Ignjatovic et al., 2019; Vernardis et al., 2023). In population studies, large-scale plasma proteomics provides new opportunities to study non-genetic associations to health-related traits, such as markers for lifestyle and environmental exposure or to detect and characterize onset and progression of disease through longitudinal monitoring of protein abundance changes (Ferkingstad et al., 2021; Palstrøm et al., 2022; Suhre et al., 2021; Sun et al., 2023). While statistical approaches ranging from multivariate linear statistics to traditional machine learning have proven successful in identifying biomarkers and utilizing them for disease prediction (Bader et al., 2020; Demichev et al., 2021), large study cohorts would enable the application of more powerful deep learning models (Bader et al., 2023). The quantification of the plasma proteome is a formidable challenge due to a combination of factors: the exceptionally high abundance of select plasma proteins, the wide dynamic range of protein concentrations, the substantial sequence variability of certain proteins, and the fluctuations in protein abundances in response to diseases, physiological changes, or lifestyle factors. This challenge is particularly pronounced in large-scale studies, where it becomes even more daunting due to the heightened technical complexities involved (Deutsch et al., 2021; Suhre et al., 2021).

Recently, different technologies emerged to measure proteins in human blood plasma or serum samples. These range from optimized single-protein assays to more flexible mass-spectrometry (MS) based workflows to affinity-based multiplex assays. For instance, highly multiplexed affinity-based platforms, such as Luminex, Olink and SomaScan, have emerged that offer attractive and fast high throughput assays for measurement of plasma protein panels in thousands of samples (Ferkingstad et al., 2021; Smith and Gerszten, 2017; Suhre et al., 2021). While throughput and sensitivity are high for these methods, their susceptibility to off-target binding as well as the lack of reproducibility raised questions about the quality of some affinity-based measures (Baker, 2015). Moreover, due to saturation of the affinity reagents, and the difficulty to differentiate among isoforms, many affinity-reagent based methods omit many highly abundant plasma proteins. Specifically, the high abundant plasma protein fraction is however enriched for proteins that exert their function in the plasma, which is responsible for nutrient transport, responds to lifestyle intervention and immune system activity. Indeed, the high abundant plasma proteome fraction is attractive for the development of marker panel assays for medical research and clinical use and encompasses most of the protein biomarkers used to date (Anderson and Anderson, 2002; Hartl et al., 2023; Macklin et al., 2020; Vernardis et al., 2023).

Mass spectrometry has a high specificity in protein identification and is not limited to the detection of a predefined group of proteins. Furthermore, it allows the detection of post-translational modifications or even interactions (Deutsch et al., 2021). MS-based proteomics increasingly allows the absolute quantification of plasma proteins and provides a simple path for the development of biomarker assays (Hartl et al., 2023; Macklin et al., 2020). While mass spectrometry per se is highly sensitive and can detect individual peptides down to zeptomoles, it struggles to detect low abundant plasma proteins due to the high dynamic range in the plasma proteome. MS methods can be made applicable for the analysis of the low abundant plasma proteome fraction, in particular, upon depletion of the high abundant fraction of the plasma proteome (Reymond et al., 2023; Tu et al., 2010). However, protein depletion adds variability and cost, and similar to the use of affinity reagents, compromises the quantification of the important high-abundant plasma fraction. With the goal to complement the existing proteomic studies with quantities about the high abundant plasma protein fraction, we recently presented a platform technology that combines a semi-automated sample preparation workflow, analytical flow rate chromatography for gradient lengths of 0.5-5 minutes, specifically optimized DIA-MS acquisition schemes, and an adapted data processing software suite which integrates artificial neural networks in raw data processing (DIA-NN) (Demichev et al., 2020; Messner et al., 2020; Szyrwiel et al., 2023). This platform achieves high measurement precision, and due to the high sample throughput as provided by the analytical flow rate chromatography, is highly cost effective in the processing of large plasma proteomes.

The Cooperative Health Research in South Tyrol (CHRIS) study (Pattaro et al., 2015) is a single-site population-based study aimed to investigate the genetic and molecular basis of common age-related chronic conditions and their interaction with lifestyle and the environment. In recent work, we have been evaluating the impact of age, sex and diet, amongst others, on the human metabolome (Verri Hernandes et al., 2022). Moreover, gene-metabolite associations (König et al., 2022), as well as genetic and metabolomic determinants of disease (Emmert et al., 2021), have been investigated. Using Scanning SWATH (Messner et al., 2021) acquisition on Triple TOF 6600 Instruments (Sciex) we created a MS-based plasma proteomics data set with low technical variability for n = 3,632 CHRIS participants. We performed a general exploratory analysis of the data set to identify relevant factors affecting the plasma proteome, including commonly used drugs. We found hormonal contraceptives to be the main factor explaining the variation in a human plasma proteome in this European cohort.

## Results

### Study Sample Characteristics and General Data Overview

To quantify high abundant plasma proteins in 3,632 participants of the CHRIS study (Pattaro et al., 2015) (demographics shown in Table 1), citrate plasma samples were randomly arrayed on 50 96-well plates assigning nuclear families to the same plate. Each plate included 79 study samples, 4 replicates of the study pool, 1 procedural blank, 4 commercial serum and 8 commercial EDTA plasma samples. These serve as quality control (QC) and reference samples to compare, cross-reference and join large studies (Dammer et al., 2023; Pino et al., 2018). Measurements were performed on tryptic digests that have been created from the plasma citrate samples. These were analyzed utilizing high-flowrate liquid chromatography data independent acquisition mass spectrometry (LC-DIA-MS). Specifically, we used Scanning SWATH (Messner et al., 2021) acquisition, using 800µL/min, 5-minute water to acetonitrile chromatographic gradients, as described in the Materials and Methods section (Messner et al., 2021).

**Table 1:**
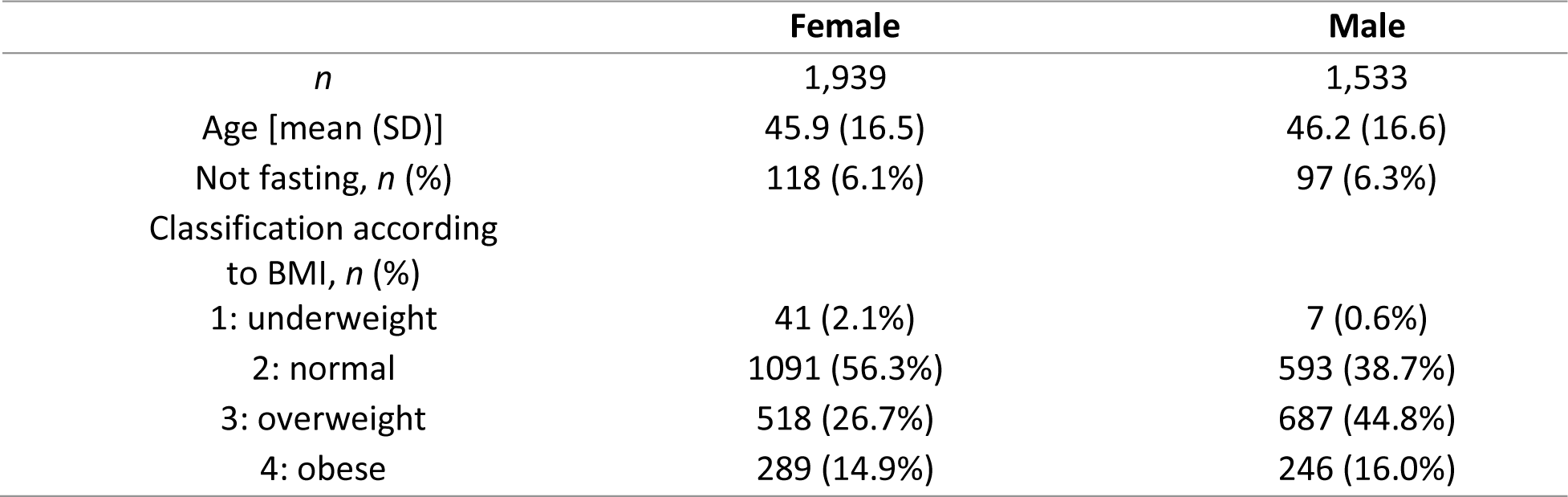
Demographic characteristics of the study participants included in the analysis.

|  | **Female** | **Male** |
| --- | --- | --- |
| *n* | 1,939 | 1,533 |
| Age [mean (SD)] | 45.9 (16.5) | 46.2 (16.6) |
| Not fasting, *n* (%) | 118 (6.1%) | 97 (6.3%) |
| Classification according to BMI, *n* (%) |  |  |
| 1: underweight | 41 (2.1%) | 7 (0.6%) |
| 2: normal | 1091 (56.3%) | 593 (38.7%) |
| 3: overweight | 518 (26.7%) | 687 (44.8%) |
| 4: obese | 289 (14.9%) | 246 (16.0%) |

The full set of 5,125 unique samples, included thus 977 quality control (QC) samples, 200 pools of study samples and 350 samples from a sub-study of CHRIS (Motta et al., 2019). This sample set was measured in 17 batches over the time span of 6 months on two Triple TOF 6600 instruments (Sciex) that were connected to 1290 Infinity II chromatography systems (Agilent technologies). Data was recorded using Analyst (Sciex) and processed in DIA-NN using a spectral library approach (Methods).

The data matrix consisted, after removal of 55 outliers and all QC samples of abundances for 6,762 peptide precursors in 4,093 samples. Peptide features with more than 40% missing values across all study samples were excluded, reducing thus the data set to a final number of 2,716 precursors. Cyclic loess normalization and plate correction were then applied to remove batch effects between plates as well as any other measurement-specific technical variation. Final summarization of peptides to protein abundances was performed after proteotypic filtering of the peptides to a final number of 2,386. In this final dataset, the quantitative precision is estimated with a median coefficient of variation (CV) to 15.45% and 30.97% for pooled and study samples (see Supplementary Figure S149 for distribution of CV before and after normalization). The set of quantified proteins along with the results from the present analysis are available in Supplementary Table S1. The final data set used for this analysis consisted, after removal of samples from pregnant women and participants with missing information for any of the traits listed in Table 1, of 148 proteins in 3,472 study samples.

While for most of the proteins the signal distribution across samples was about log-normal, some proteins, including AGT, SERPING1, IGHV5-10-1 and IGHV6-1, showed a clear bimodal signal distribution in the present data set (see Supplementary Figures S1-S148 for signal distributions of all proteins).

### Protein Coverage and Variation in the CHRIS Cohort

Of the consistently measured high abundant proteins, 139 were enriched in the following GO:BP pathways: complement cascade pathways (complement activation; complement activation, classical pathway), immune response pathways (humoral immune response; immunoglobulin mediated immune response; B cell mediated immunity; adaptive immune response; innate immune response among others); phagocytosis, blood coagulation, hemostasis, endocytosis, response to bacterium and many others (see Supplementary Table S2). For a more detailed investigation of protein functionality, we in addition used the PANTHER classification system (Mi et al., 2013). Of the 148 plasma proteins consistently quantified, 134 could be assigned to 25 different functional classes. The most prominent were immunoglobulins (n = 27), protease inhibitors (22), serine proteases (13), components of the complement system (10) and apolipoproteins (9) (see Supplementary Table S2).

We next evaluated the variation of these proteins across our population study. The median coefficient of variation (CV) of all proteins across study samples was 31.1% with the 25% and 75% quantile being 21.5% and 47.3%, respectively (CV for all proteins included in Supplementary Table S1). Among the proteins with the lowest CV were, next to albumin (ALB; 13.3%), also proteins related to blood coagulation (F2, SERPINC1, KNG1 and SERPINF2; CV between 13% and 18.3%) as well as many proteins from the complement system (C3, C5, CFH, CFB, C1S, C8A, CFI and C1R, with a CV between 13.8% and 19.9%; see also Supplementary Table S3), reflecting that most of the individuals reported no acute condition at the time of sampling. To identify highly variable proteins, we calculated a relative CV defined as the ratio between the CVs in study and pooled QC samples. Among the top 30 proteins ranked by their relative CV were 12 immunoglobulins (IGHG4, IGHM, IGHV6-1, IGHV5-10-1, IGHA2, IGHG2, IGLV8-61, JCHAIN, IGHA1, IGHV2-26, IGKC and IGKV1-5, with a CV between 22% and 89.2%), 4 hemolysis-related proteins (HP, HPR, CP and HBB, with a CV between 26.9% and 54.5%) as well as the hormone transporters SERPINA6 (48.3%) and SHBG (123%; see Supplementary Table S4 for the full list). Of 46 common disease biomarker proteins detected previously using Scanning SWATH on neat plasma (Messner et al., 2020), 31 were also quantified in the present data set (Supplementary Table S5). Of these, 9, including ALB (13.3%), SERPINC1 (13.7%) and HPX (13.8%) had a CV in study samples below 20% while 10 had a CV larger than 40%, with the highest variable proteins being LPA (134%), SHBG (123%) and FN1 (CV = 88%).

Next, we compared quantified protein abundances to related available accredited laboratory measurements that either determine the enzymatic activity of the respective proteins or quantify protein complexes containing them (such as HDL and LDL). Correlation coefficients (Spearman’s rho) for the investigated pairs of measurements range from 0.32 for albumin to 0.79 for APOB and LDL (Supplementary Table S6 and Supplementary Figure S150). Hence, while generally significant correlations between protein abundance and the diagnostic assays was achieved, this comparison equally indicates that the protein quantification by mass spectrometry, and the indirect quantification of the parameters by enzyme assays or estimation of protein complexes levels, produces varying quantitative results.

### Oral Hormonal Contraceptives Shape the Plasma Proteome in Female Study Participants

To explore the data set and to investigate main influence factors on the present plasma proteome, we next performed a principal component analysis (PCA) on the z-score transformed abundances. This analysis revealed a subset of almost exclusively female individuals that separated from the main bulk of study participants on principal component 1 (PC1; see Figure 1, upper panel). This principal component, explaining the largest variance in the data set, also showed a clear relationship with the participants’ age (Figure 1, lower panel; the loading for age in the PCA was also approximately parallel to the direction of PC1) and, to a lower extent, with the participants’ sex.

**Figure 1:**
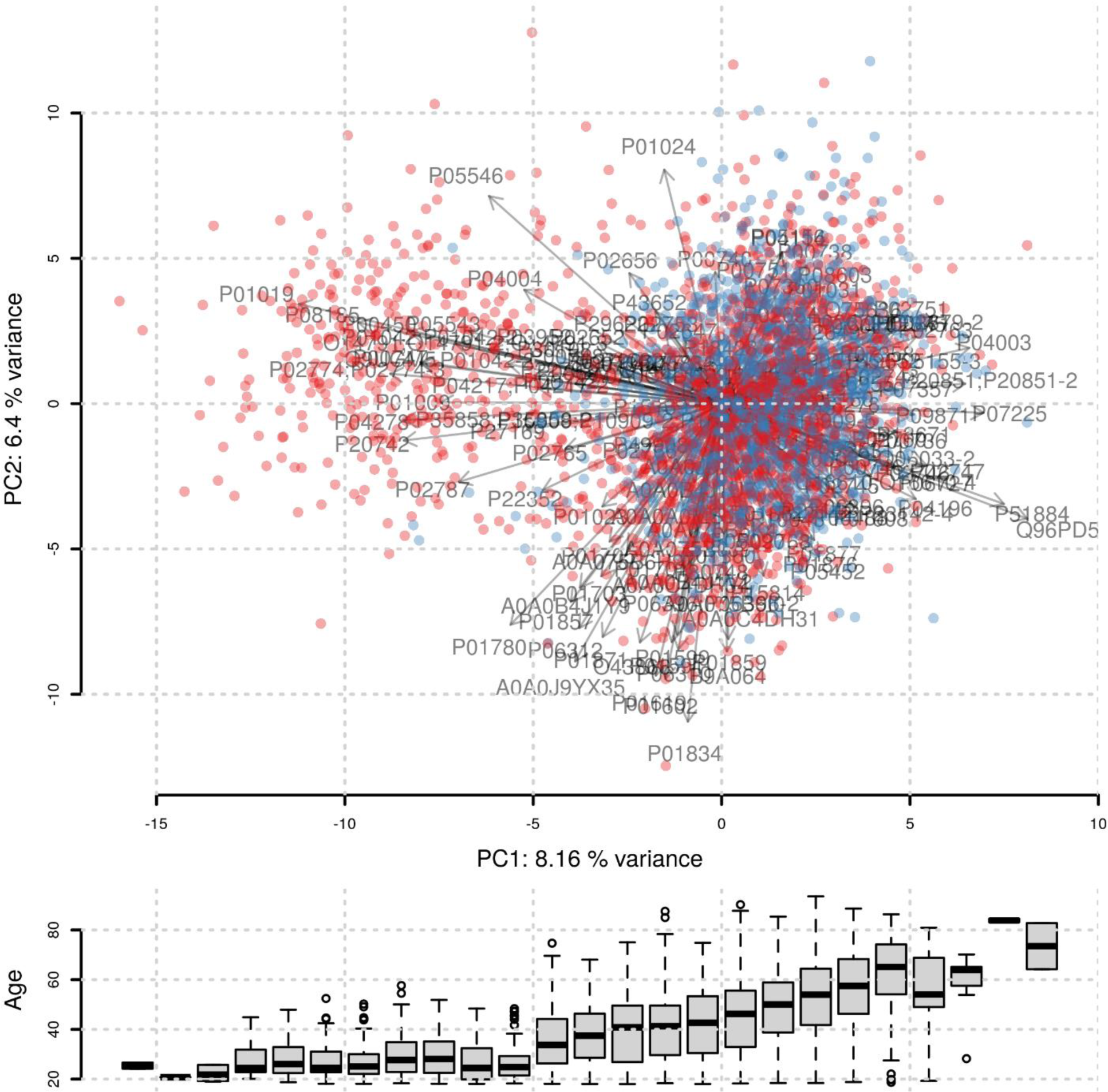
Principal Component Analysis of the CHRIS plasma proteome dataset. Individuals are colored by sex (red: female, blue: male). Loadings from the PCA are shown as arrows. Each arrow represents one protein with its length and direction indicating their impact and importance for that principal component. The lower panel shows the age distribution of participants along PC1.

By integrating phenotypic and medication data from the study participants, we found the subset of individuals separating on PC1 to be best characterized by a medication related to systemic oral hormonal contraceptives (ATC level 3 code G03A, hormonal contraceptives for systemic use, and ATC level 4 G03HB, antiandrogens and estrogens; Supplementary Figure S151). No other tested physiological parameter had a comparably strong influence on the plasma proteome. Thus, in the present data set, oral hormonal contraceptives are the major contributors to the variance observed on the quantified plasma proteome in a generally healthy population.

To identify the plasma proteome associated to hormonal contraceptives we next fitted multiple linear regression models to the proteomics data with explanatory variables for age, (categorical) BMI, (binary) fasting status and (binary) oral hormonal contraceptive use (HCU). To avoid any unwanted influence of age or sex on the results, we performed this analysis on the data subset of female participants below the age of 40 (n = 729, with 275 participants using hormonal contraceptives). As significance criteria we required for categorical variables, in addition to a statistical significance level, also the observed average difference in abundances to be larger than the CV of the respective protein determined on study specific quality control samples measured in the same data set. With this setting, we identified 50 plasma proteins that were significantly associated with the use of hormonal contraceptives, where the strongest association, and largest effect size, was found for AGT (angiotensinogen) (see Table 2). We further assessed the predictive power of AGT for HCU based on a sex, age, BMI, and fasting status-matched control group (n = 275) for the 275 female participants taking hormonal contraceptives. The AUROC (Area Under Receiver Operating Characteristic curve) for AGT was 89% confirming its high predictive ability (Figure 2B).

**Figure 2:**
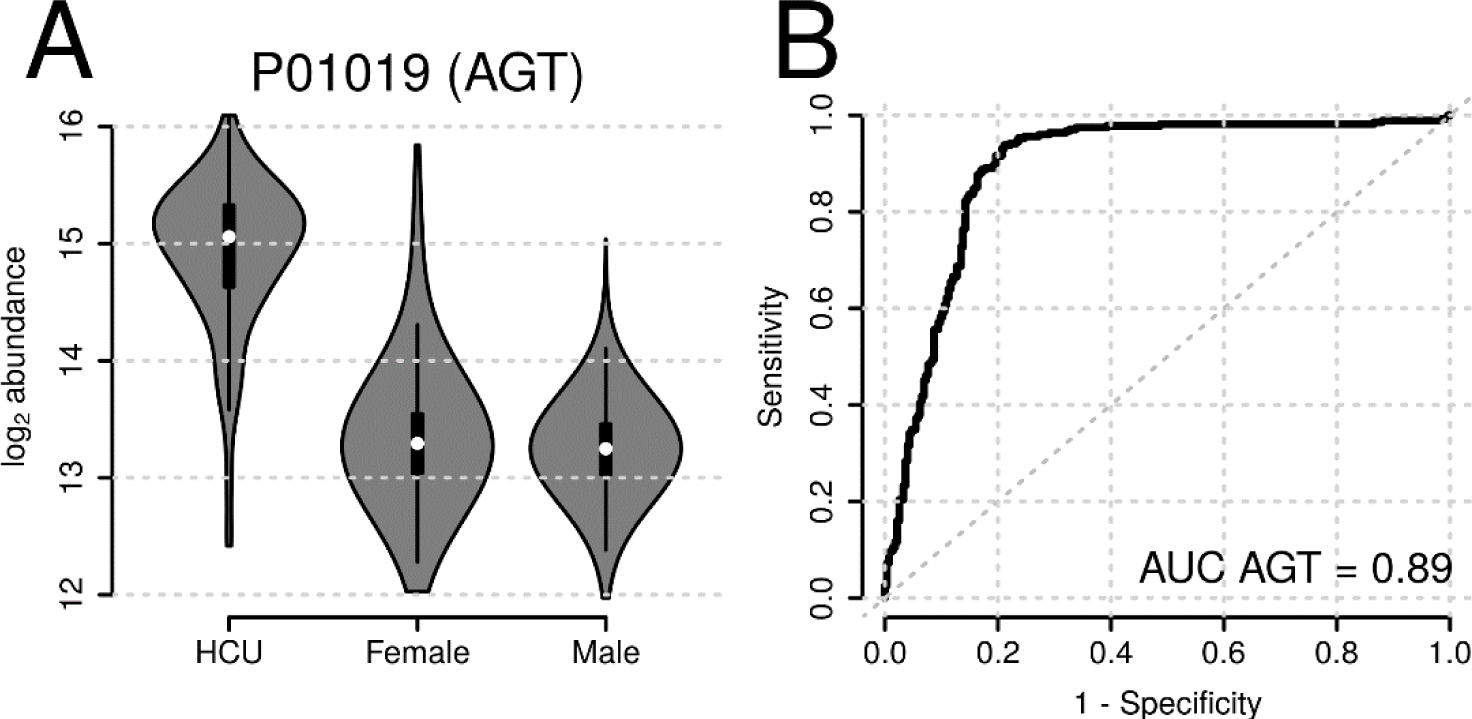
Abundance of the protein angiotensinogen (AGT) in study participants taking hormonal contraceptives (HCU) and female and male participants that don’t (A). ROC (Receiver Operating Characteristics) curve demonstrating the high predictive power of ACT for hormonal contraceptive usage (B).

**Table 2:** Proteins significantly associated with hormonal contraceptive use in females with age <= 40 years. Columns coef, p_adj_ and ES contain the abundance difference (in log2 scale), the p-value adjusted for multiple hypothesis testing and the effect size, respectively. Proteins are ordered by p-value.

| UniProt | Gene | coef | $p_{adj}$ | ES |
| --- | --- | --- | --- | --- |
| P01019 | AGT | 1.35 | 2.77e-95 | 1.38 |
| P00450 | CP | 0.552 | 1.14e-72 | 1.3 |
| Q96PD5 | PGLYRP2 | -0.624 | 5.39e-61 | -1.19 |
| P08185 | SERPINA6 | 0.822 | 5.43e-60 | 1.16 |
| P04278 | SHBG | 1.69 | 2.93e-59 | 1.16 |
| Q9UGM5 | FETUB | 1.22 | 6.38e-51 | 1.12 |
| P00747 | PLG | 0.286 | 1.14e-48 | 1.11 |
| P02774;P02774-3 | GC | 0.29 | 5.84e-46 | 1.06 |
| P01009 | SERPINA1 | 0.344 | 2.83e-44 | 1.04 |
| P01042;P01042-2 | KNG1 | 0.253 | 1.31e-42 | 1.05 |
| P01008 | SERPINC1 | -0.22 | 8.8e-38 | -0.967 |
| P51884 | LUM | -0.428 | 1.4e-36 | -0.979 |
| P05543 | SERPINA6 | 0.509 | 2.13e-36 | 0.96 |
| P01043-2 | KNG1 | 0.421 | 5.36e-35 | 0.961 |
| P05546 | SERPIND1 | 0.375 | 6.38e-35 | 0.954 |
| P04196 | HRG | -0.435 | 2.8e-34 | -0.907 |
| O14791;O14791-2;O14791-3 | APOL1 | 0.457 | 7.87e-29 | 0.875 |
| P13671 | C6 | -0.308 | 1.47e-27 | -0.868 |
| P04004 | VTN | 0.216 | 2.49e-25 | 0.777 |
| P07225 | PROS1 | -0.27 | 3.94e-25 | -0.823 |
| P20851;P20851-2 | C4BPB | -0.405 | 4.8e-25 | -0.825 |
| P02763 | ORM1 | -0.374 | 4.94e-25 | -0.783 |
| P05155;P05155-3 | SERPING1 | -0.864 | 3.67e-24 | -0.825 |
| P02753 | RBP4 | 0.27 | 6.76e-24 | 0.815 |
| P20742 | PZP | 0.707 | 4.54e-21 | 0.76 |
| P04003 | C4BPA | -0.227 | 3.49e-20 | -0.731 |
| P19652 | ORM2 | -0.333 | 1.67e-19 | -0.731 |
| P80108 | GPLD1 | 0.341 | 2.21e-19 | 0.74 |
| P05452 | CLEC3B | -0.244 | 6.23e-19 | -0.72 |
| Q06033;Q06033-2 | ITIH3 | -0.423 | 1e-18 | -0.724 |
| P02768 | ALB | -0.134 | 1.82e-16 | -0.686 |
| P02656 | APOC3 | 0.281 | 3.86e-16 | 0.685 |
| P07357 | C8A | -0.208 | 1.04e-14 | -0.657 |
| Q16610;Q16610-4 | ECM1 | -0.327 | 1.5e-14 | -0.651 |
| P02748 | C9 | -0.231 | 2.58e-14 | -0.638 |
| P06727 | APOA4 | -0.284 | 1.45e-13 | -0.619 |
| P01024 | C3 | 0.12 | 3.54e-13 | 0.572 |
| P00748 | F12 | 0.287 | 2.28e-12 | 0.594 |
| P02652 | APOA2 | 0.139 | 7.23e-12 | 0.594 |
| P02747 | C1QC | -0.204 | 1.4e-11 | -0.584 |
| P27169 | PON1 | 0.272 | 1.55e-11 | 0.59 |
| P02746 | C1QB | -0.191 | 9.25e-10 | -0.543 |
| P04217;P04217-2 | A1BG | 0.152 | 3.92e-09 | 0.534 |
| P00739 | HPR | 0.285 | 5.77e-07 | 0.47 |
| P02787 | TF | 0.11 | 1.78e-06 | 0.444 |
| O43866 | CD5L | -0.261 | 0.00037 | -0.377 |
| A0A075B6I0 | IGLV8-61 | -0.272 | 0.00519 | -0.329 |
| P01023 | A2M | -0.104 | 0.0171 | -0.287 |
| P00738 | HP | -0.17 | 0.0455 | -0.28 |
| P01871 | IGHM | -0.196 | 0.047 | -0.289 |

Similarly, we observed a clear and strong difference in abundance of AGT between females taking oral contraceptives and all other female as well as male participants when considering the overall study population (see Figure 2). This large difference also explained the observed bimodal signal distribution for this protein mentioned above.

Recently, a long-lasting effect of menopausal hormonal therapy (MHT) in the circulating proteome was reported (Thomas et al., 2022). To test whether also hormonal contraceptives would have a similar impact on the high abundant plasma proteome, we categorized female participants below 40 years of age into 3 groups: current use of hormonal contraceptives (n = 275), previous use of contraceptives (n = 280), and never used hormonal contraceptives (n = 76) and identified proteins with significant differences in abundances between these. Restricting the analysis to young females ensured balanced groups and reduced a potential influence of age and menopause on the results. In contrast to current use of hormonal contraceptives, not a single protein had significantly different concentrations between women with previous HCU and women that never took contraceptives (Supplementary Figure S152). Thus, in the present data set we could not observe any long-term effects of hormonal contraceptives on the plasma proteome.

### Plasma Proteome Associations to Age, Sex, BMI

To identify associations of proteins with age, sex, and body mass index (BMI) we next fitted multiple regression models to the abundances of each quantified plasma protein adjusting in addition for participants’ fasting status, and usage of oral hormonal contraceptives (the full results are provided in Supplementary Table S1).

With this analysis we identified 22 plasma proteins significantly differing between female and male participants (Supplementary Table S7; Figure 3A), with the top five candidates being the proteins RBP4, GPLD1, SERPINF1, and TTR showing lower, and CP higher abundance in females, respectively. Ninety-one proteins were found to be significantly related to the participants’ age (Supplementary Table S8; Figure 3B), with the proteins IGFALS and VTN having the strongest effects, showing decreasing abundances with higher age (see Supplementary Figures S153 and S154). We observed multiple proteins that had significantly different concentrations when comparing participants of BMI category 1 (underweight; BMI < 18.5; *n* = 48), BMI category 3 (overweight; 25 <= BMI < 30; *n* = 1,205), BMI category 4 (obese; BMI >= 30; *n* = 535) to participants from BMI category 2 (normal; 18.5 <= BMI < 25; *n* = 1,684). In total we observed 4 significantly associated proteins for underweight, 4 proteins for overweight and 20 proteins for obese status (Supplementary Tables S9, S10 and S11). Most of the BMI-associated proteins showed a difference in concentrations which was consistently increasing (or decreasing) with BMI (Figure 3C), such as SHBG and APOD having lower abundances with increasing BMI. Also, one protein, APOA4, was significantly associated with fasting status showing a 9% higher abundance in non-fasting participants (Supplementary Table S12).

**Figure 3:**
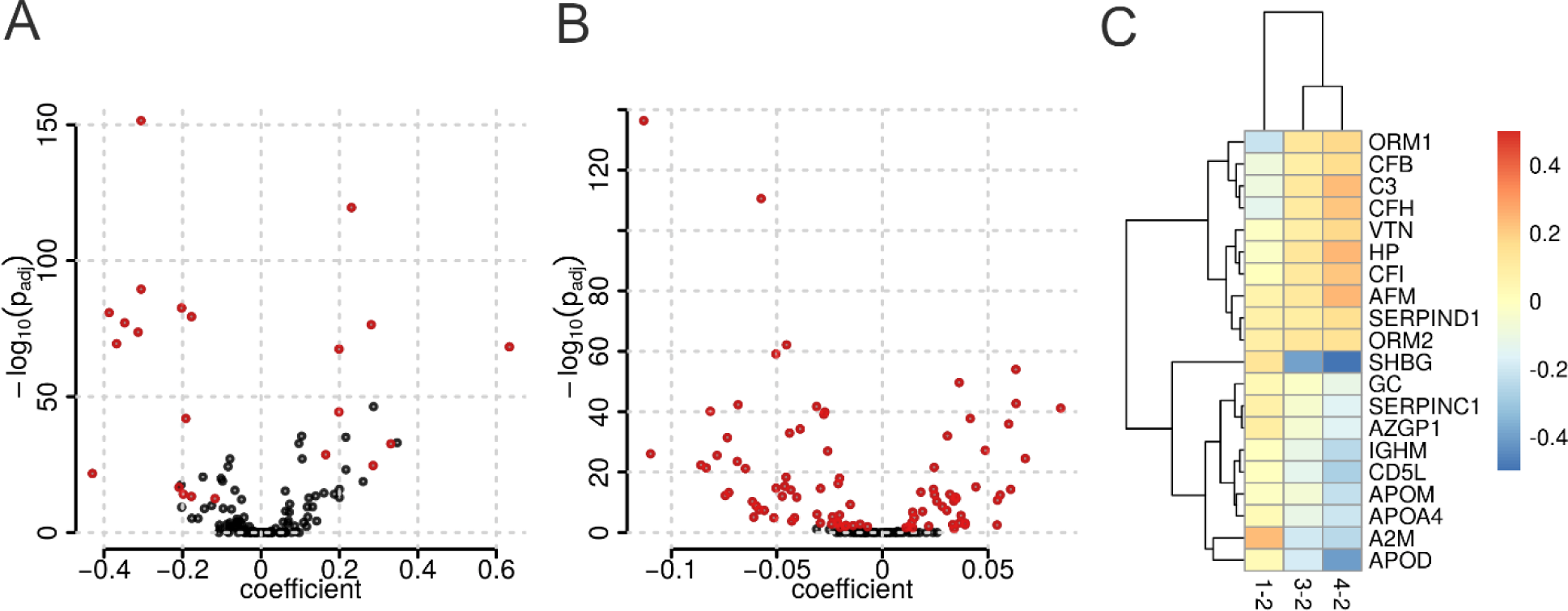
Sex-, age- and BMI-associated plasma proteins in the CHRIS study. A) Sex-associations of high abundant plasma proteins (Volcano plot). The coefficient represents the log2 difference in average concentrations between female and male participants. B) Age-dependency of high abundant plasma proteins (Volcano plot). The coefficient represents the log2-change in abundance over 10 years. Significant proteins are highlighted in red. C) Coefficients for proteins found to be significantly associated with at least one BMI category (Heatmap). The coefficients represent the log2-difference in average abundances between underweight, overweight, and obese (BMI categories 1, 3, and 4 respectively) compared to the reference normal weight (BMI category 2).

Off note, the results for HCU associations from this analysis were similar to the results from the analysis on the subset of female participants yielding almost identical coefficients and ranks of p-values and their effect sizes were larger than those for sex or age associated proteins (see Supplementary Figure S155).

Given the large impact of HCU on the plasma proteome, we evaluated to what extent adjustment for HCU influences the results of a general analysis for age, sex and BMI-associations. We thus conducted a sensitivity analysis by fitting the same linear models to the data omitting only the explanatory variable for HCU and compared the results of the two models. Indeed, 8 from the 28 sex- and 15 from the 101 age-associated proteins identified in this sensitivity analysis were significantly related to HCU but not to sex or age in the full analysis model (see Supplementary Tables S13 and S14 for coefficients and p-values for these proteins in both analyses). In contrast, BMI-associations were not affected. Thus, the adjustment for HCU had a clear impact on the age and sex-association results while it did not affect the BMI-associations (see Figure 4).

**Figure 4:**
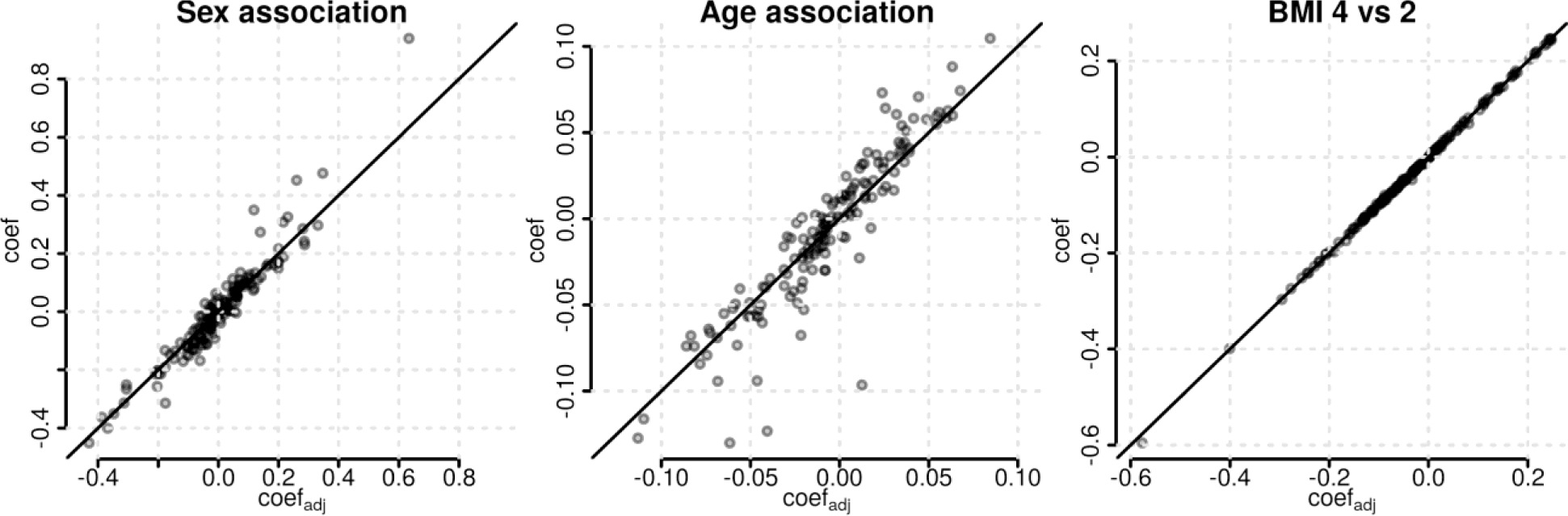
Sensitivity analysis for sex, age, and BMI-associations of plasma proteins. Shown are the coefficients from the linear model adjusting for hormonal contraceptive use (x-axis) against the coefficients from a linear model without that adjustment (y-axis). The solid black line represents the identity line.

### Influence of Medication on Quantified Plasma Proteins

To evaluate whether also other medications influence the abundances of plasma proteins, we next determined the most common medications in the present data set and evaluated their impact on the high abundant plasma proteome. To identify associations between plasma proteins and therapeutical subgroups of general medication, we first identified all ATC level 3 medications taken by at least 15 participants (0.4% of the sample set) on a regular basis (at least two times per week) and defined a binary variable for each of them. These were then included as explanatory variables into the per-protein multiple regression models, that accounted also for age, sex, (categorical) BMI and (binary) fasting status of each study participant. The tested medications along with the number of participants as well as significant protein associations are shown in Table 3.

**Table 3:** Overview of association results for ATC level 3 medications in the CHRIS study subset. Only medications taken on a regular basis by more than 14 of the in total 3,632 participants were considered. Columns *Participants* and *Proteins* list the number of participants taking the medication and number of significantly associated proteins.

| ATC | Name | Participants | Proteins |
| --- | --- | --- | --- |
| **G03A** | HORMONAL CONTRACEPTIVES FOR SYSTEMIC USE | 286 | 50 |
| **H03A** | THYROID PREPARATIONS | 262 | 0 |
| **B01A** | ANTITHROMBOTIC AGENTS | 243 | 2 |
| **C10A** | LIPID MODIFYING AGENTS, PLAIN | 203 | 1 |
| **C09A** | ACE INHIBITORS, PLAIN | 156 | 0 |
| **C07A** | BETA BLOCKING AGENTS | 149 | 1 |
| **N06A** | ANTIDEPRESSANTS | 141 | 0 |
| **C09D** | ANGIOTENSIN II ANTAGONISTS, COMBINATIONS | 113 | 1 |
| **A02B** | DRUGS FOR PEPTIC ULCER AND GASTRO-OESOPHAGEAL<br>REFLUX DISEASE (GORD) | 94 | 0 |
| **C08C** | SELECTIVE CALCIUM CHANNEL BLOCKERS WITH MAINLY<br>VASCULAR EFFECTS | 91 | 0 |
| **C09C** | ANGIOTENSIN II ANTAGONISTS, PLAIN | 74 | 1 |
| **A12A** | CALCIUM | 65 | 2 |
| **C09B** | ACE INHIBITORS, COMBINATIONS | 59 | 0 |
| **A10B** | BLOOD GLUCOSE LOWERING DRUGS, EXCL. INSULINS | 57 | 4 |
| **M01A** | ANTIINFLAMMATORY AND ANTIRHEUMATIC PRODUCTS,<br>NON-STEROIDS | 48 | 3 |
| **G02B** | CONTRACEPTIVES FOR TOPICAL USE | 46 | 19 |
| **G04C** | DRUGS USED IN BENIGN PROSTATIC HYPERTROPHY | 46 | 4 |
| **M04A** | ANTIGOUT PREPARATIONS | 36 | 2 |
| **N03A** | ANTIEPILEPTICS | 34 | 1 |
| **N05C** | HYPNOTICS AND SEDATIVES | 32 | 0 |
| **S01E** | ANTIGLAUCOMA PREPARATIONS AND MIOTICS | 30 | 0 |
| **R03A** | ADRENERGICS, INHALANTS | 29 | 1 |
| **N05B** | ANXIOLYTICS | 24 | 0 |
| **N05A** | ANTIPSYCHOTICS | 21 | 0 |
| **G03F** | PROGESTOGENS AND ESTROGENS IN COMBINATION | 18 | 0 |
| **G03H** | ANTIANDROGENS | 16 | 38 |
| **N04B** | DOPAMINERGIC AGENTS | 16 | 1 |

Among the 27 tested ATC3 medications, most frequent were hormonal contraceptives for systemic use (ATC3 G03A), thyroid preparations (ATC3 H03A), antithrombotic agents (ATC3 B01A) and lipid modifying agents (ATC3 C10A), each with more than 200 participants taking these on a regular basis. In line with results from the previous sections, medications related to contraception (ATC3 G03A, G03H and G02B) yielded by far the highest number of significantly associated plasma proteins (50, 38 and 19, respectively, see Table 3). For these medications we observe a considerable overlap of the protein signatures as well as similar effect sizes (see Figure 5). Even contraceptives for topical use (ATC3 G02B), which was not considered in the definition of the oral hormonal contraceptive use (HCU) variable in the previous section, showed, despite smaller effect sizes, a similar protein signature. For the remaining medications, no or only few significant protein associations were found (see Table 3). Tables with significant proteins for each medication are provided in the supplement (Supplementary Tables S15-S30).

**Figure 5:**
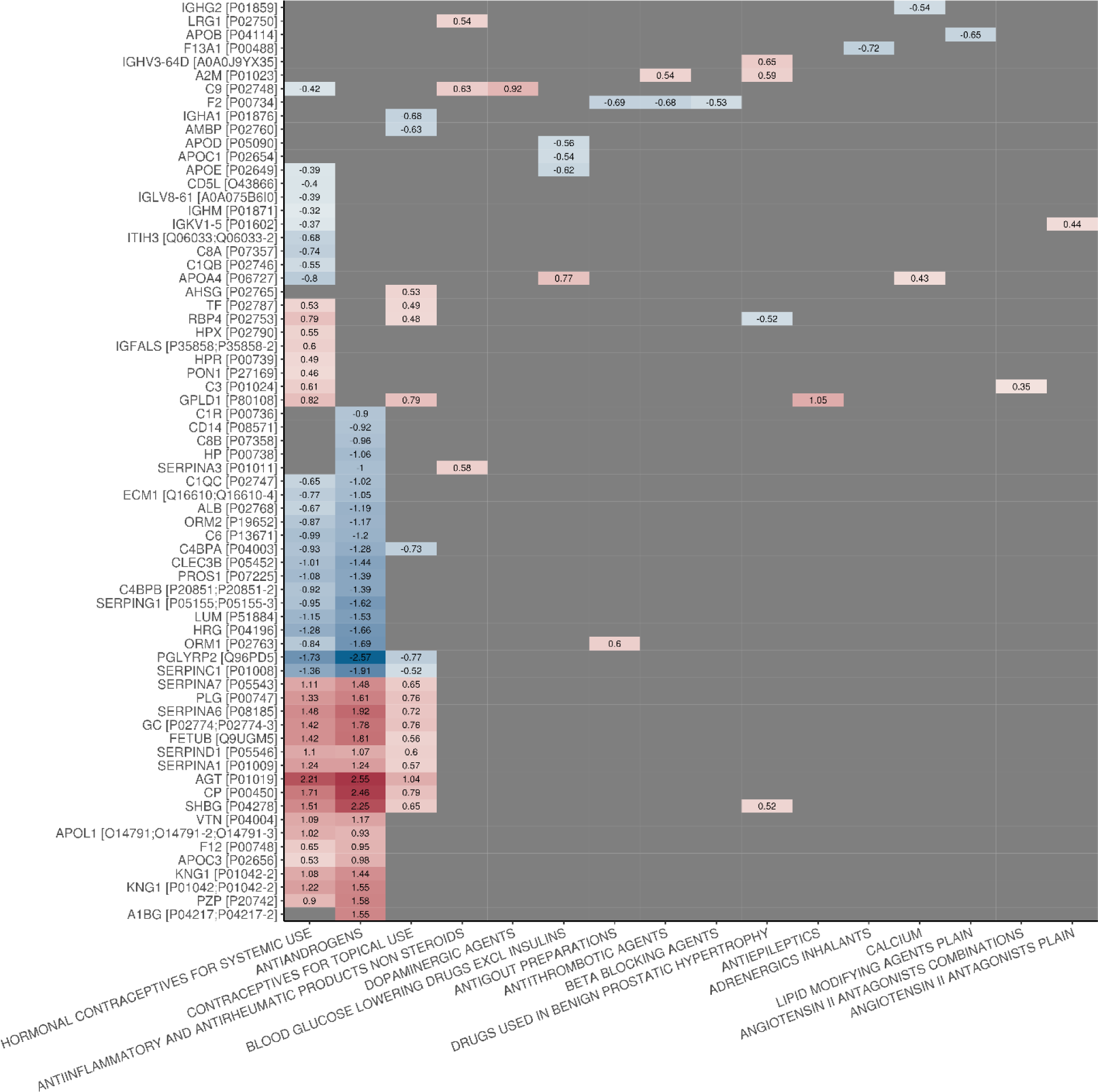
Significant associations between proteins (rows) and ATC level 3 medications (columns). Effect sizes (as a number and color coded) are only shown for significant associations.

We next repeated the analysis on ATC level 4 medications to eventually identify additional associations with the more specific chemical or pharmacological subgroups defined by this ATC level. Also in this analysis, medications related to hormonal contraceptives yielded the highest number of significant protein associations: 52, 38, 36 and 35 for progestogens and estrogens, fixed combinations (ATC4 G03AA), antiandrogens and estrogens (ATC4 G03HB), progestogens and estrogens, sequential preparations (ATC4 G03AB) and intravaginal contraceptives (ATC4 G02BB), respectively (see Table S31). Further, the signatures and effect sizes were highly similar for these medications, irrespective of the route of administration (Supplementary Figure S156): intravaginal contraceptives had a similar signature and effect sizes than all other, orally administered, hormonal contraceptives. In contrast, no significant protein was found for the ATC4 medication intrauterine contraceptives (ATC4 G02BA), which is part of the same ATC level 3 medication subgroup (ATC3 G02B, contraceptives for topical use) as intravaginal contraceptives (ATC4 G02BB). The medication subgroup with the next most significant proteins (14) was vitamin K antagonists (ATC4 B01AA), while for platelet aggregation inhibitors excl. heparin (ATC4 B01AC, part of the same ATC3 therapeutic subgroup antithrombotic agents) only a single significant association was found. For the remaining medications, only few significant proteins were identified, and no significant association was detected for 13 of the in total 29 tested medication subgroups (see Supplementary Tables S32-S44 for significant proteins for the tested medications).

Thus, summarizing, hormonal contraceptives had, among all tested medications, by far the strongest influence on the quantified plasma proteome in this study.

## Discussion

In this study, we explored the high abundant plasma proteome fraction of 3,632 CHRIS study participants using proteomics. Using a combination of semi-automated sample preparation of neat plasma, fast analytical flow-rate chromatography, and Scanning SWATH (Messner et al., 2021), we recorded the proteomes for over 5,125 plasma samples (including QCs). Using an HPLC setup with two binary pump systems, one gradient and one wash/equilibration pump, reduced overheads to 1.8 min and eventually allowed one operator to run 3 LC-MS batches (3x 96-well plates each) per week and to complete the measurements within 4 weeks of instrument time. We obtained precise quantities and limited batch effects, demonstrating that mass spectrometry-based proteomics suits the quantification of the high abundant plasma proteomic fraction in large-scale epidemiological cohorts. Applying strict filtering criteria, we report a set of 148 reliably measured proteins considering all study samples. We would like to note that in other applications of our plasma proteomic platform a higher proteomic depth was achieved, albeit in lower sample numbers (Demichev et al., 2021; Vernardis et al., 2023). We speculate that the sample matrix or a confounding technical factor (like plastic materials), have affected tryptic digestion and hence proteomics depths achieved in the present data set. However, the quantitative precision of the obtained data was high, thus allowing us to test for associations between the high abundant plasma proteome and epidemiological parameters.

Many of the high abundant plasma proteins identified were either members of the complement system and innate immunity, coagulation factors, or immunoglobulins, and thus representing processes with high disease relevance. Indeed, a considerable number of these proteins varied significantly across individuals and at least 31 of them are linked to disease biomarkers, reflecting the high importance of the upper plasma protein fraction for biomarker discovery.

A general comparison of quantified protein abundances with typical laboratory tests yielded an overall significance concordance, considering that the assays test for different parameters. While the proteomics platform assesses protein abundance projecting to the average proteoform, the diagnostic tests either test for total protein, enzyme activity or assess protein complexes like LDL or HDL. Given these constraints, the related tests yielded significant correlations. For instance, the correlation coefficients were high for some proteins (0.78 between APOB and HDL, 0.68 for transferrin) while APOA1 levels correlated to a somewhat lower degree with the LDL, of which APOA1 is a component (0.41).

Our plasma proteome reflected both known and new associations with epidemiological parameters. For instance, when comparing to literature, 12 from the 22 sex associated proteins identified in our study, have been described using other methods. Reassuringly, for most, these show the same directionality of the effect (Cho et al., 2006; Ding et al., 2012; Gaya da Costa et al., 2018; Hammond, 2011; Lehallier et al., 2019; Lin et al., 2013; Lyutvinskiy et al., 2013; Miike et al., 2010), whereas for GPLD1 an opposite effect was reported (Lehallier et al., 2019) (see Supplementary Table S45). Nine proteins which showed a sex association in our dataset (TTR, AZGP1, HBA1, LRG1, APOA4 IGHA1, CD5L, APOD) have not been identified as sex dependent in previous studies, to the best of our knowledge. Our dataset revealed 91 age-associated protein abundances. Of these, 63 have been previously associated with ageing in the literature (Cominetti et al., 2018; Larsson et al., 2020; Lehallier et al., 2019; Orwoll et al., 2020; Siino et al., 2022; Tanaka et al., 2020; M. Xu et al., 2020; R. Xu et al., 2020). About 80% of these (n = 50) showed a similar pattern of regulation in our study. Of the remaining 29 proteins, 28 were previously reported to have no statistically significant correlation with age (Siino et al., 2022; Tanaka et al., 2020; R. Xu et al., 2020). However, despite the lack of statistical significance in previous studies, 75% of these showed a similar trend in fold change as observed in our study (see Supplementary Table S45). Regarding BMI, we identified 20 proteins with significant differences in concentration between the various BMI categories and the reference level. Among these, 15 have been previously reported for BMI, with same directionality, based on plasma proteomes from large population studies such as KORA, INTERVAL, and QMDiab (Goudswaard et al., 2021; Zaghlool et al., 2021). Finally, a lower overlap with results from literature was found for proteins that were significantly associated with hormonal contraceptive use in our data (22 out of 50) (Josse et al., 2012; Ramsey et al., 2016). In proteomics, comparisons with results from literature generally suffer from a low overlap and coverage of measured proteins due to the different employed platforms (various MS-based protocols, different affinity proteomics approaches) resulting in gaps and missing reference values. This was particularly the case with Enroth et al. (Enroth et al., 2018), where the intersection of analytes was limited to a single commonly measured protein, thus precluding an extensive confrontation of results. Arguably, differences in study sizes as well as employed statistical tests and significance cut-offs will impact comparisons with literature. Taken together, the overall acceptable coefficient of variation of the data set together with the correlation of protein abundances with estimates from independent laboratory tests on the same blood samples and the agreement of results from the investigated traits with previous results from literature suggest the present MS-based plasma proteome data set to be of high quality and proves its utility to identify biologically and clinically interesting signals.

In our data set, hormonal contraceptives explained most of the variance of the quantified plasma proteome. Indeed, an impact of oral contraceptives, and to a lower extend also of the menstrual cycle, on plasma protein and metabolite abundances has also been reported previously (Klipping et al., 2021; Ramsey et al., 2016), yet, the use of hormonal contraceptives is not yet systematically accounted for in the typical epidemiological studies. In the liver, oral contraceptives stimulate the synthesis of steroid-binding globulins, such as sex hormone-binding globulin (SHBG), thereby affecting circulating, free steroid levels and they further increase low-grade inflammation, alter lipid metabolism and affect the coagulation system, resulting in an increased risk for thromboembolic events (Kangasniemi et al., 2023). We were able to independently validate these findings using our extensive dataset, which identifies a substantial number of plasma proteins that are significantly impacted by the use of oral hormonal contraceptives. In addition to oral contraceptives, also (hormonal) intravaginal contraceptives resulted in an almost identical protein signature with highly similar effect sizes. The effect of hormonal contraceptives on the plasma proteome is thus independent of the route of administration. Also, compared to the other analyzed traits (age, sex, BMI), the numbers of significant proteins and related effect sizes were much larger for hormonal contraceptive use. Notably, levels in angiotensinogen (AGT), the most discriminatory protein, separated users and non-users of hormonal contraceptives, and might serve as a biomarker of contraceptive use. Indeed, exogenous estrogens have been reported to cause upregulation of hepatic angiotensinogen (Elger et al., 2017; Gordon et al., 1992) associated with an activation of the renin angiotensin system with however little renal and systemic consequences (Cherney et al., 2007; Kang et al., 2001). A sensitivity analysis unequivocally underscored the necessity of incorporating hormonal contraceptive use into the analytical models, in order to prevent proteins influenced by this treatment from being erroneously linked to factors such as age, sex, or correlated phenotypes. Recently, a long-term effect of menopausal hormonal therapy on the circulating plasma proteome was described (Thomas et al., 2022). We, however, could not identify any such effect for hormonal contraceptives on the high abundant plasma proteome, possibly, due to differences in hormone composition and dosage between the two medications. In addition, the intersection of quantified proteins in both studies is very small (22 proteins) and, except for SERPINA3, none of the proteins from the reported signature was detected in our data set. Thus, hormonal contraceptives represent a large influence factor on plasma protein concentrations and should be considered, in addition to clinical practice as suggested by Ramsey et al. (Ramsey et al., 2016), in any study involving plasma proteomics data sets, particularly if the analyzed trait might be related to either age or sex. For example, hormonal contraceptive use should be accounted for in an association analysis for cardiovascular health in the present data set to avoid confounded results.

In contrast to hormonal contraceptives, the majority of the commonly used medications among the participants in the CHRIS study were those prescribed for the management of cardiometabolic conditions. None of these drugs had a similarly strong influence on plasma protein levels but affected specific pathways. The disparity in concentration observed for these medications was notably less pronounced than that seen with hormonal contraceptives. For instance, lipid-modifying agents were found to influence APOB levels, and it was this specific protein that exhibited a statistically significant association with the medication in our study.

To conclude, we assessed the potential of the high abundant fraction of the plasma proteome in an epidemiological study, the CHRIS cohort. We identified associations between protein abundances and common factors such as sex and age, but notably, report that hormonal contraceptives had the largest effect on the plasma proteome. Due to its high prevalence, and apparently strong influence, this type of medication should be accounted for in any epidemiological or clinical study of the plasma proteome to avoid spurious findings. Moreover, future studies should clarify to which degree desired and undesired effects resulting from the use of contraceptives are associated with the changes in the plasma proteome.

## Material and Methods

### Study cohort

Details on the CHRIS study including recruitment are given in (Pattaro et al., 2015). In brief, study participants were recruited from the adult population of the middle and upper Vinschgau/Val Venosta district located in the mountainous northern-most region of Italy. Next to collection and subsequent biobanking of blood and urine samples a self-reported, questionnaire-based health assessment was performed. Medication information was collected by scanning the barcode of the medication boxes study participants brought along and assignment of the respective Anatomical Therapeutic Chemical (ATC) codes. Standard blood parameters were measured in blood samples at the Hospital of Merano using standardized clinical assays. Details on measurements of clinical laboratory parameters for the present study set including the description of sample handling are described in (Noce et al., 2017). In brief, antithrombin was measured in plasma citrate samples using the enzymatic Siemens Innovance Antithrombine assay on a ROCHE SYSMEX CA1500 and for a subset of participants using the STA-Stachrom AT III assay on a STAGO STA COMPACT MAX instrument. Albumin, HDL, LDL, Triglycerides and transferring were measured in serum samples using the colorimetric Cobas ALB plus Albumin BCG assay, the enzymatic Cobas HDL-C Plus 3 generation assay, the enzymatic Cobas LDL-C plus 2^nd^ generation assay, the Cobas Triglyceride GPO-PAP assay and the immunological Cobas Tina-quant Transferrin ver.2 assay, respectively, on a ROCHE MODULAR PPE and for a subset of samples using the colorimetric ALBUMIN BCG assay, the ULTRA HDL assay, the DIRECT LDL assay, the TRIGLYCERIDE assay and the immunological TRANSFERRIN assay, respectively, on an ABBOT DIAGNOSTIC ARCHITECT. Hemoglobin (HGB) was measured in EDTA plasma using the electronic impedance laser light scattering based assay on an ABBOT CD SAPPHIRE instrument and on a subset of samples on a SYSMEX XN-1000.

From the in total 13,393 participants of the CHRIS study 3,632, participating to the study between August 2011 and August 2014, were selected for mass spectrometry-based quantification of their plasma proteome.

### Sample Preparation

In this study, a total of 5,125 samples were subjected to measurement. Among these, 479 samples were quality control samples, 498 were plasma set samples and 200 were pooled study samples that were utilized to monitor measurement quality and control technical variation. The measurement comprised of 3,948 study samples including 350 samples from a CHRIS sub-study, which were, however, excluded from the present data analysis. These plasma citrate samples were randomly distributed across 50 96-well plates, together with 4 study pools, 4 commercial serum (ZenBio SER-SPL) samples, and 8 plasma samples (ZenBio HSER-P500ML) per plate. Measurements were performed in 17 batches, each consisting of 3 plates (except for MS batch 17), over a span of 6 months.

Semi-automated in-solution digestion was performed as previously described for high throughput clinical proteomics (Messner et al., 2020). All stocks and stock plates were prepared in advance to reduce variability and were stored at −80°C until use. Briefly, 5 μl of thawed samples were transferred to the denaturation and reduction solution (50 μl 8 M Urea, 100 mM ammonium bicarbonate (ABC), 5 µl 50 mM dithiothreitol per well) mixed and incubated at 30°C for 60 minutes. Five microliters were then transferred from the iodoacetamide stock solution plate (100 mM) to the sample plate and incubated in the dark at RT for 30 minutes before dilution with 100 mM ABC buffer (340 μl). 220 μl of this solution was transferred to the pre-made trypsin stock solution plate (12.5 μl, 0.1 μg/μl) and incubated at 37°C for 17 h (Benchmark Scientific Incu-Mixer MP4). The digestion was quenched by addition of formic acid (10% v/v, 25 μl) and cleaned using C18 solid phase extraction in 96-well plates (BioPureSPE Macro 96-Well, 100 mg PROTO C18, The Nest Group). The eluent was dried under vacuum and reconstituted in 60 μl 0.1% formic acid. Insoluble particles were removed by centrifugation and the samples transferred to a new plate.

### Liquid Chromatography and Mass Spectrometry

The digested peptides were separated on a 5-min high-flow chromatographic gradient and recorded by mass spectrometry using Scanning SWATH (Messner et al., 2021) on an Agilent Infinity II HPLC combined with a SCIEX 6600 TripleTOF platform. Five micrograms of sample were injected onto a reverse phase HPLC column (Luna®Omega 1.6µm C18 100A, 30 × 2.1 mm, Phenomenex) and resolved by gradient elution at a flow rate of 800 µl/min and column temperature of 30⁰C. All solvents were of LC-MS grade. The fast separation used 0.1% formic acid in water (Solvent A) and 0.1% formic acid in acetonitrile (Solvent B) using an alternating column regeneration system where the gradient separation of one sample is performed on one LC column by a gradient pump while a second identical column is being washed and equilibrated using a regeneration pump. The gradient separation, wash and equilibration programs are described in Supplementary Table S46. For mass spectrometry analysis, the scanning SWATH precursor isolation window was 10 m/z, the bin size was set to 20% of the window size, the cycle time was 0.52 s, the precursor range was set to 400 – 900 m/z, the fragment range to 100 – 1500 m/z as previously described in Messner et al. (Messner et al., 2021). A Sciex IonDrive TurboV source was used with ion source gas 1 (nebulizer gas), ion source gas 2 (heater gas) and curtain gas set to 50 psi, 40 psi and 25 psi, respectively. The source temperature and ion spray voltage were set to 450⁰C and 5500 V, respectively.

### Data Processing and Statistical Data Analysis

Raw MS data was processed using DIA-NN v1.8 (Demichev et al., 2020). Although DIA-NN can optimize mass accuracies and the scan window size, we fixed them to ensure the reproducibility of our results (MS1: 12 ppm; MS2: 20 ppm; scan window size: 6). An external, publicly available spectral library was used for all measurements (Bruderer et al., 2019). The spectral library was annotated using the Human UniProt (UniProt Consortium, 2008) isoform sequence database (Proteome ID: 3AUP000005640).

All preprocessing steps of the DIA-NN output matrix were performed in the R programming language (v4.0.4). All libraries used were in compatible versions to the R version used. Imputation of missing values was performed using the knn function implemented in the impute package, with k = 9 nearest neighbors applied to samples within each MS batch. Data were normalized using cyclic loess (with the ‘fast’ option) (Ballman et al., 2004; Bolstad et al., 2003) and plate effects were corrected using the removeBatchEffect function from the limma Bioconductor package (Ritchie et al., 2015). To map peptide precursors to proteins, precursor filtering (retaining only proteotypic precursors) and median polish summarization were applied, as implemented in the preprocessCore Bioconductor package. Functional analysis was performed with gProfiler package (Kolberg et al., 2020). Only GO Terms with an FDR-corrected p-value > 0.05 were considered significant. Protein class information was obtained from the PANTHER Classification System (Mi et al., 2013). The results of the functional annotations are summarized in Supplementary Table S2. To assess the predictive ability of AGT, a control group was defined using the MatchIt package with an “optimal” matching strategy and calculation of propensity scores by a generalized linear model. ROC curves were calculated by using the pROC package.

To compute the CV for each protein or peptic precursor, the empirical standard deviation (the square root of the variance) was divided by the empirical mean (the average abundance), and the result was expressed as a percentage. For PCA analysis, protein abundances were scaled to zero mean and standard deviation of one (autoscaling or z-score transformation). To identify associations with sex, age, BMI, fasting status, and hormonal contraceptive use, linear regression models were fitted separately for each protein using its concentration as response variable and sex, age, fasting status, BMI and hormonal contraceptive use as covariates. For easier interpretation of relative effects, participants’ age is divided by 10, thus age-related coefficients and effect sizes are related to 10 years of difference (Steyerberg, 2019). For BMI, clinical categories were used (WHO Consultation on Obesity (1999: Geneva and Organization, 2000): underweight (category 1, BMI < 18.5), normal range (category 2, 18.5 ≤ BMI < 25), overweight (category 3, 25 ≤ BMI < 30) and obese (category 4, BMI > 30). For fasting status, a binary variable based on the self-reported fasting information from the questionnaire was used (1 for participants declaring to have had a meal within the 12 hours prior blood drawing and 0 for all others). Medication information was recorded by scanning the medication boxes participants were asked to bring along to the visit at the study site and Anatomical Therapeutic Chemical (ATC) codes of the medications were extracted. A binary variable for oral hormonal contraceptives was defined using ATC level 3 categories “HORMONAL CONTRACEPTIVES FOR SYSTEMIC USE” (ATC3 G03A) and “ANTIANDROGENS” (ATC3 G03H). To evaluate influence on common medication on plasma protein levels, age, sex, BMI and fasting status-adjusted linear models were fitted including in addition explanatory variables for ATC3 or ATC4 medications taken on a regular basis by at least 15 participants. P-values from linear models were adjusted for multiple testing using the Bonferroni method. Proteins with an adjusted p-value smaller than 0.05 were considered statistically significant. In addition, for categorical variables (such as sex, age, BMI, or medications), to call a protein significant, its absolute difference in concentration for the variable had to be larger than its CV across the sample pools. The observed difference in concentrations is thus required to be larger than the technical variability that was observed for that protein in the present data set (calculated on the QC CHRIS Pool samples).

Categorization of female participants into groups “current HCU”, “previous HCU” and “never used any hormonal contraceptives” was based on self-reported questionnaire data combined with the definition of hormonal contraceptive use described above. Females with missing or ambiguous information were excluded from the analysis. To identify proteins with significant differences in abundances between these categories, linear models were fitted to the data adjusting in addition for age, BMI, and fasting status.

Additionally, linear regression models were fitted to protein concentrations standardized to mean of 0 and standard deviation of 1. Coefficients from this analysis, where differences in one unit are equal to a standard deviation of 1, are comparable and reported as “effect size”. All analyses were performed on log2-transformed protein concentrations.

Data analysis was performed in R (version 4.2.2), R markdown documents defining and describing the analysis are available on github (https://github.com/EuracBiomedicalResearch/chris_plasma_proteome).

## Data Availability Statement

CHRIS study data can be requested for research purposes by submitting a dedicated request to the CHRIS Access Committee. Please contact for more information on the process.

## Supporting information

Document (pdf) with Supplementary Figures S1-S156 and Supplementary Tables S3-S44.

Spread sheet (xlsx) with Supplementary Table S1 (results from association analyses).

Spread sheet (xlsx) with Supplementary Table S2 (functional annotations of detected proteins).

Spread sheet (xlsx) with Supplementary Table S45 (comparison of results with literature).

## Acknowledgements

The CHRIS study is a collaborative effort between the Eurac Research Institute for Biomedicine and the Healthcare System of the Autonomous Province of Bozen/Bolzano (SüdtirolerSanitätsbetrieb/Azienda Sanitaria dell’Alto Adige). The investigators thank all study participants from the middle and upper Vinschgau/Val Venosta, the general practitioners, the personnel of the Hospital of Schlanders/Silandro, the field study team and the personnel of the CHRIS Biobank (BRIF code BRIF6107) for their support and collaboration. The authors thank the Department of Innovation, Research University and Museums of the Autonomous Province of Bozen/Bolzano for covering the Open Access publication costs. We thank the Charité Core Facility High Throughput Mass Spectrometry, especially Daniela Ludwig for sample preparation. The graphical abstract was created with biorender.com.

## Institutional Review Board Statement

The study was conducted according to the guidelines of the Declaration of Helsinki and approved by the Ethics Committee of the Health Authority of the Autonomous Province of Bolzano (Südtiroler Sanitätsbetrieb/Azienda Sanitaria dell’Alto Adige; protocol No. 21/2011, 19 April 2011).

## Informed Consent Statement

Informed consent was obtained from all subjects involved in the study.

## Author Contributions

Conceptualization: J.R., M.R., N.D., C.D., E.H.; Investigation: V.V.H., F.A., A.D., M.M.; Formal Analysis: N.D., C.D., J.R. V.F., M.M.; Writing – original draft preparation: E.H., C.D., J.R., N.D., F.A.; Writing – review and editing: N.D., C.D., E.H., M.M., V.F., V.V.H., A.D., F.S.D., P.P.P., M.R., J.R.; Funding acquisition: M.R., P.P.P..

## Supplementary Information

Document (pdf) with Supplementary Figures S1-S156 and Supplementary Tables S3-S44. Spread sheet (xlsx) with Supplementary Table S1 (results from association analyses). Spread sheet (xlsx) with Supplementary Table S2 (functional annotations of detected proteins). Spread sheet (xlsx) with Supplementary Table S45 (comparison of results with literature).

