## Supplementary material for "Pervasive Influence of Hormonal Contraceptives on the Human Plasma Proteome in a Broad Population Study": Document (pdf) with Supplementary Figures S1-S156 and Supplementary Tables S3-S44.

### Supplementary Information for: *Hormonal Contraceptives are Shaping the Human Plasma Proteome in a Large Population Cohort*

**Plasma proteins associated with medication (ATC level 3)** **49**

**Plasma proteins associated with medication (ATC level 4)** **54**

**Extended Material and Methods** **62**

#### General data overview

##### Signal distributions of individual proteins

Distribution of $log_2$ transformed signal distributions in the analyzed samples is shown for each protein.

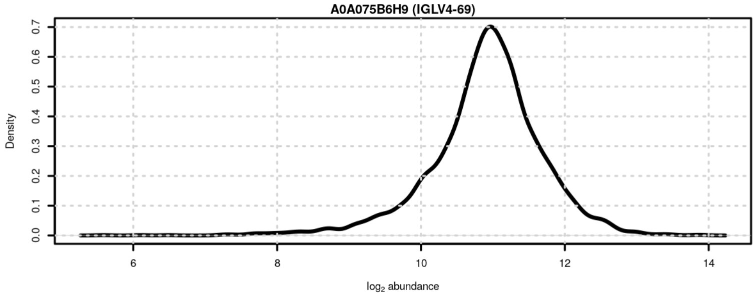

**Figure S1**: Signal distribution for A0A075B6H9 (IGLV4-69).

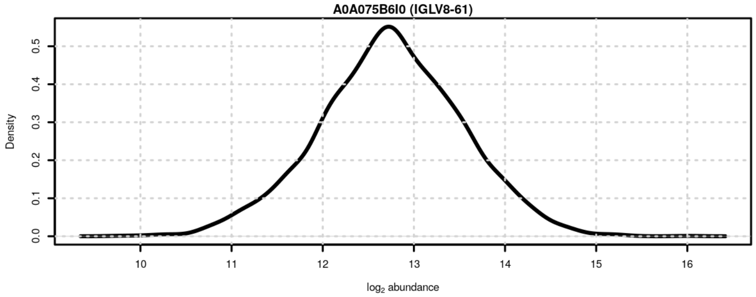

**Figure S2**: Signal distribution for A0A075B6I0 (IGLV8-61).

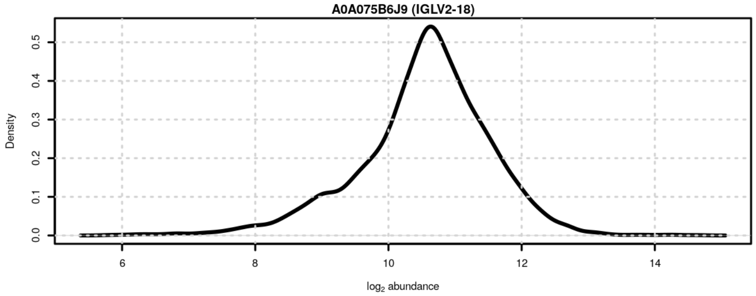

**Figure S3**: Signal distribution for A0A075B6J9 (IGLV2-18).

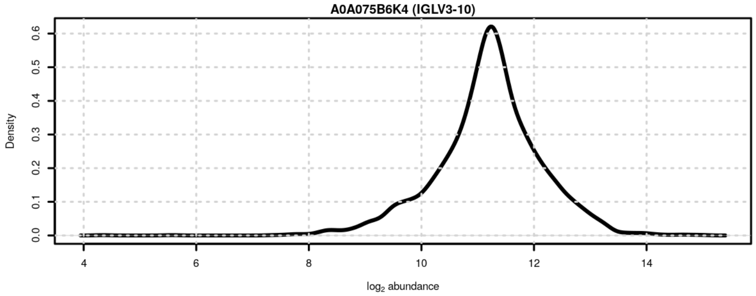

**Figure S4**: Signal distribution for A0A075B6K4 (IGLV3-10).

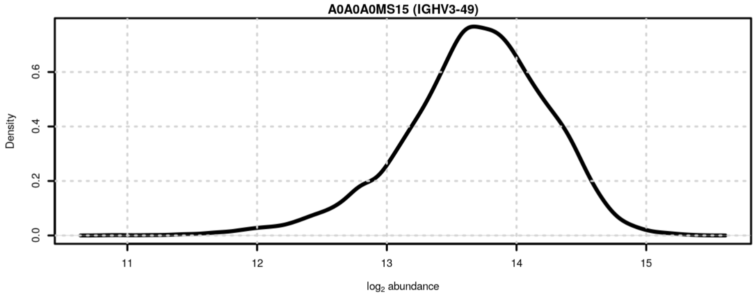

**Figure S5**: Signal distribution for A0A0A0MS15 (IGHV3-49).

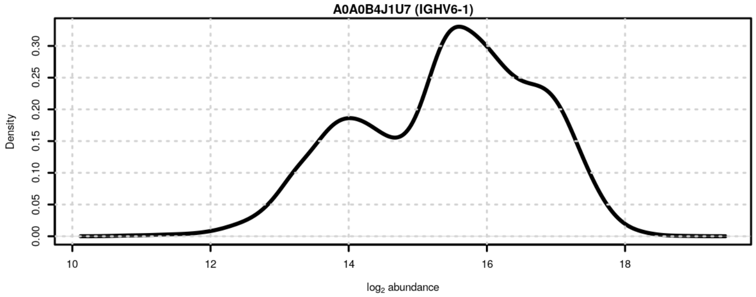

**Figure S6**: Signal distribution for A0A0B4J1U7 (IGHV6-1).

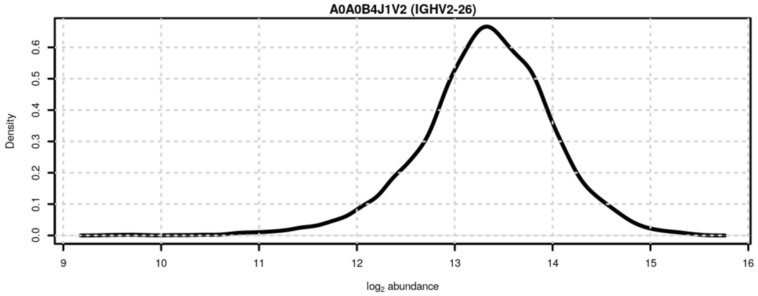

**Figure S7**: Signal distribution for A0A0B4J1V2 (IGHV2-26).

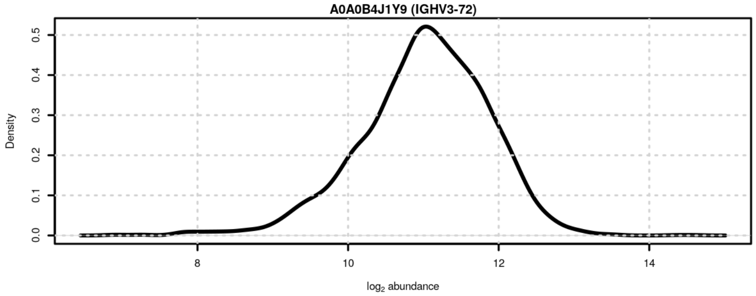

**Figure S8**: Signal distribution for A0A0B4J1Y9 (IGHV3-72).

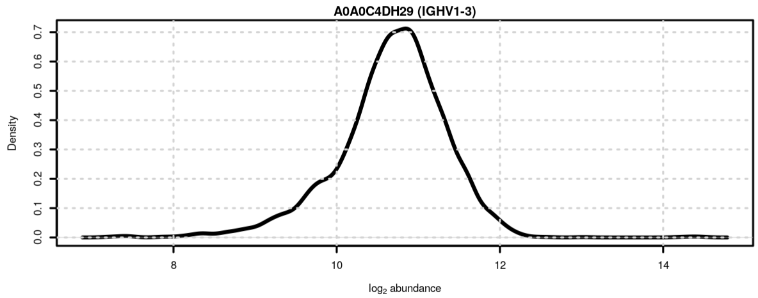

**Figure S9**: Signal distribution for A0A0C4DH29 (IGHV1-3).

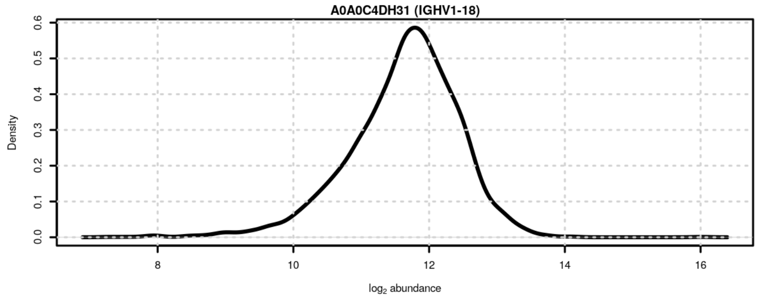

**Figure S10**: Signal distribution for A0A0C4DH31 (IGHV1-18).

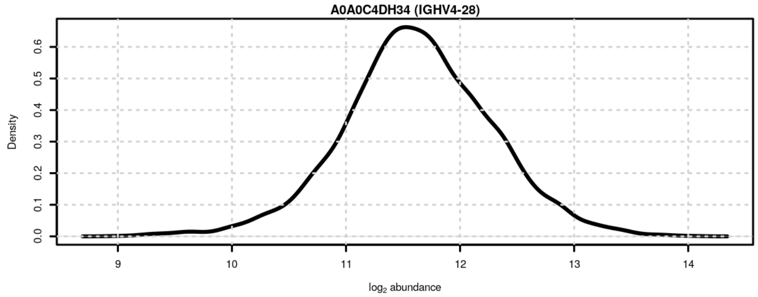

**Figure S11**: Signal distribution for A0A0C4DH34 (IGHV4-28).

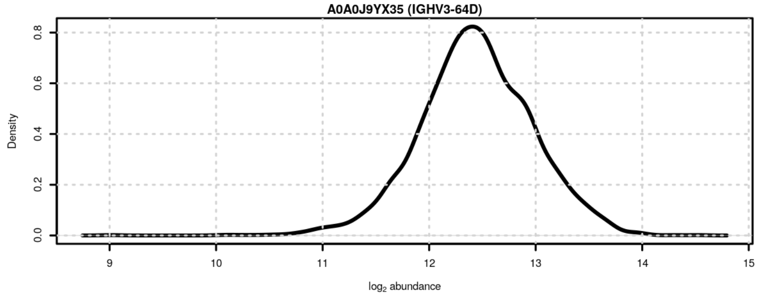

**Figure S12**: Signal distribution for A0A0J9YX35 (IGHV3-64D).

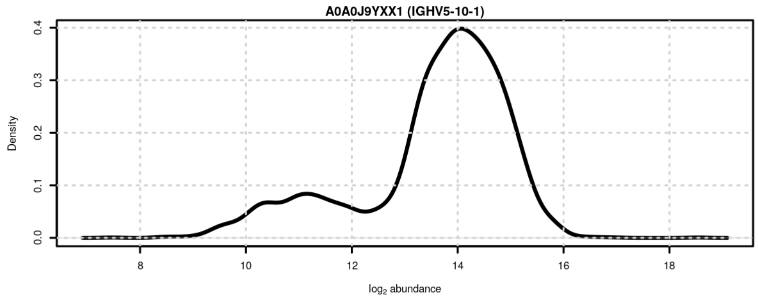

**Figure S13**: Signal distribution for A0A0J9YXX1 (IGHV5-10-1).

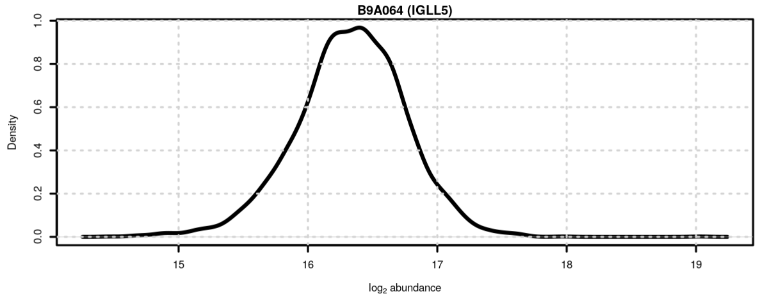

**Figure S14**: Signal distribution for B9A064 (IGLL5).

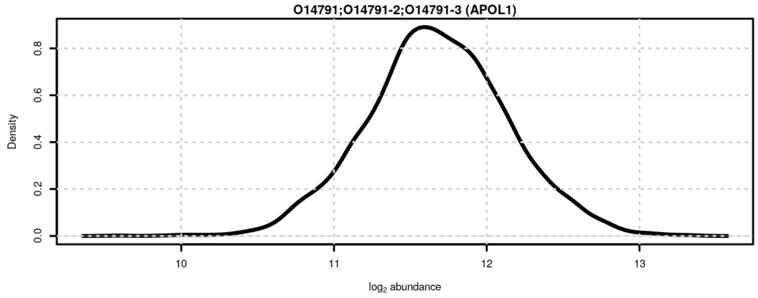

**Figure S15**: Signal distribution for O14791;O14791-2;O14791-3 (APOL1).

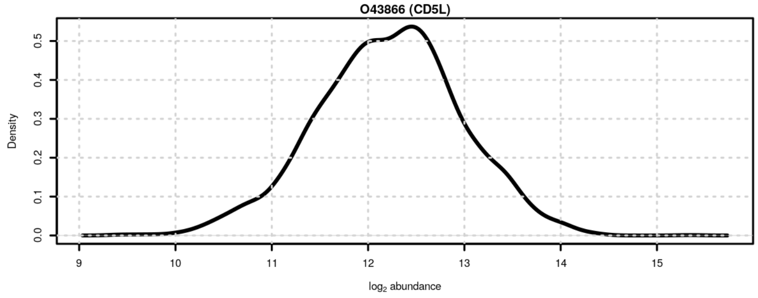

**Figure S16**: Signal distribution for O43866 (CD5L).

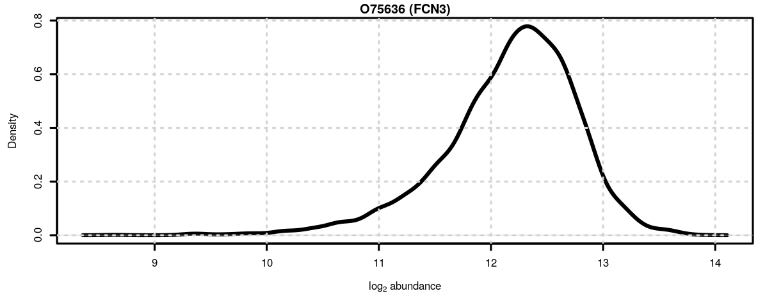

**Figure S17**: Signal distribution for O75636 (FCN3).

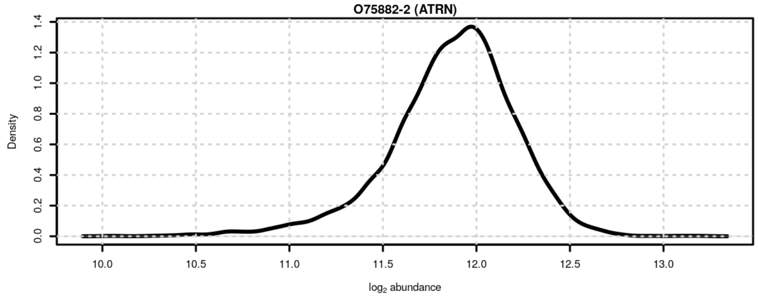

**Figure S18**: Signal distribution for O75882-2 (ATRN).

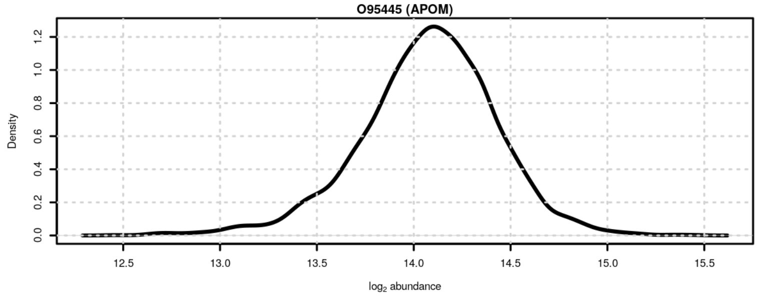

**Figure S19**: Signal distribution for O95445 (APOM).

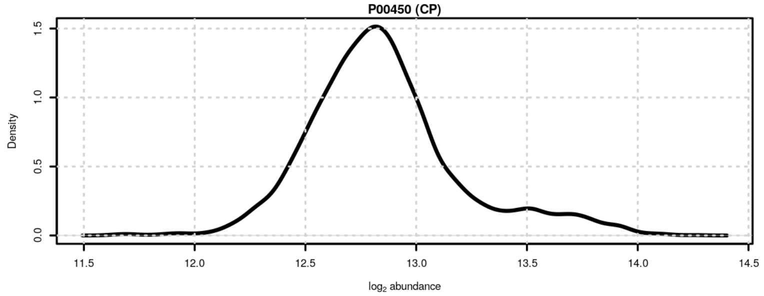

**Figure S20**: Signal distribution for P00450 (CP).

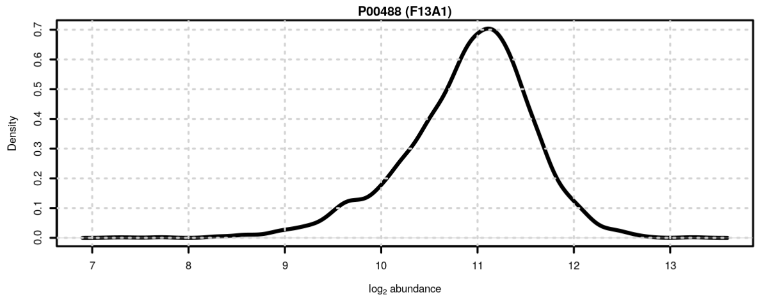

**Figure S21**: Signal distribution for P00488 (F13A1).

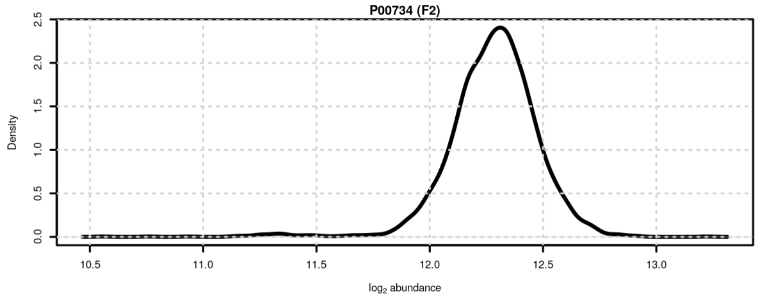

**Figure S22**: Signal distribution for P00734 (F2).

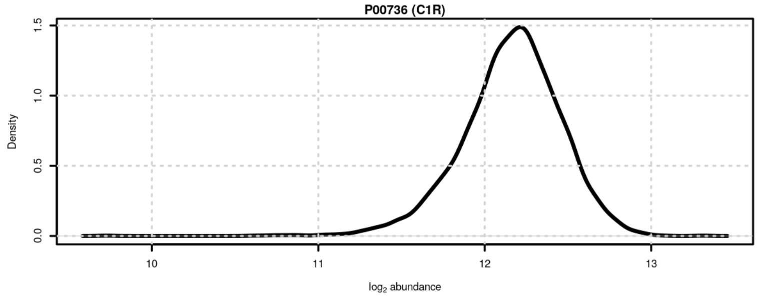

**Figure S23**: Signal distribution for P00736 (C1R).

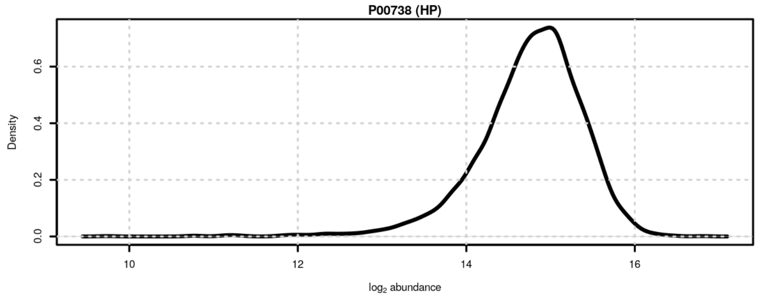

**Figure S24**: Signal distribution for P00738 (HP).

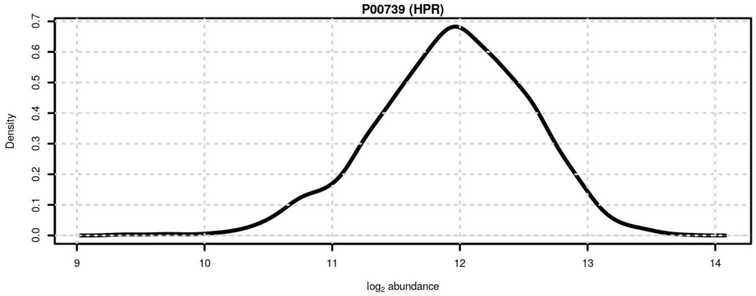

**Figure S25**: Signal distribution for P00739 (HPR).

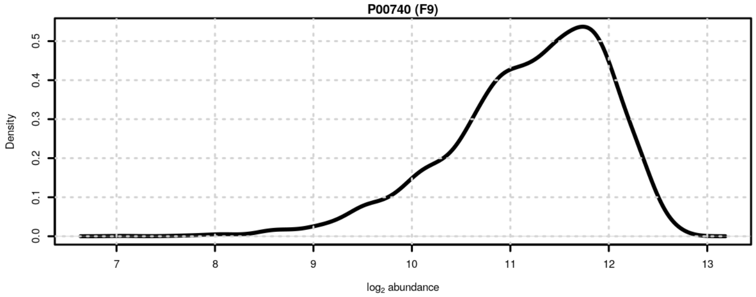

**Figure S26**: Signal distribution for P00740 (F9).

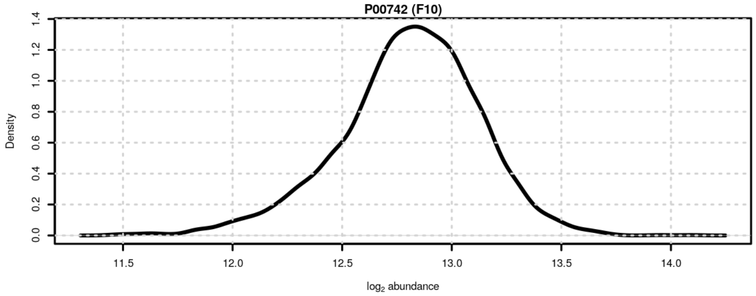

**Figure S27**: Signal distribution for P00742 (F10).

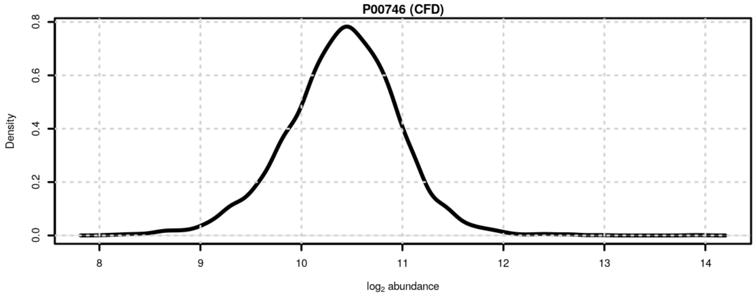

**Figure S28**: Signal distribution for P00746 (CFD).

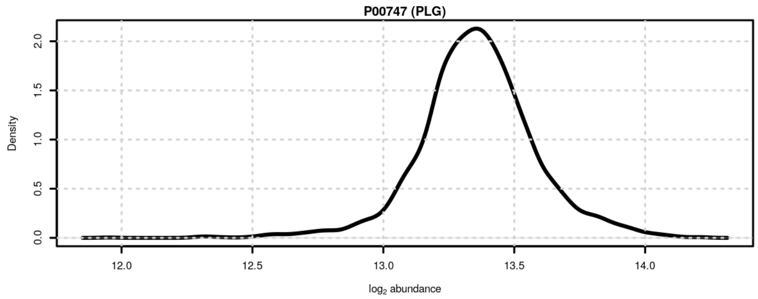

**Figure S29**: Signal distribution for P00747 (PLG).

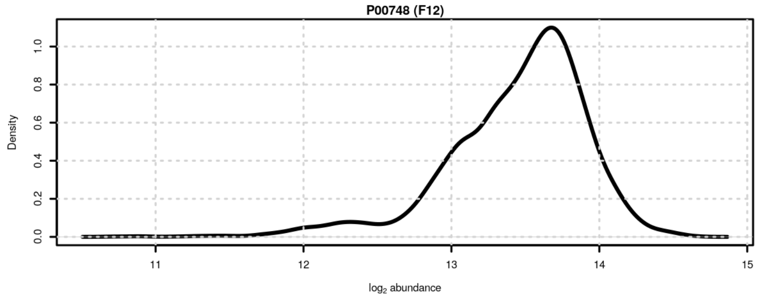

**Figure S30**: Signal distribution for P00748 (F12).

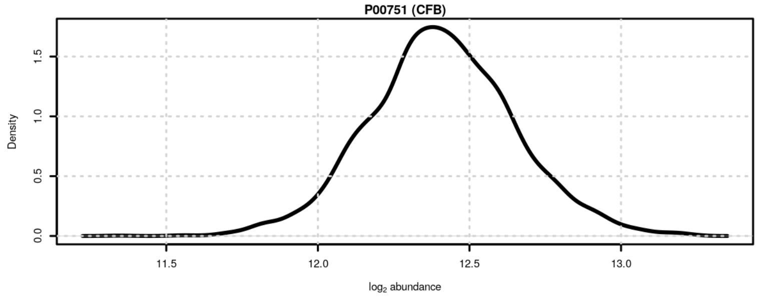

**Figure S31**: Signal distribution for P00751 (CFB).

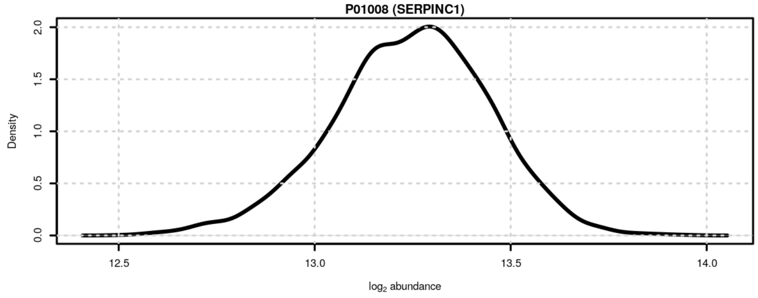

**Figure S32**: Signal distribution for P01008 (SERPINC1).

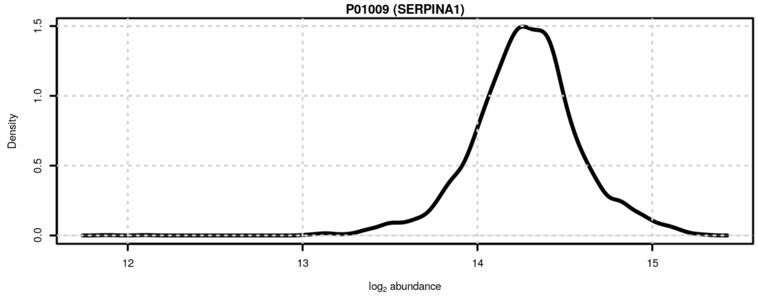

**Figure S33**: Signal distribution for P01009 (SERPINA1).

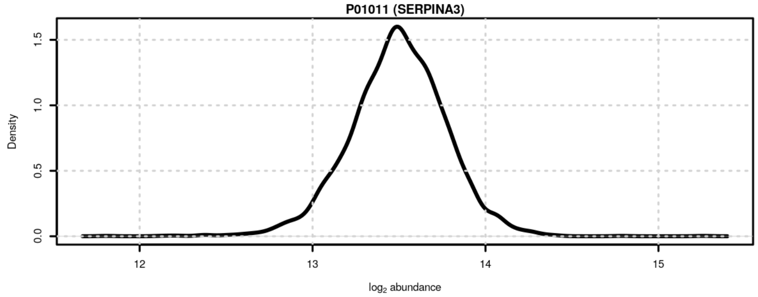

**Figure S34**: Signal distribution for P01011 (SERPINA3).

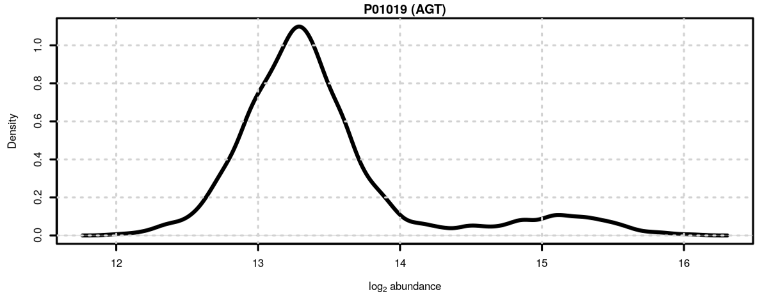

**Figure S35**: Signal distribution for P01019 (AGT).

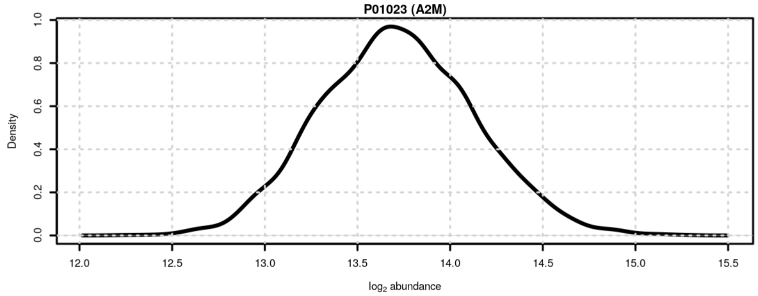

**Figure S36**: Signal distribution for P01023 (A2M).

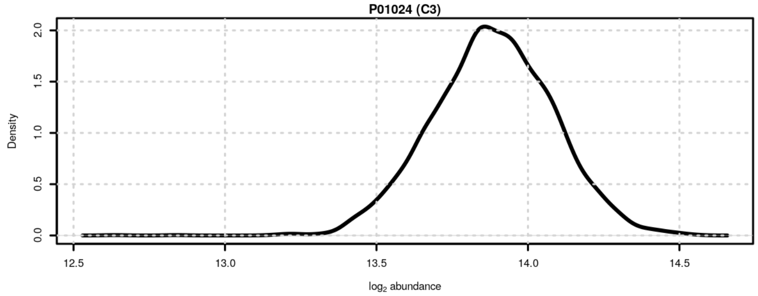

**Figure S37**: Signal distribution for P01024 (C3).

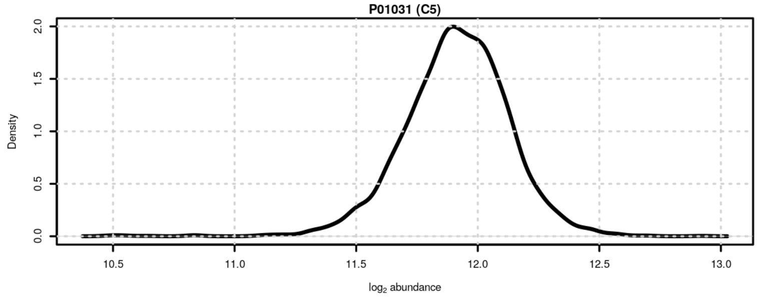

**Figure S38**: Signal distribution for P01031 (C5).

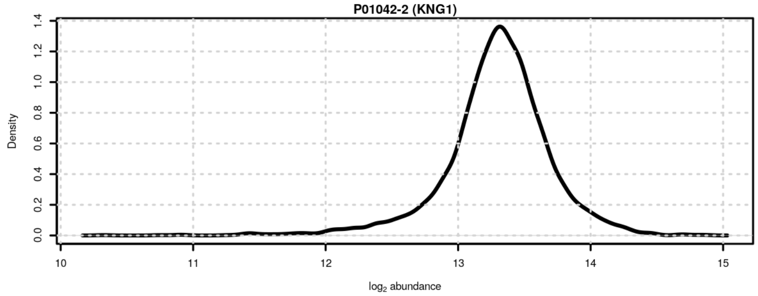

**Figure S39**: Signal distribution for P01042-2 (KNG1).

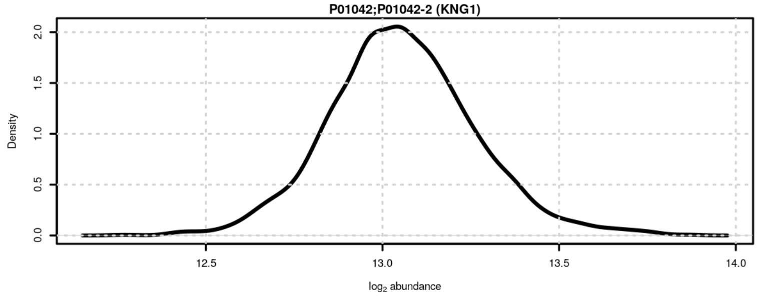

**Figure S40**: Signal distribution for P01042;P01042-2 (KNG1).

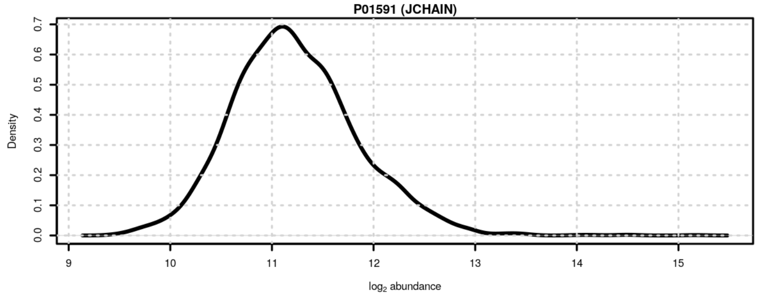

**Figure S41**: Signal distribution for P01591 (JCHAIN).

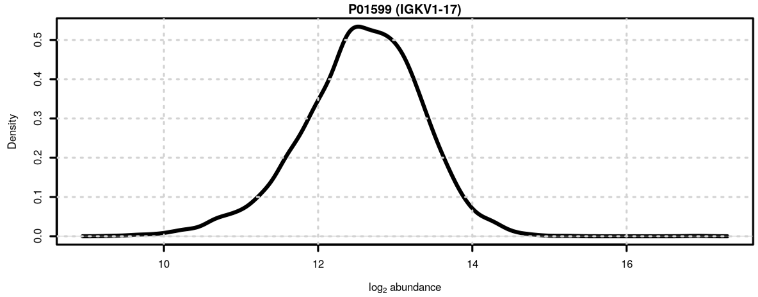

**Figure S42**: Signal distribution for P01599 (IGKV1-17).

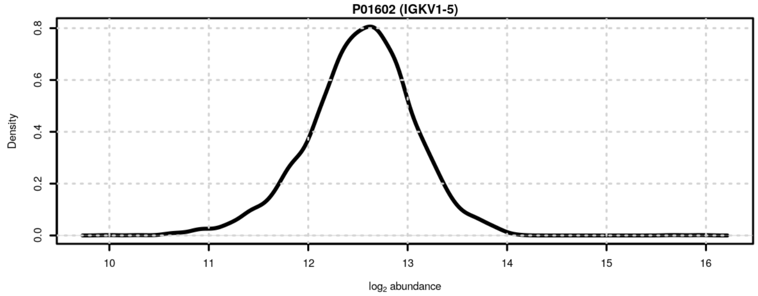

**Figure S43**: Signal distribution for P01602 (IGKV1-5).

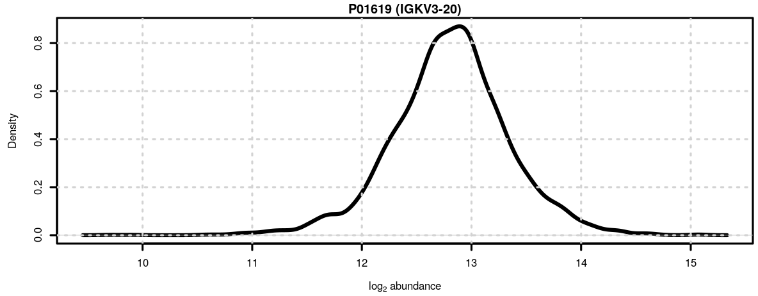

**Figure S44**: Signal distribution for P01619 (IGKV3-20).

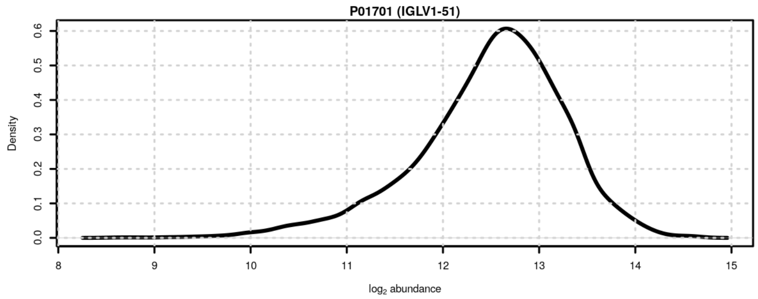

**Figure S45**: Signal distribution for P01701 (IGLV1-51).

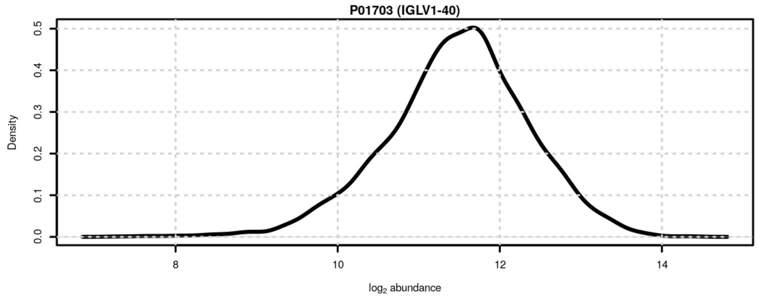

**Figure S46**: Signal distribution for P01703 (IGLV1-40).

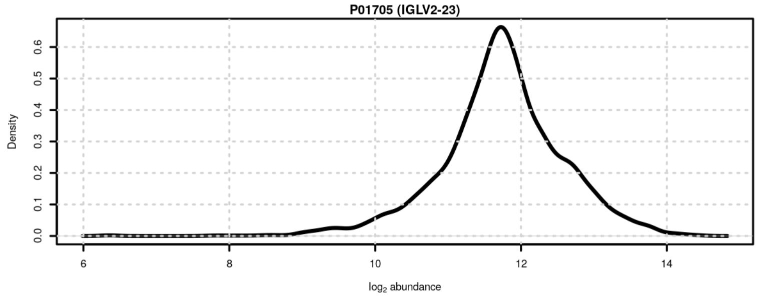

**Figure S47**: Signal distribution for P01705 (IGLV2-23).

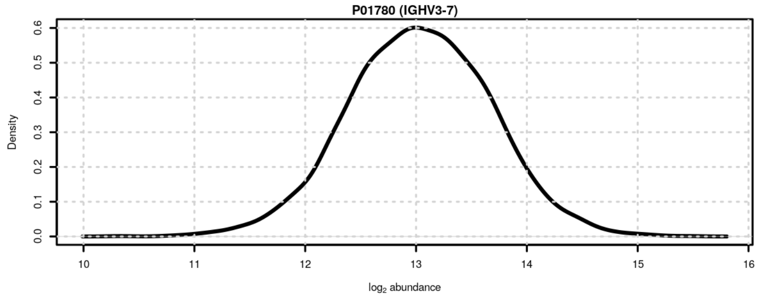

**Figure S48**: Signal distribution for P01780 (IGHV3-7).

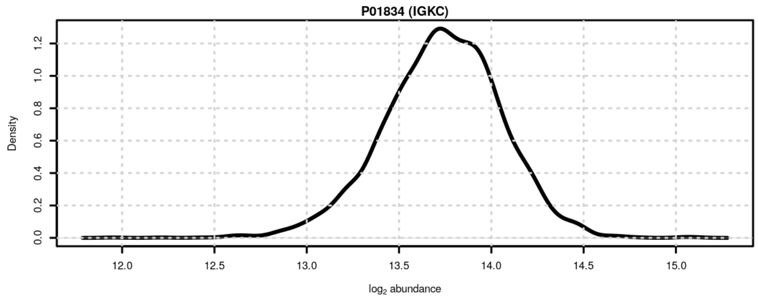

**Figure S49**: Signal distribution for P01834 (IGKC).

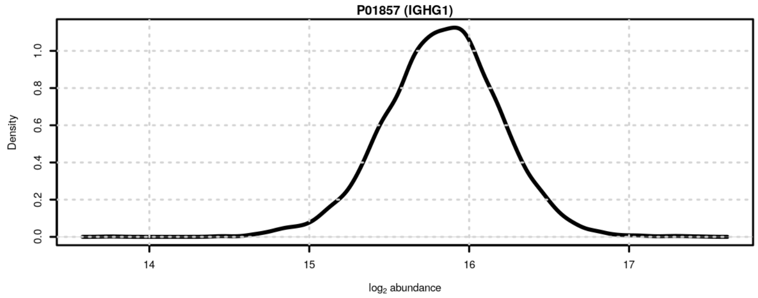

**Figure S50**: Signal distribution for P01857 (IGHG1).

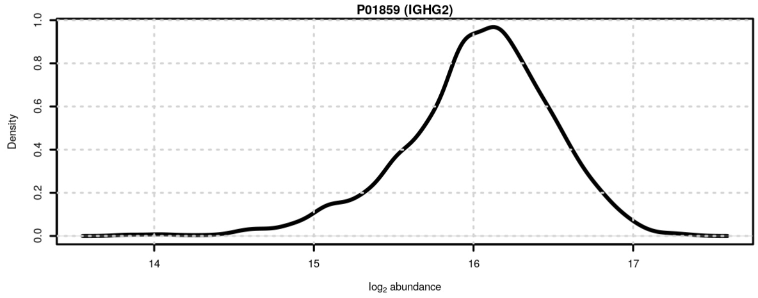

**Figure S51**: Signal distribution for P01859 (IGHG2).

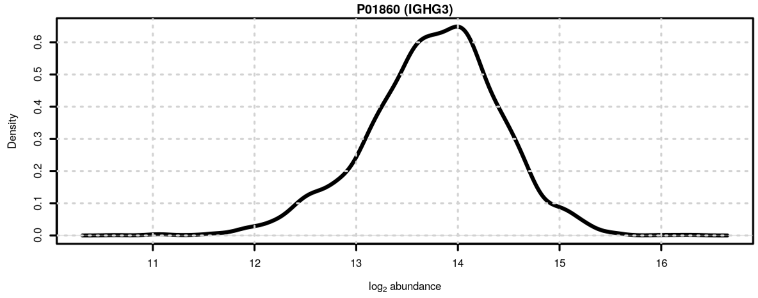

**Figure S52**: Signal distribution for P01860 (IGHG3).

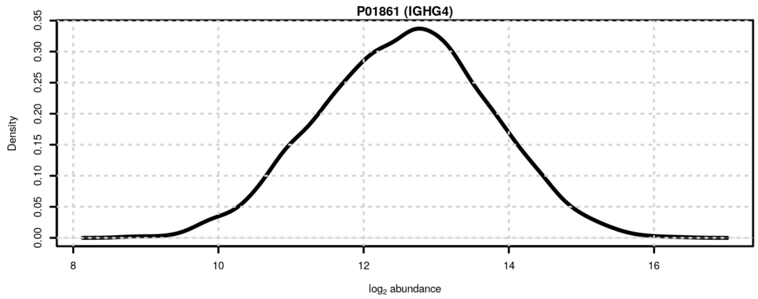

**Figure S53**: Signal distribution for P01861 (IGHG4).

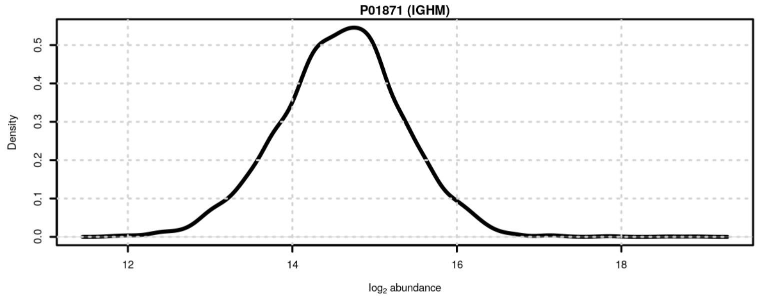

**Figure S54**: Signal distribution for P01871 (IGHM).

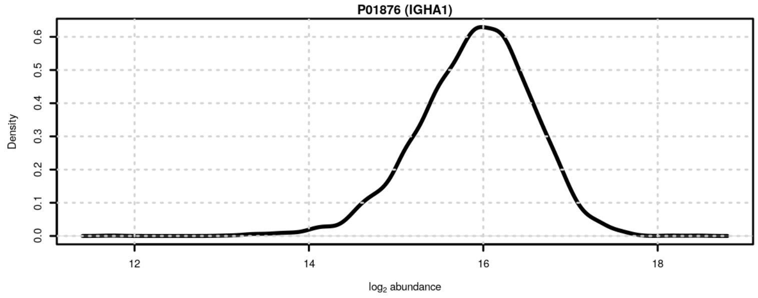

**Figure S55**: Signal distribution for P01876 (IGHA1).

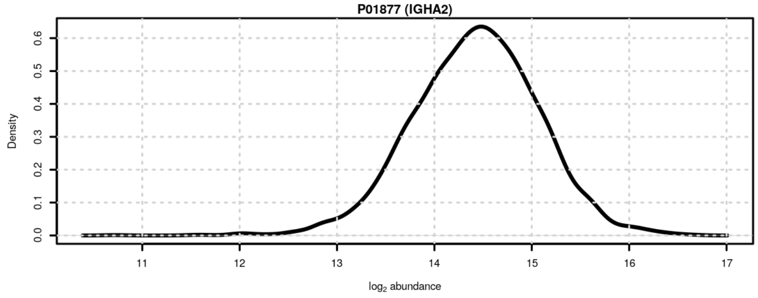

**Figure S56**: Signal distribution for P01877 (IGHA2).

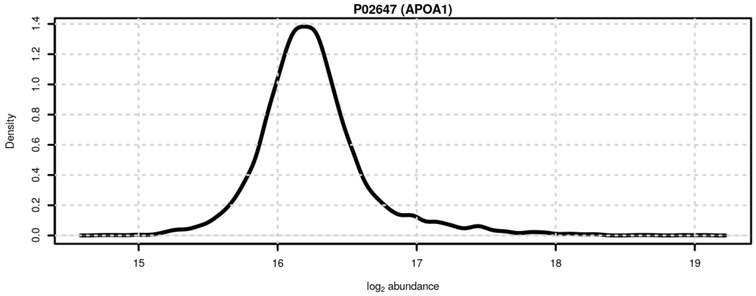

**Figure S57**: Signal distribution for P02647 (APOA1).

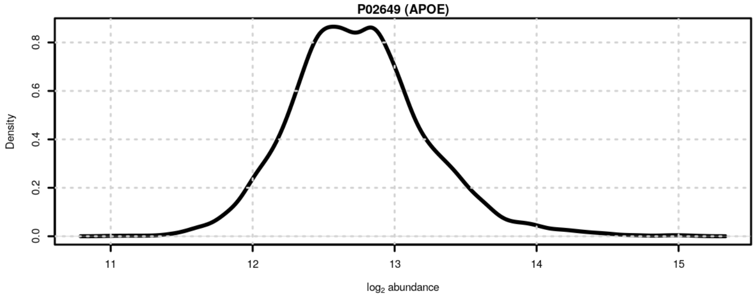

**Figure S58**: Signal distribution for P02649 (APOE).

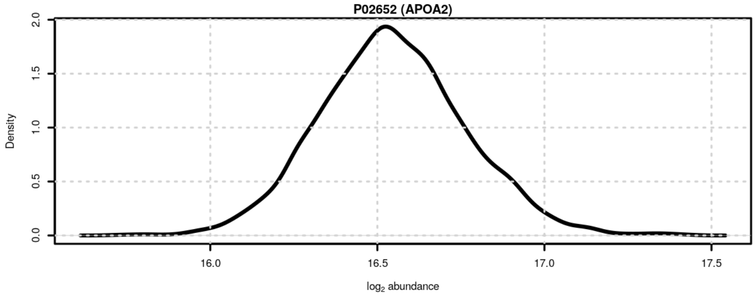

**Figure S59**: Signal distribution for P02652 (APOA2).

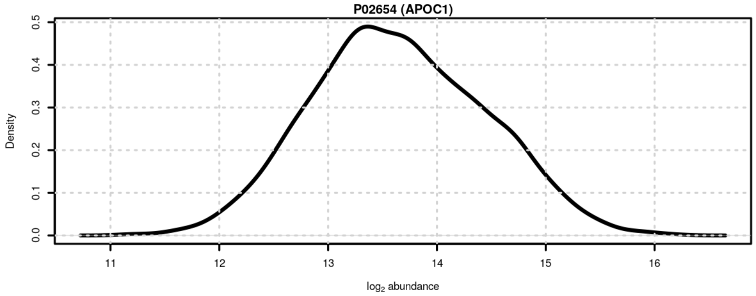

**Figure S60**: Signal distribution for P02654 (APOC1).

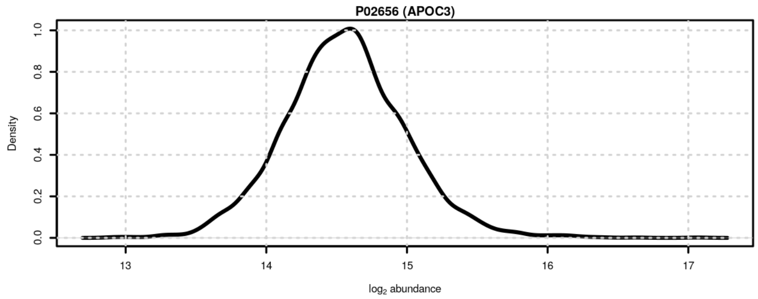

**Figure S61**: Signal distribution for P02656 (APOC3).

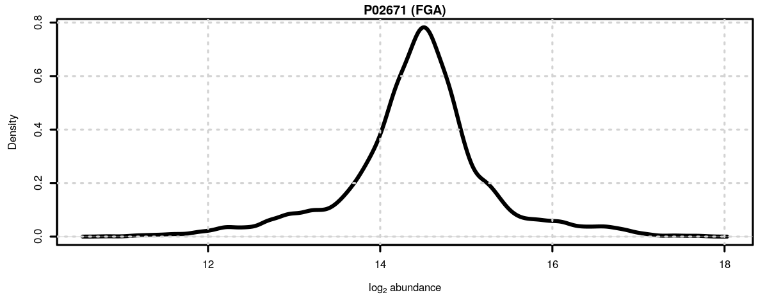

**Figure S62**: Signal distribution for P02671 (FGA).

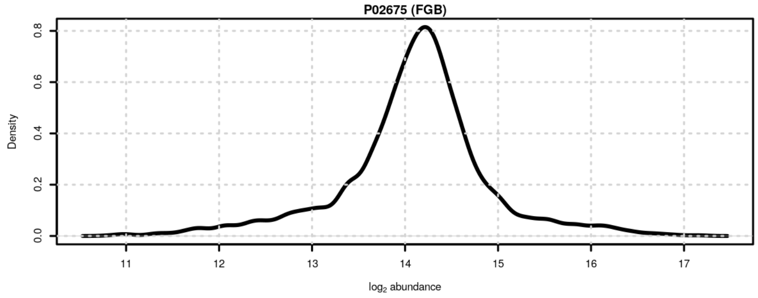

**Figure S63**: Signal distribution for P02675 (FGB).

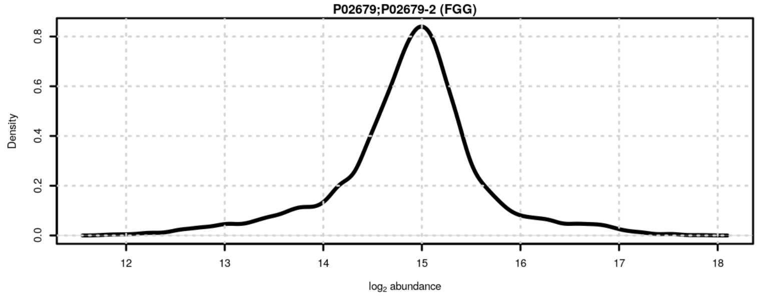

**Figure S64**: Signal distribution for P02679;P02679-2 (FGG).

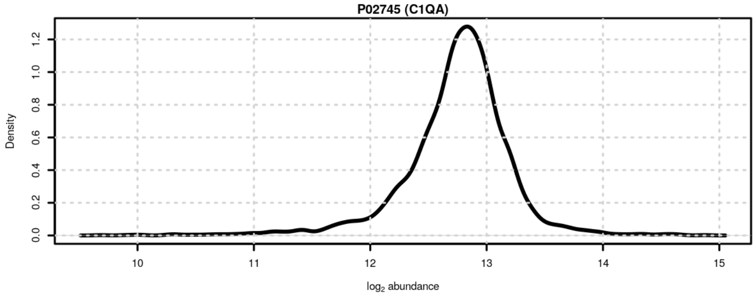

**Figure S65**: Signal distribution for P02745 (C1QA).

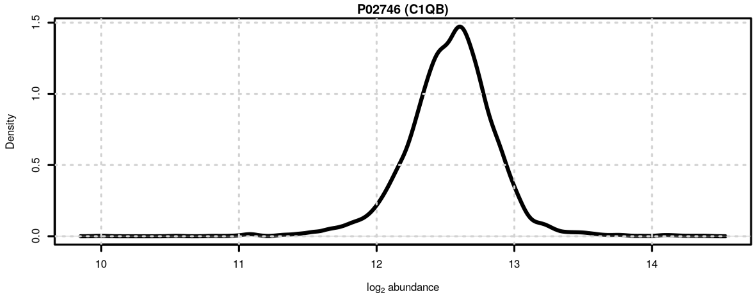

**Figure S66**: Signal distribution for P02746 (C1QB).

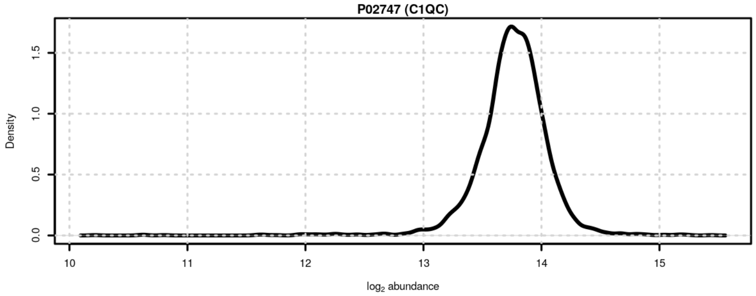

**Figure S67**: Signal distribution for P02747 (C1QC).

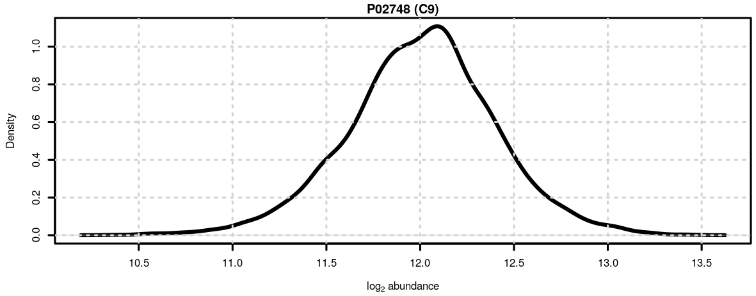

**Figure S68**: Signal distribution for P02748 (C9).

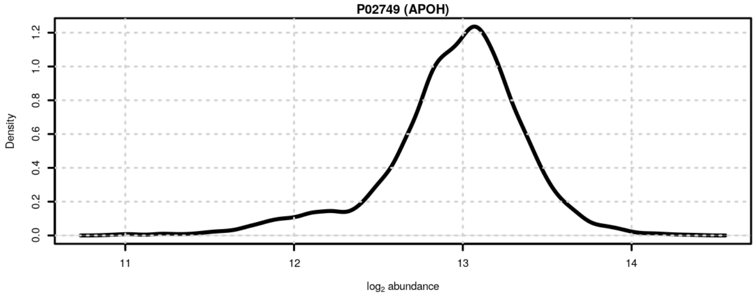

**Figure S69**: Signal distribution for P02749 (APOH).

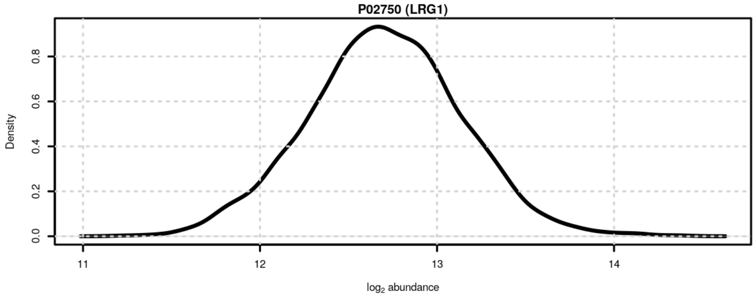

**Figure S70**: Signal distribution for P02750 (LRG1).

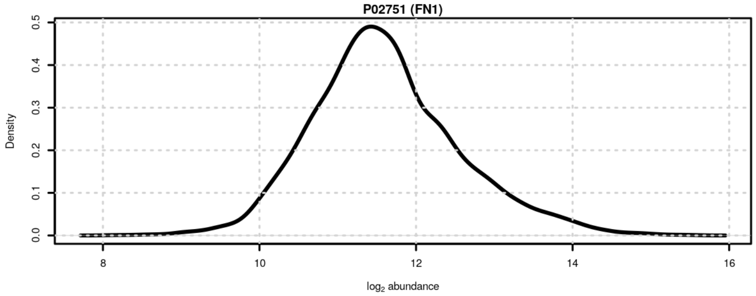

**Figure S71**: Signal distribution for P02751 (FN1).

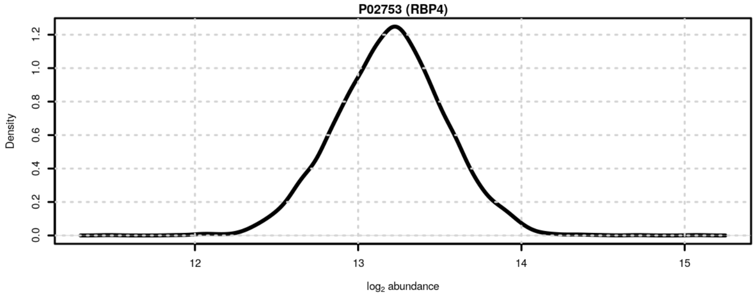

**Figure S72**: Signal distribution for P02753 (RBP4).

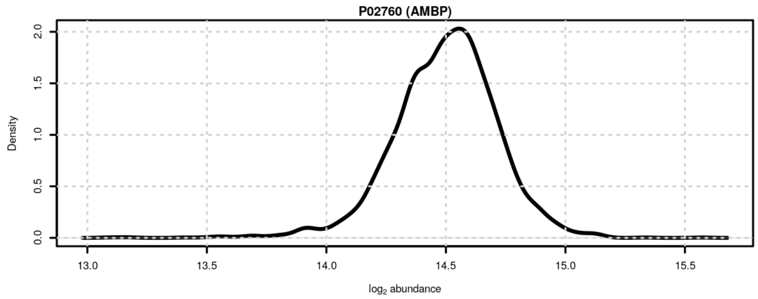

**Figure S73**: Signal distribution for P02760 (AMBP).

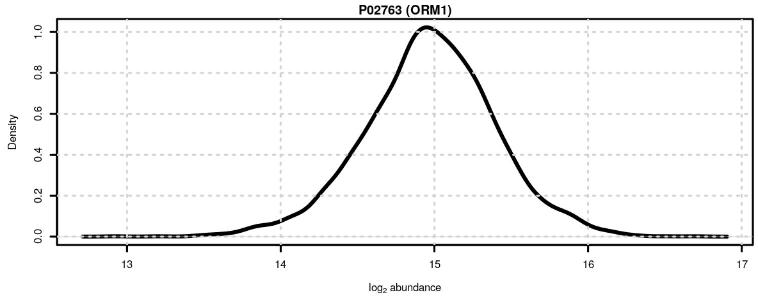

**Figure S74**: Signal distribution for P02763 (ORM1).

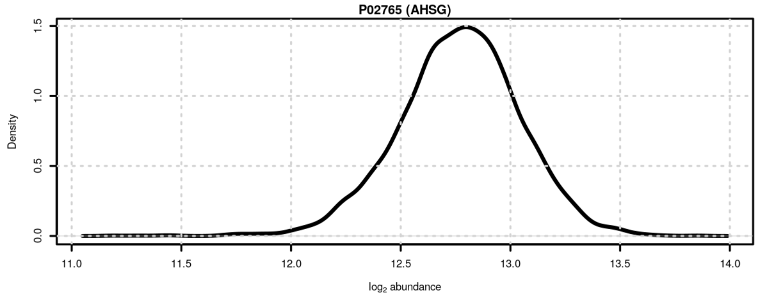

**Figure S75**: Signal distribution for P02765 (AHSG).

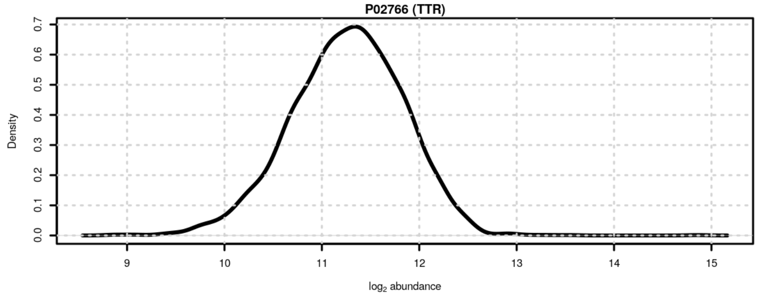

**Figure S76**: Signal distribution for P02766 (TTR).

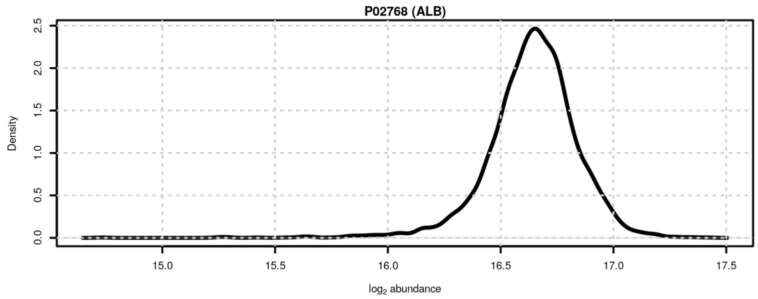

**Figure S77**: Signal distribution for P02768 (ALB).

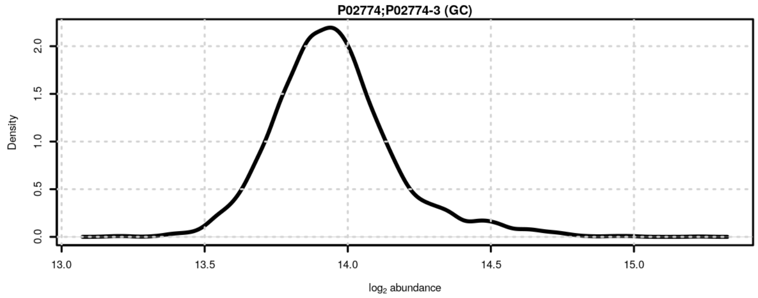

**Figure S78**: Signal distribution for P02774;P02774-3 (GC).

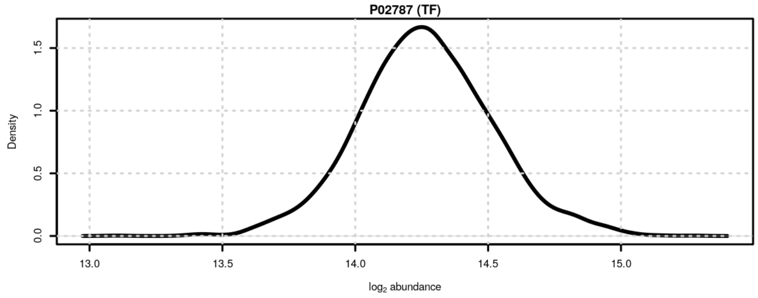

**Figure S79**: Signal distribution for P02787 (TF).

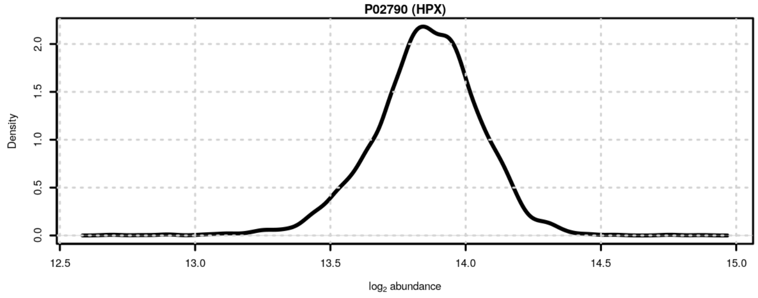

**Figure S80**: Signal distribution for P02790 (HPX).

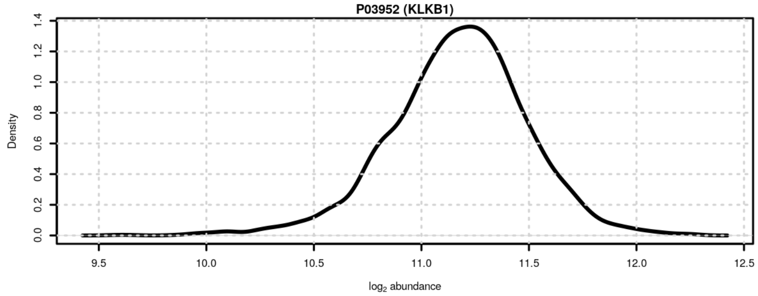

**Figure S81**: Signal distribution for P03952 (KLKB1).

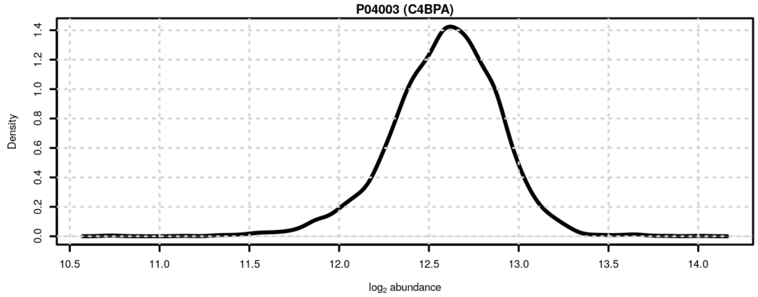

**Figure S82**: Signal distribution for P04003 (C4BPA).

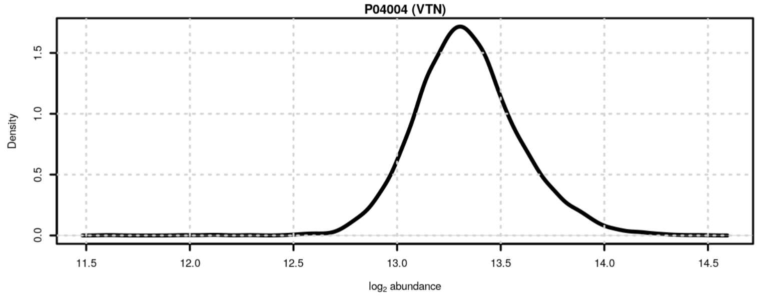

**Figure S83**: Signal distribution for P04004 (VTN).

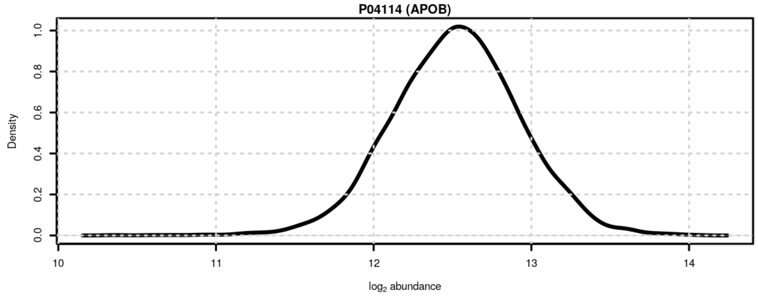

**Figure S84**: Signal distribution for P04114 (APOB).

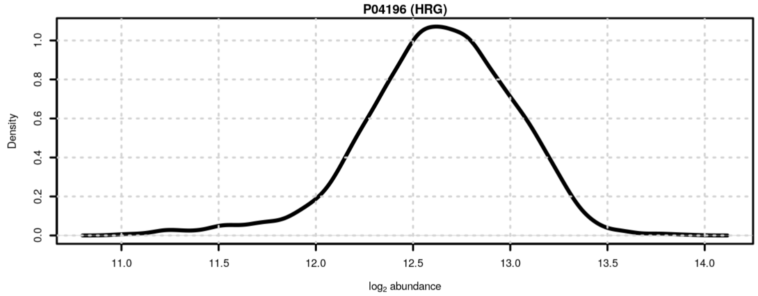

**Figure S85**: Signal distribution for P04196 (HRG).

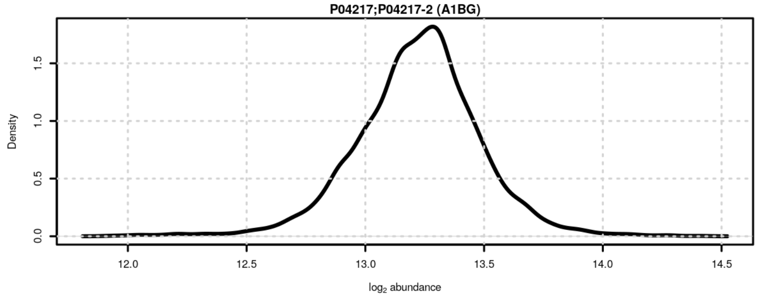

**Figure S86**: Signal distribution for P04217;P04217-2 (A1BG).

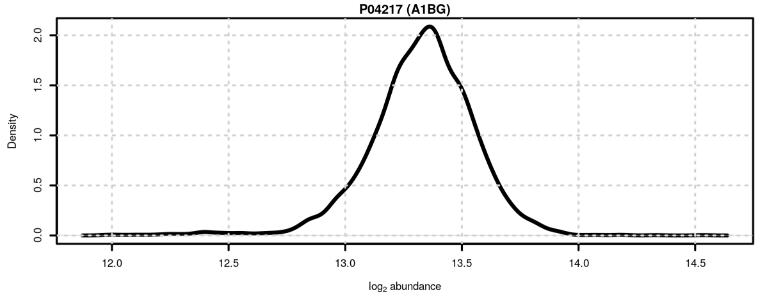

**Figure S87**: Signal distribution for P04217 (A1BG).

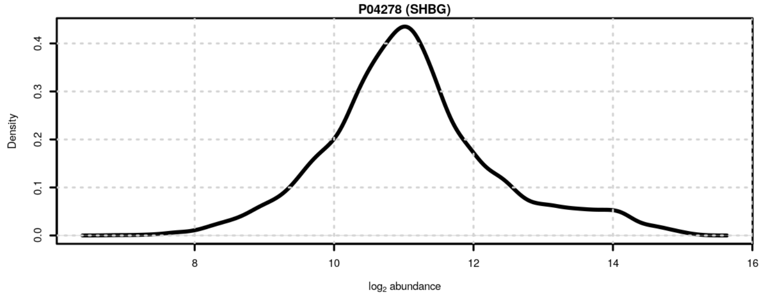

**Figure S88**: Signal distribution for P04278 (SHBG).

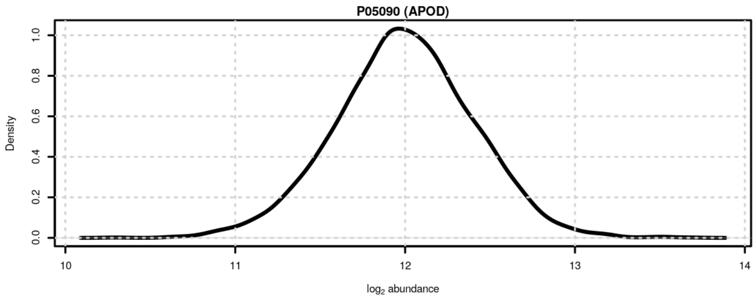

**Figure S89**: Signal distribution for P05090 (APOD).

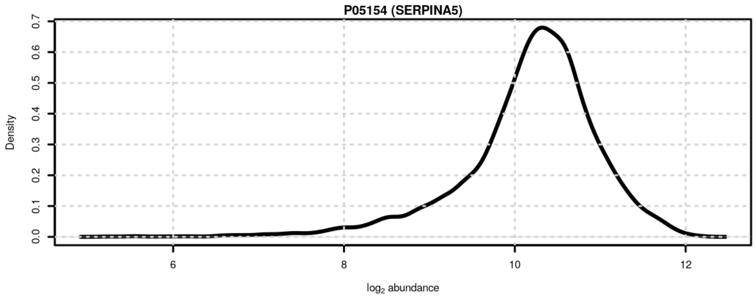

**Figure S90**: Signal distribution for P05154 (SERPINA5).

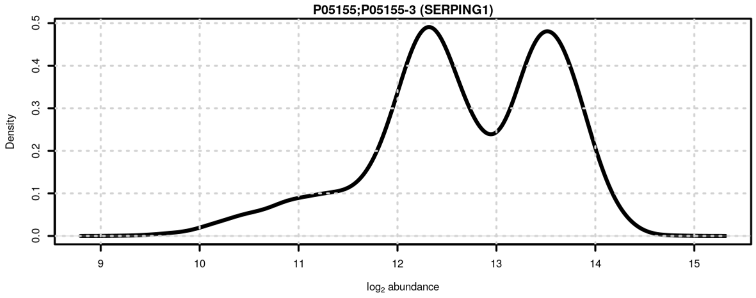

**Figure S91**: Signal distribution for P05155;P05155-3 (SERPING1).

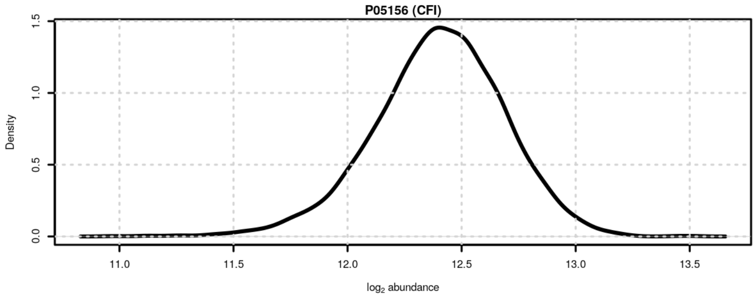

**Figure S92**: Signal distribution for P05156 (CFI).

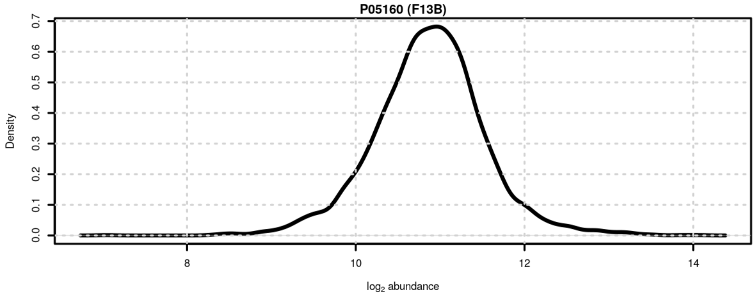

**Figure S93**: Signal distribution for P05160 (F13B).

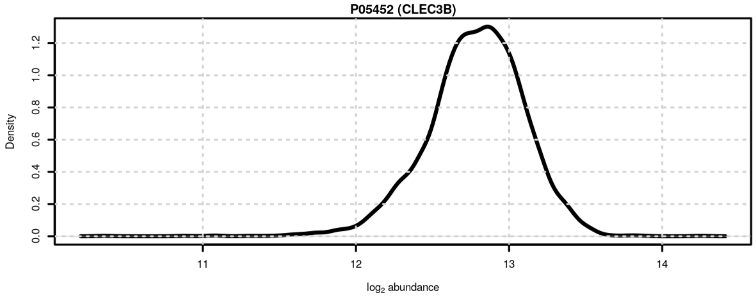

**Figure S94**: Signal distribution for P05452 (CLEC3B).

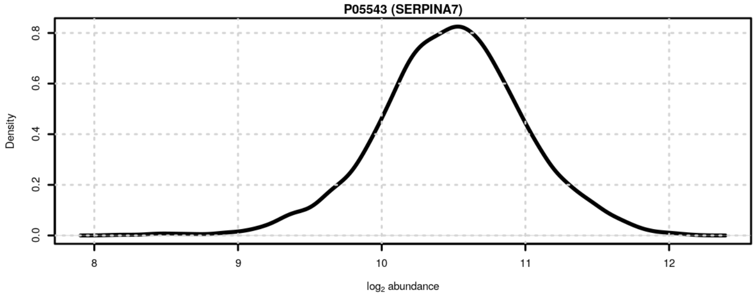

**Figure S95**: Signal distribution for P05543 (SERPINA7).

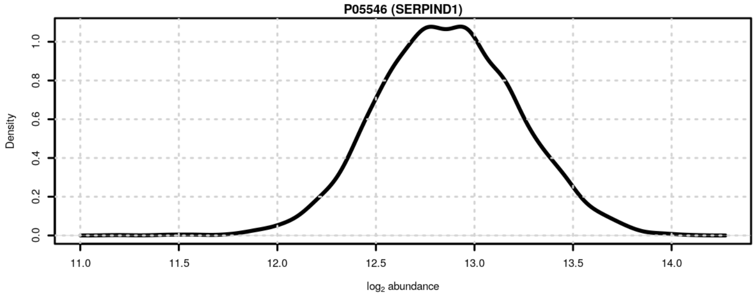

**Figure S96**: Signal distribution for P05546 (SERPIND1).

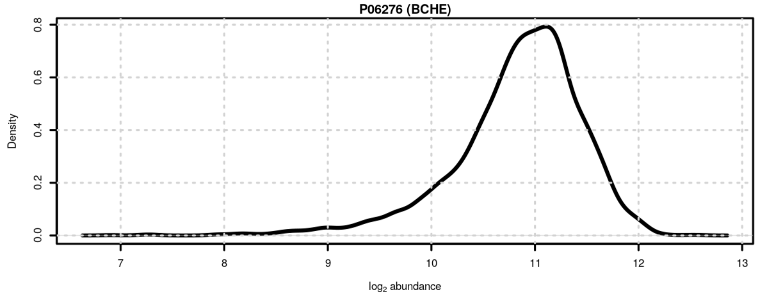

**Figure S97**: Signal distribution for P06276 (BCHE).

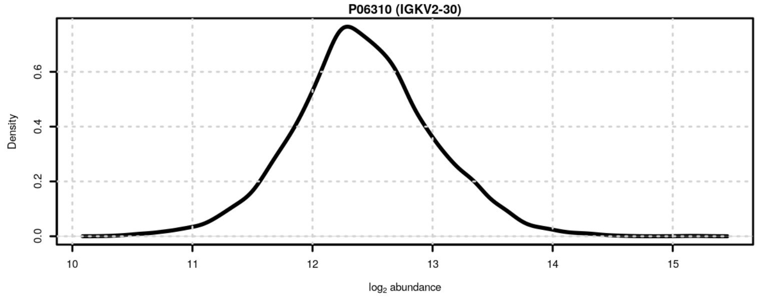

**Figure S98**: Signal distribution for P06310 (IGKV2-30).

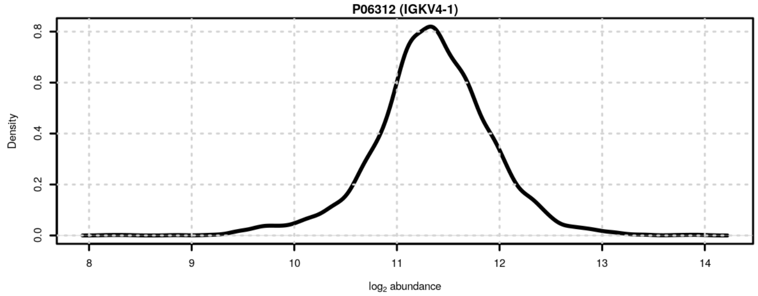

**Figure S99**: Signal distribution for P06312 (IGKV4-1).

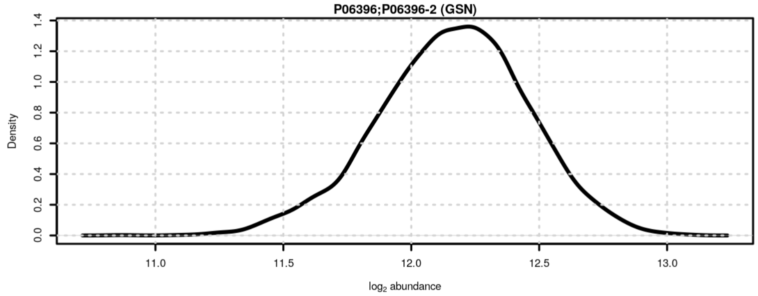

**Figure S100**: Signal distribution for P06396;P06396-2 (GSN).

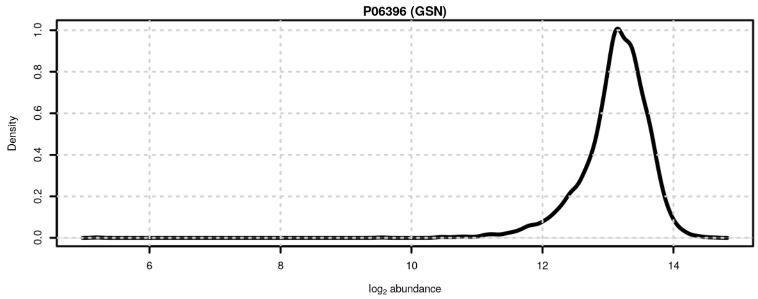

**Figure S101**: Signal distribution for P06396 (GSN).

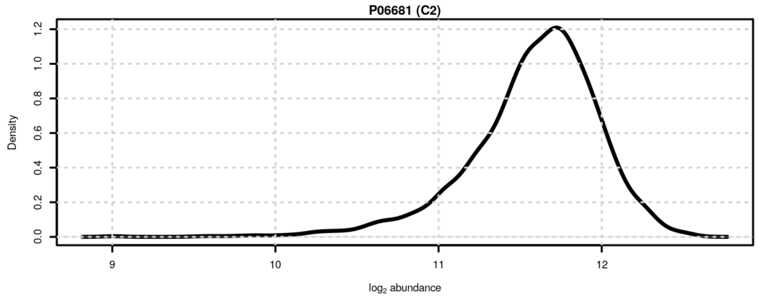

**Figure S102**: Signal distribution for P06681 (C2).

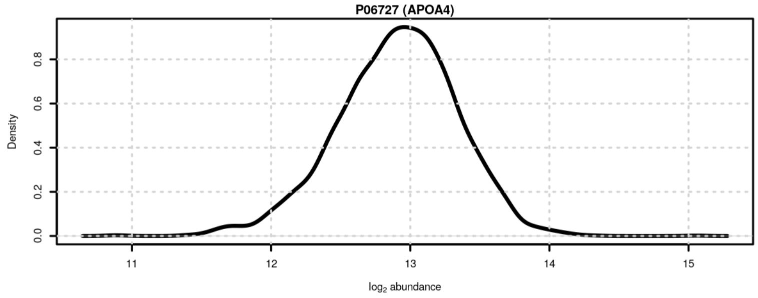

**Figure S103**: Signal distribution for P06727 (APOA4).

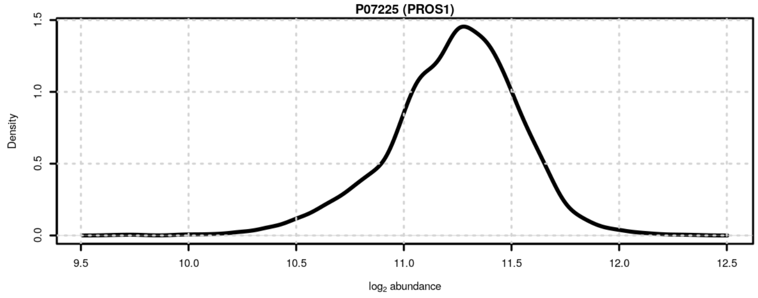

**Figure S104**: Signal distribution for P07225 (PROS1).

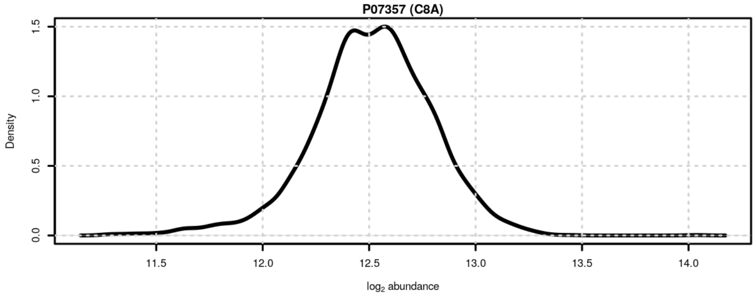

**Figure S105**: Signal distribution for P07357 (C8A).

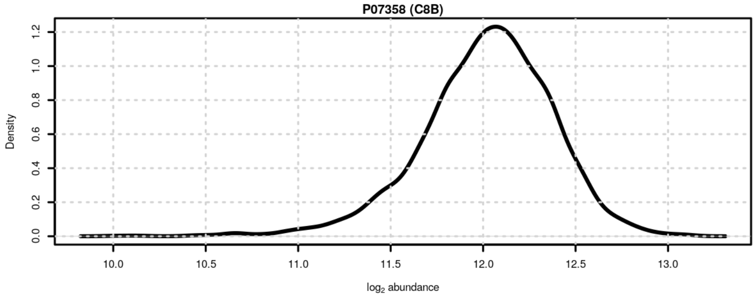

**Figure S106**: Signal distribution for P07358 (C8B).

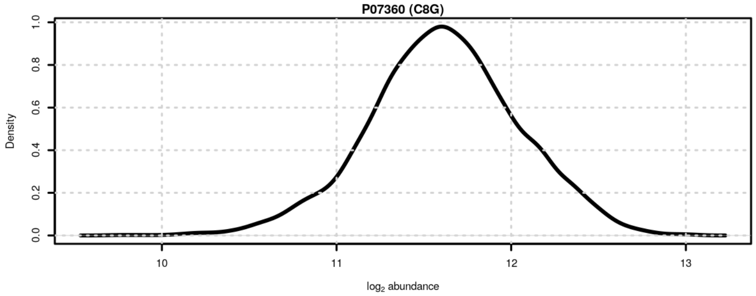

**Figure S107**: Signal distribution for P07360 (C8G).

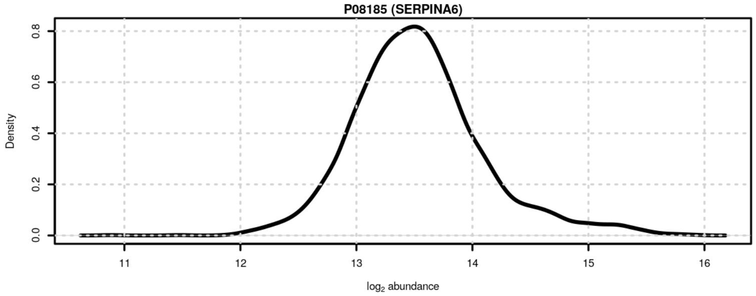

**Figure S108**: Signal distribution for P08185 (SERPINA6).

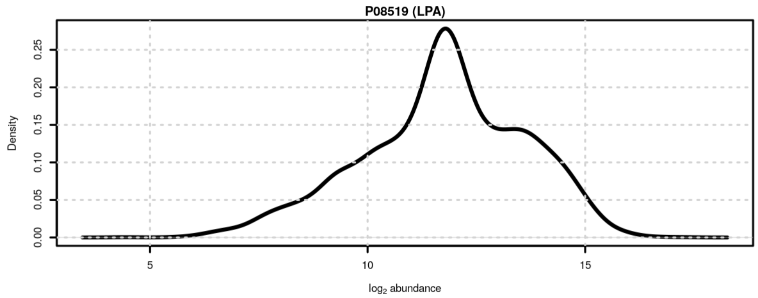

**Figure S109**: Signal distribution for P08519 (LPA).

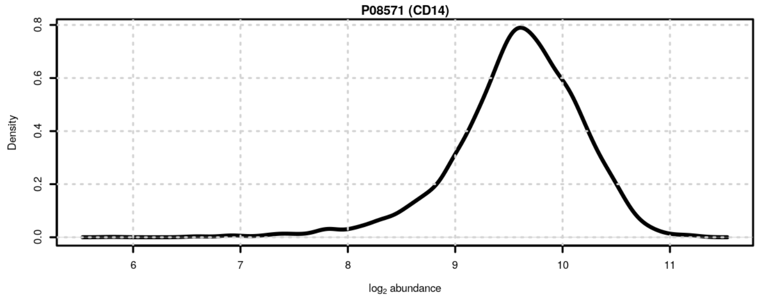

**Figure S110**: Signal distribution for P08571 (CD14).

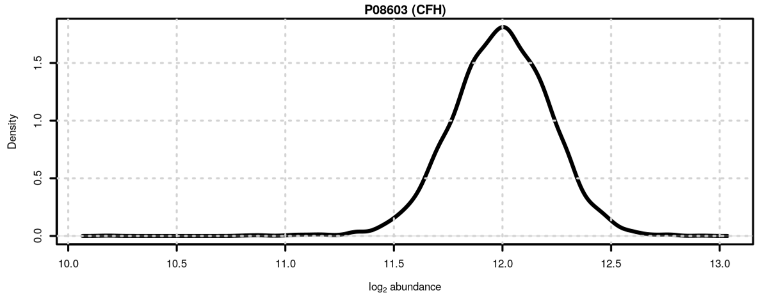

**Figure S111**: Signal distribution for P08603 (CFH).

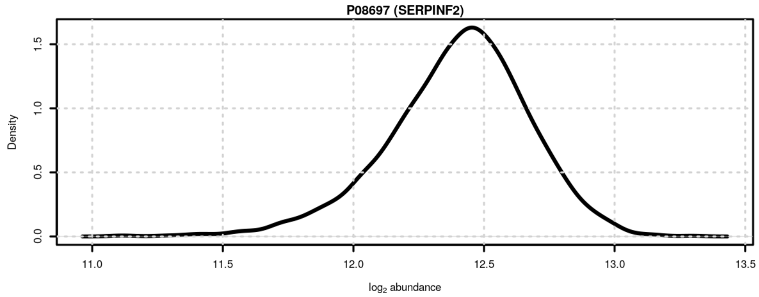

**Figure S112**: Signal distribution for P08697 (SERPINF2).

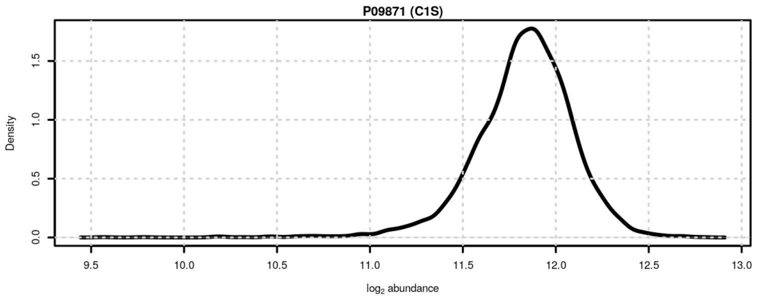

**Figure S113**: Signal distribution for P09871 (C1S).

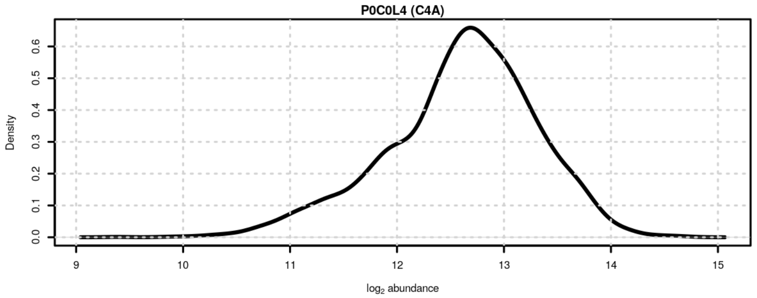

**Figure S114**: Signal distribution for P0C0L4 (C4A).

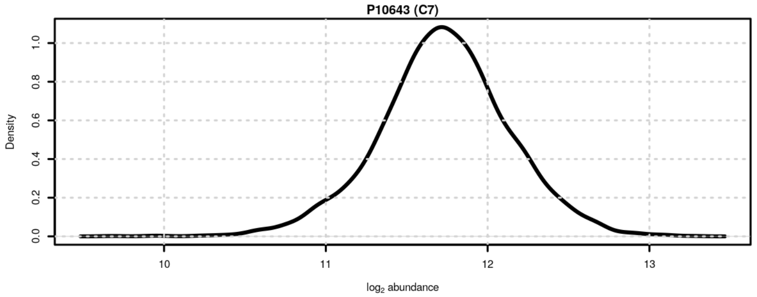

**Figure S115**: Signal distribution for P10643 (C7).

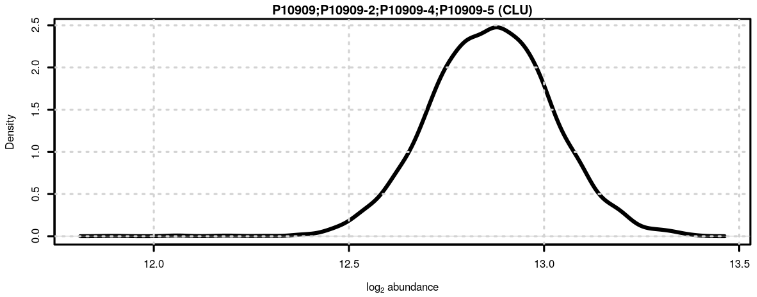

**Figure S116**: Signal distribution for P10909;P10909-2;P10909-4;P10909-5 (CLU).

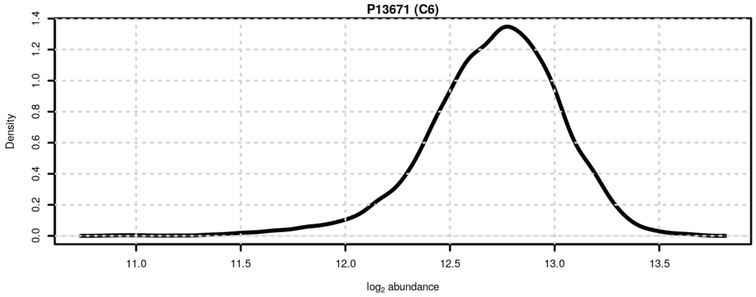

**Figure S117**: Signal distribution for P13671 (C6).

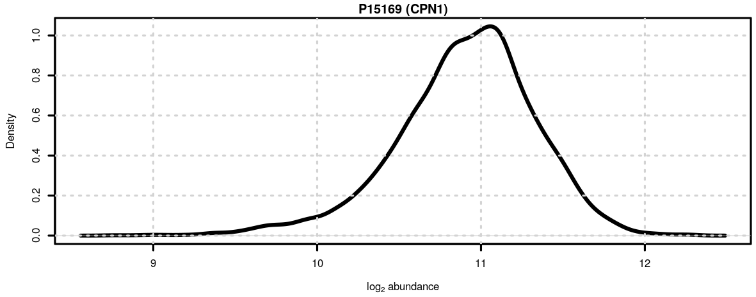

**Figure S118**: Signal distribution for P15169 (CPN1).

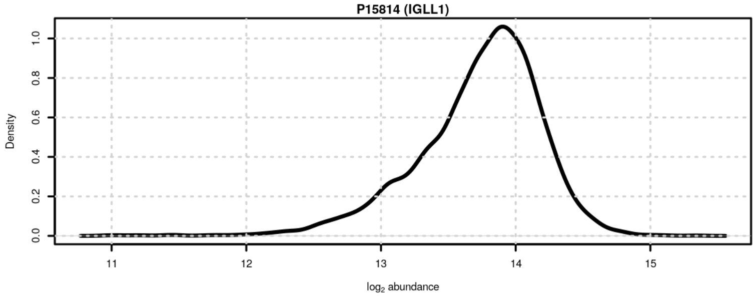

**Figure S119**: Signal distribution for P15814 (IGLL1).

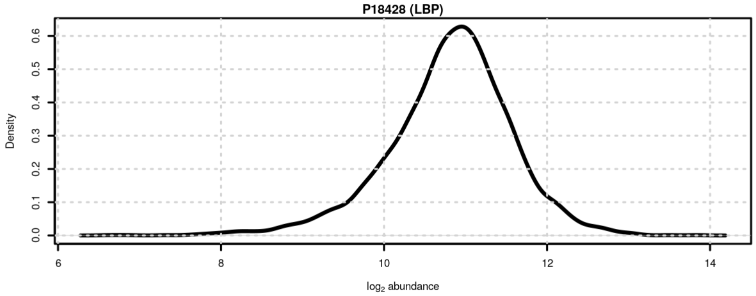

**Figure S120**: Signal distribution for P18428 (LBP).

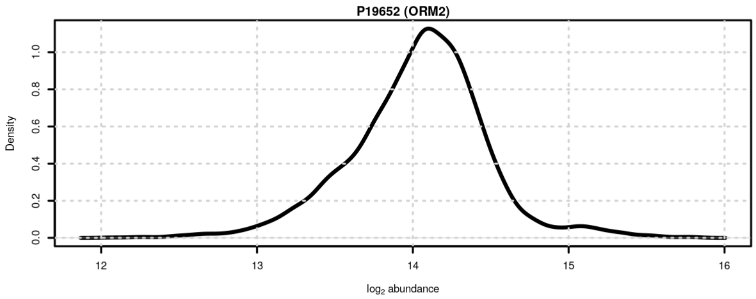

**Figure S121**: Signal distribution for P19652 (ORM2).

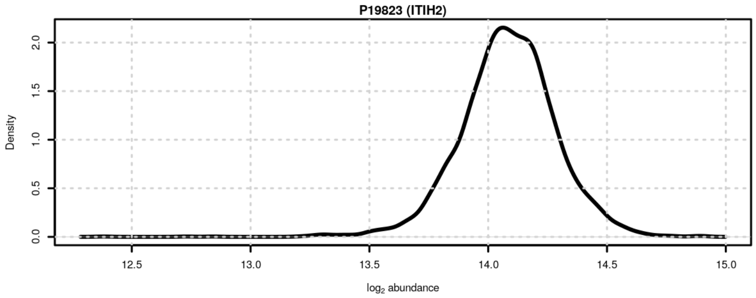

**Figure S122**: Signal distribution for P19823 (ITIH2).

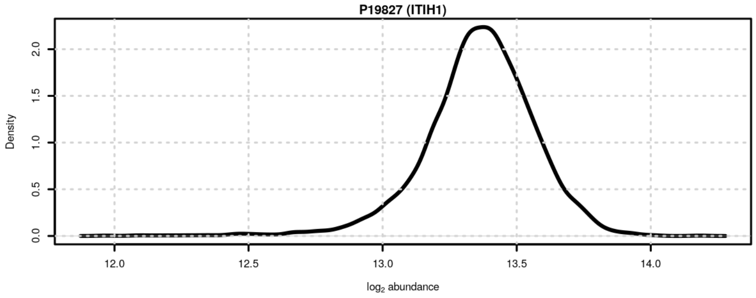

**Figure S123**: Signal distribution for P19827 (ITIH1).

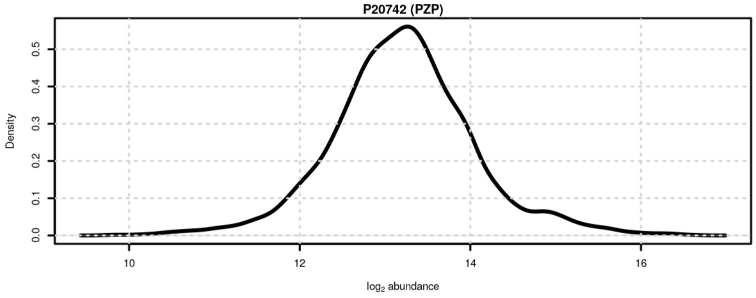

**Figure S124**: Signal distribution for P20742 (PZP).

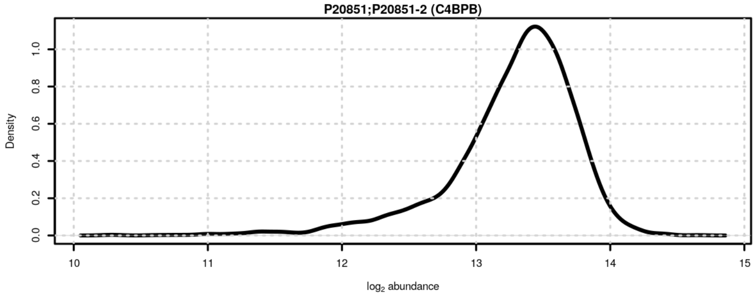

**Figure S125**: Signal distribution for P20851;P20851-2 (C4BPB).

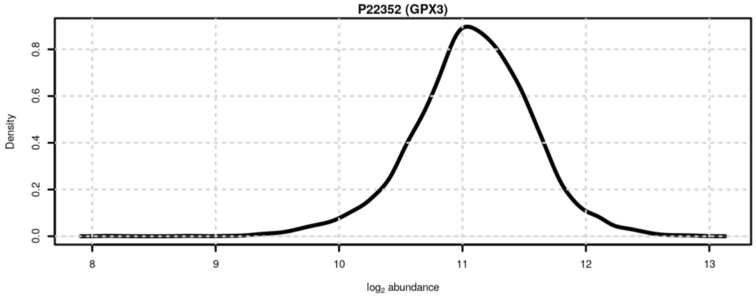

**Figure S126**: Signal distribution for P22352 (GPX3).

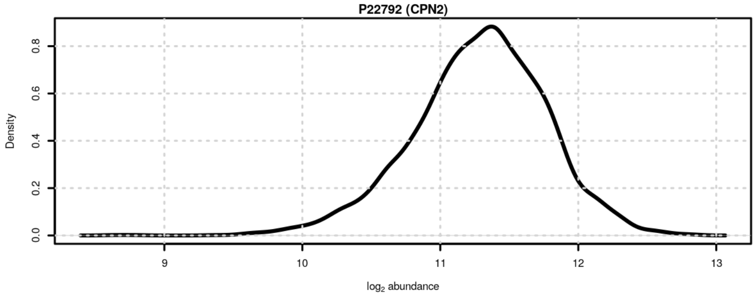

**Figure S127**: Signal distribution for P22792 (CPN2).

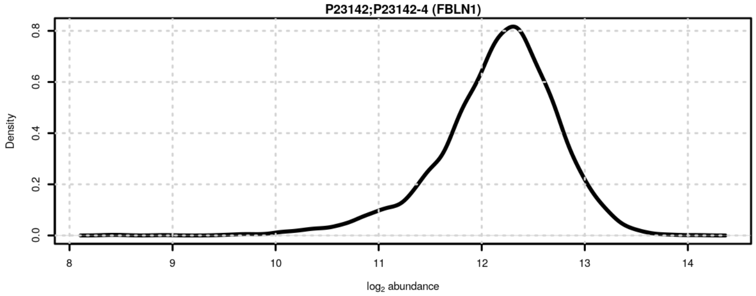

**Figure S128**: Signal distribution for P23142;P23142-4 (FBLN1).

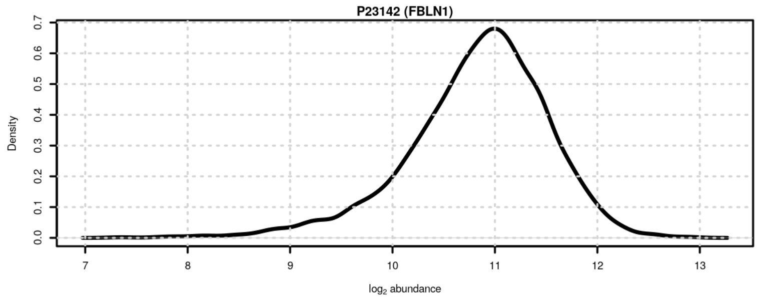

**Figure S129**: Signal distribution for P23142 (FBLN1).

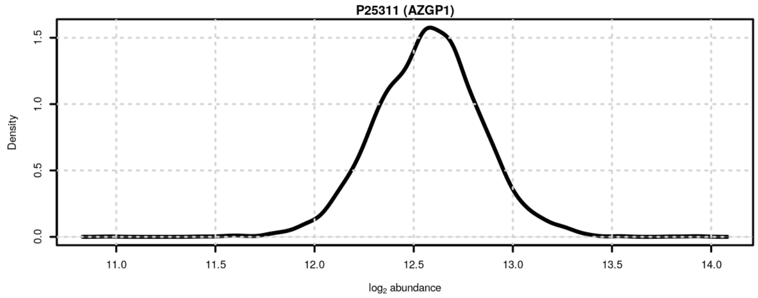

**Figure S130**: Signal distribution for P25311 (AZGP1).

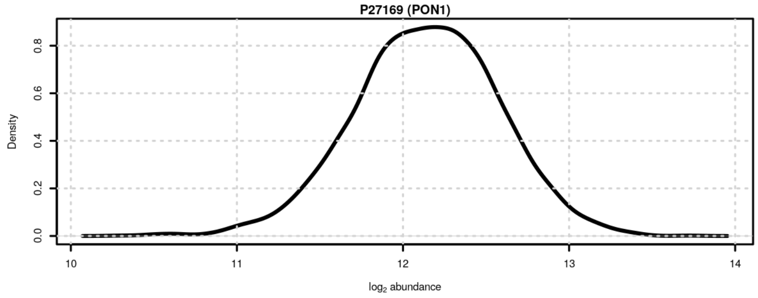

**Figure S131**: Signal distribution for P27169 (PON1).

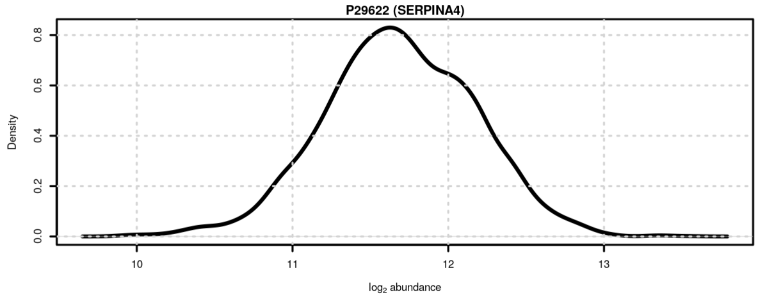

**Figure S132**: Signal distribution for P29622 (SERPINA4).

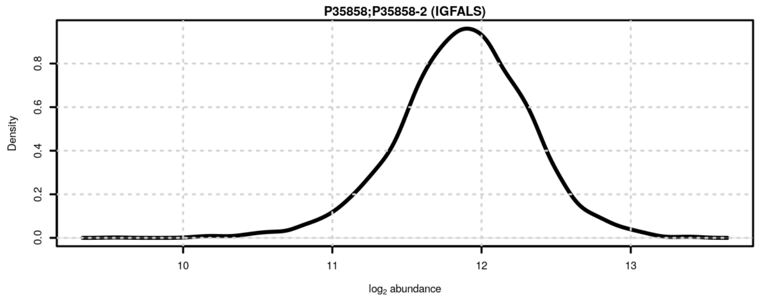

**Figure S133**: Signal distribution for P35858;P35858-2 (IGFALS).

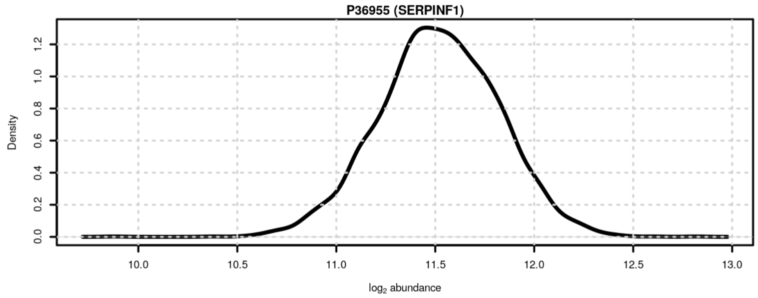

**Figure S134**: Signal distribution for P36955 (SERPINF1).

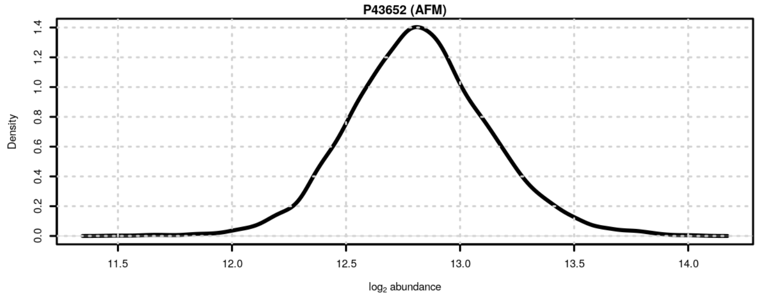

**Figure S135**: Signal distribution for P43652 (AFM).

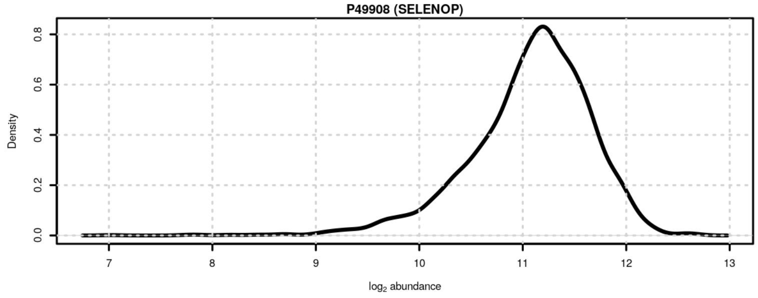

**Figure S136**: Signal distribution for P49908 (SELENOP).

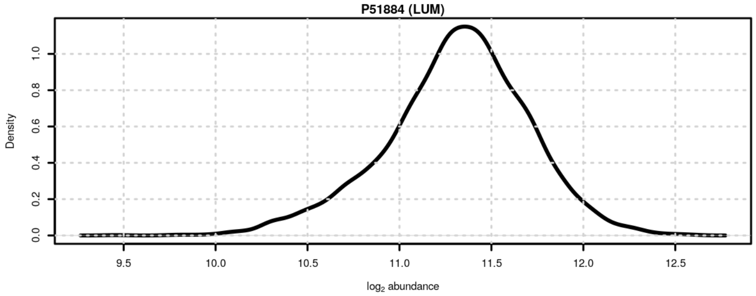

**Figure S137**: Signal distribution for P51884 (LUM).

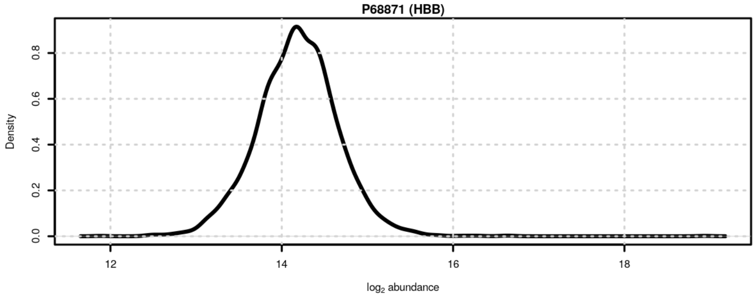

**Figure S138**: Signal distribution for P68871 (HBB).

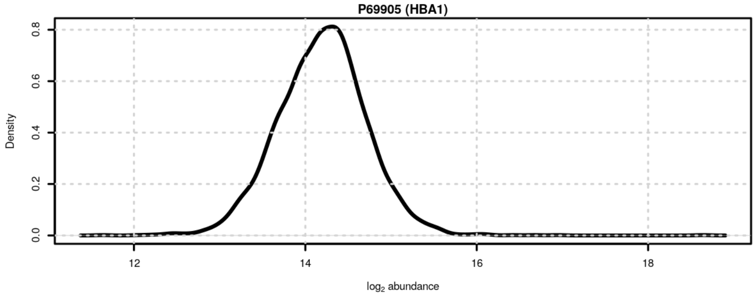

**Figure S139**: Signal distribution for P69905 (HBA1).

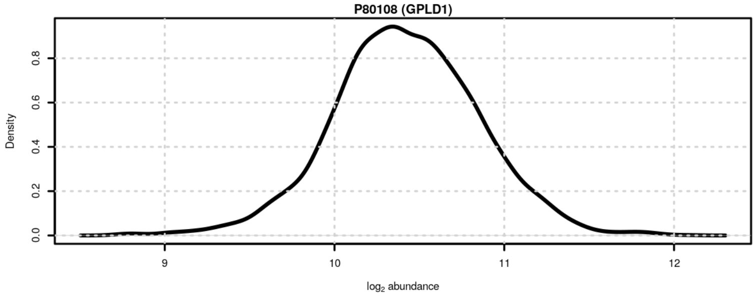

**Figure S140**: Signal distribution for P80108 (GPLD1).

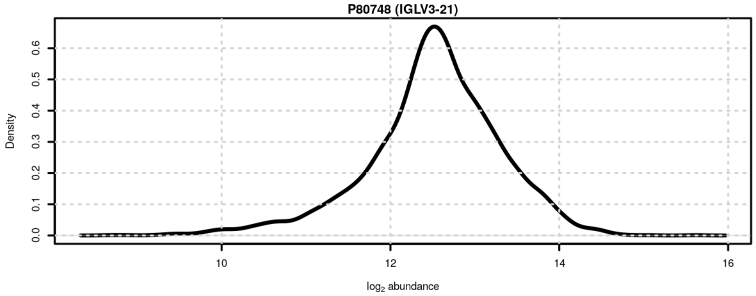

**Figure S141**: Signal distribution for P80748 (IGLV3-21).

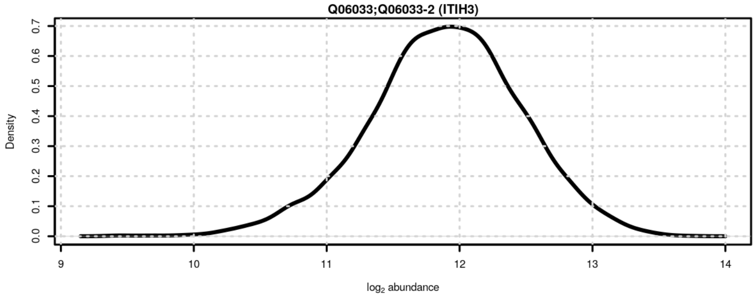

**Figure S142**: Signal distribution for Q06033;Q06033-2 (ITIH3).

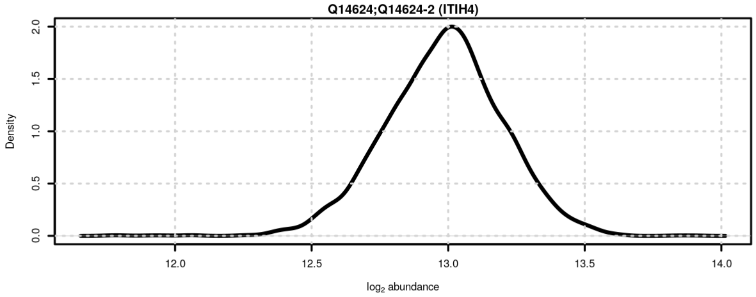

**Figure S143**: Signal distribution for Q14624;Q14624-2 (ITIH4).

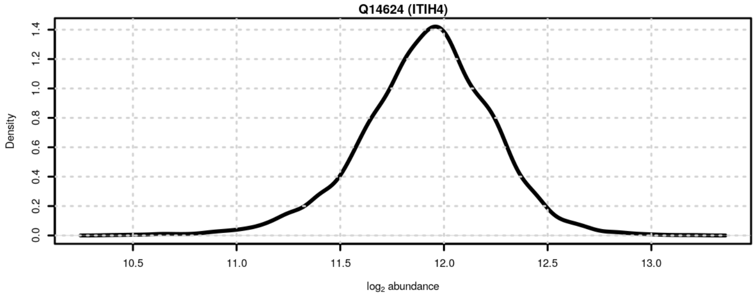

**Figure S144**: Signal distribution for Q14624 (ITIH4).

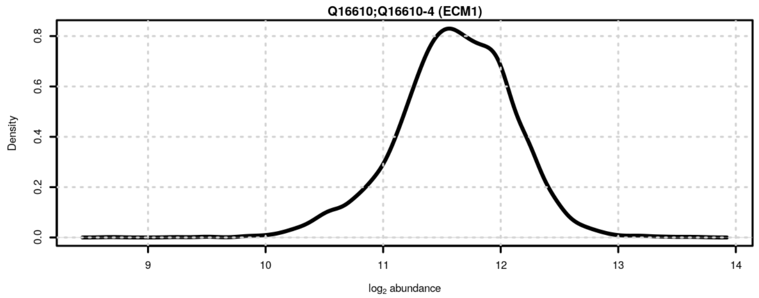

**Figure S145**: Signal distribution for Q16610;Q16610-4 (ECM1).

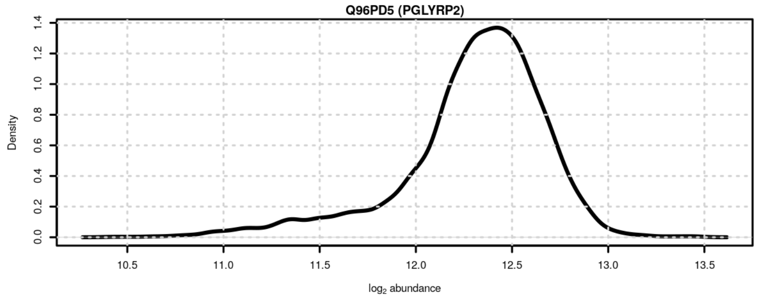

**Figure S146**: Signal distribution for Q96PD5 (PGLYRP2).

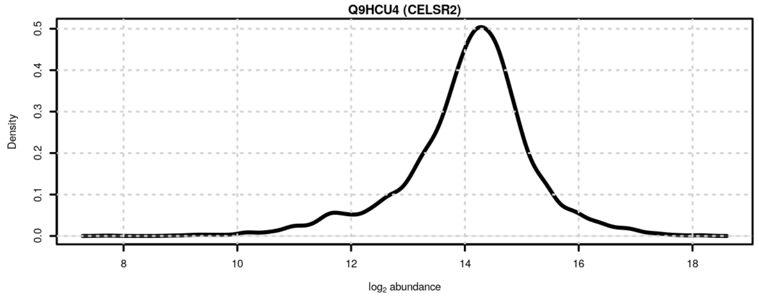

**Figure S147**: Signal distribution for Q9HCU4 (CELSR2).

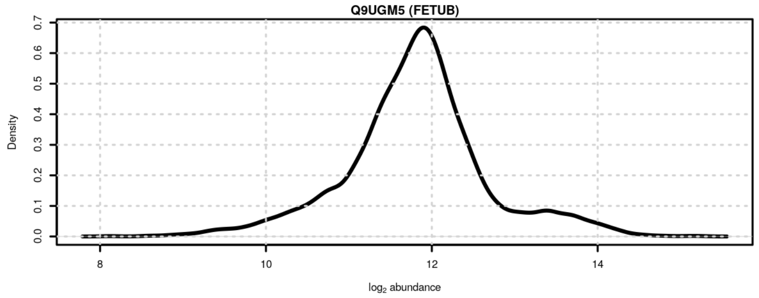

**Figure S148**: Signal distribution for Q9UGM5 (FETUB).

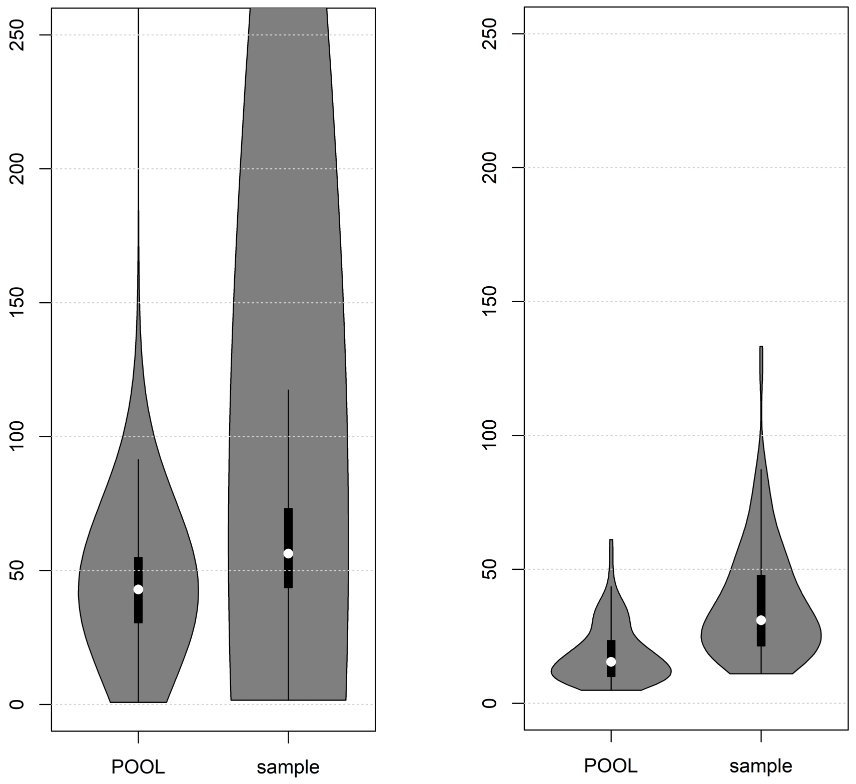

**Figure S149**: Distribution of coefficients of variation (CV, in %) before and after data pre-processing. Left: CVs of raw peptide data (DIA-NN normalized data). Right: CVs of protein data after normalization

**Table S1** (external spreadsheet, xlsx format): Results from association analyses.

**Table S2** (external spreadsheet, xlsx format): Functional annotations for detected proteins.

**Table S3**: Proteins with lowest coefficient of variation across study samples. $CV_{study}$: coefficient of variation in study samples, $CV_{QC}$: coefficient of variation in pooled QC samples and $CV_{rel}$: relative coefficient of variation expressed as the ratio between the CV in study and in pooled QC samples.

| *UniProt* | *Gene* | $CV_{study}$ | $CV_{QC}$ | $CV_{rel}$ |
|---|---|---|---|---|
| P10909;P10909-2;P10909-4;P10909-5 | CLU | 11 | 6.27 | 1.75 |
| P00734 | F2 | 13 | 7.02 | 1.85 |
| P02768 | ALB | 13.3 | 9.47 | 1.4 |
| P01008 | SERPINC1 | 13.7 | 7.37 | 1.86 |
| P02790 | HPX | 13.8 | 7.89 | 1.74 |
| P01024 | C3 | 13.8 | 4.87 | 2.84 |
| P19827 | ITIH1 | 13.9 | 6.75 | 2.05 |
| P19823 | ITIH2 | 13.9 | 8.58 | 1.62 |
| P02760 | AMBP | 14.7 | 8.8 | 1.67 |
| P01031 | C5 | 14.7 | 8.02 | 1.83 |
| P01042;P01042-2 | KNG1 | 14.8 | 8.42 | 1.76 |
| Q14624;Q14624-2 | ITIH4 | 14.9 | 7.54 | 1.97 |
| P04217 | A1BG | 15.3 | 9.36 | 1.64 |
| P02652 | APOA2 | 15.5 | 8.17 | 1.9 |
| P00747 | PLG | 15.6 | 6.52 | 2.39 |

| *UniProt* | *Gene* | $CV_{study}$ | $CV_{QC}$ | $CV_{rel}$ |
|---|---|---|---|---|
| P08603 | CFH | 15.7 | 7.93 | 1.98 |
| P02774;P02774-3 | GC | 16.3 | 6.13 | 2.66 |
| P00751 | CFB | 17 | 7.5 | 2.26 |
| P09871 | C1S | 17.5 | 11.6 | 1.51 |
| P02787 | TF | 17.8 | 7.59 | 2.35 |
| P04217;P04217-2 | A1BG | 18.2 | 10.6 | 1.72 |
| P08697 | SERPINF2 | 18.3 | 12.3 | 1.49 |
| P04004 | VTN | 18.4 | 8.55 | 2.15 |
| P25311 | AZGP1 | 18.9 | 9.66 | 1.95 |
| P02765 | AHSG | 19.1 | 9.31 | 2.05 |
| P07357 | C8A | 19.3 | 12.2 | 1.58 |
| P01011 | SERPINA3 | 19.5 | 11 | 1.77 |
| P05156 | CFI | 19.5 | 11.3 | 1.73 |
| P00736 | C1R | 19.9 | 15.1 | 1.32 |
| P06396;P06396-2 | GSN | 20 | 10.5 | 1.91 |

**Table S4**: Proteins with the highest (relative) coefficient of variation across study samples. $CV_{study}$: coefficient of variation in study samples, $CV_{QC}$: coefficient of variation in pooled QC samples and $CV_{rel}$: relative coefficient of variation expressed as the ratio between the CV in study and in pooled QC samples.

| *UniProt* | *Gene* | $CV_{study}$ | $CV_{QC}$ | $CV_{rel}$ |
|---|---|---|---|---|
| P01861 | IGHG4 | 89.2 | 12.5 | 7.15 |
| P01019 | AGT | 68.8 | 10.6 | 6.5 |
| P01871 | IGHM | 66.7 | 11 | 6.09 |
| A0A0B4J1U7 | IGHV6-1 | 80.4 | 15.2 | 5.29 |
| P01023 | A2M | 29.9 | 7.06 | 4.23 |
| P06727 | APOA4 | 29.8 | 7.04 | 4.23 |
| A0A0J9YXX1 | IGHV5-10-1 | 79.5 | 19.2 | 4.13 |
| P01877 | IGHA2 | 46.1 | 11.8 | 3.91 |
| P00738 | HP | 38.7 | 10.1 | 3.84 |
| P01859 | IGHG2 | 30.7 | 8.07 | 3.8 |
| A0A075B6I0 | IGLV8-61 | 58.7 | 15.6 | 3.76 |
| P02656 | APOC3 | 33.3 | 9.07 | 3.67 |
| P04278 | SHBG | 123 | 34.7 | 3.55 |
| P01591 | JCHAIN | 56.7 | 16 | 3.54 |
| P00739 | HPR | 41.1 | 11.9 | 3.45 |
| P01876 | IGHA1 | 44.5 | 13.1 | 3.39 |
| P05155;P05155-3 | SERPING1 | 55.7 | 16.5 | 3.38 |
| P00450 | CP | 26.9 | 8.33 | 3.23 |
| P68871 | HBB | 54.5 | 17.1 | 3.18 |
| P05546 | SERPIND1 | 25 | 7.99 | 3.13 |
| P19652 | ORM2 | 31.3 | 10 | 3.12 |
| O43866 | CD5L | 54.4 | 17.8 | 3.06 |
| P02748 | C9 | 27.7 | 9.33 | 2.96 |
| A0A0B4J1V2 | IGHV2-26 | 46.8 | 15.9 | 2.94 |
| P02750 | LRG1 | 31.3 | 10.8 | 2.89 |
| P01834 | IGKC | 22 | 7.62 | 2.89 |
| P04114 | APOB | 28.6 | 9.9 | 2.89 |
| P01024 | C3 | 13.8 | 4.87 | 2.84 |
| P08185 | SERPINA6 | 48.3 | 17.1 | 2.83 |
| P01602 | IGKV1-5 | 41.1 | 14.6 | 2.82 |

| *UniProt* | *Gene* | $CV_{study}$ | $CV_{QC}$ | $CV_{rel}$ |
|---|---|---|---|---|

**Table S5**: Common disease biomarker proteins quantified in the present data set. $CV_{study}$: coefficient of variation in study samples and $CV_{QC}$: coefficient of variation in pooled QC samples.

| *UniProt* | *Genes* | $CV_{study}$ | $CV_{QC}$ |
|---|---|---|---|
| P02768 | ALB | 13.3 | 9.47 |
| P02763 | ORM1 | 30 | 11.6 |
| P01009 | SERPINA1 | 21.4 | 7.95 |
| P08697 | SERPINF2 | 18.3 | 12.3 |
| P02765 | AHSG | 19.1 | 9.31 |
| P01023 | A2M | 29.9 | 7.06 |
| P01008 | SERPINC1 | 13.7 | 7.37 |
| P04114 | APOB | 28.6 | 9.9 |
| P00450 | CP | 26.9 | 8.33 |
| P06276 | BCHE | 37 | 34.8 |
| P09871 | C1S | 17.5 | 11.6 |
| P02745 | C1QA | 31.1 | 24 |
| P01024 | C3 | 13.8 | 4.87 |
| P0C0L4 | C4A | 46.3 | 18.8 |
| P01031 | C5 | 14.7 | 8.02 |
| P00740 | F9 | 47.2 | 30.1 |
| P00742 | F10 | 21.5 | 16.9 |
| P00488 | F13A1 | 43.7 | 30.2 |
| P05160 | F13B | 54.4 | 30.8 |
| P02671 | FGA | 71.2 | 31.1 |
| P02675 | FGB | 65.2 | 29.2 |
| P02751 | FN1 | 88 | 61.1 |
| P00738 | HP | 38.7 | 10.1 |
| P02790 | HPX | 13.8 | 7.89 |
| P08519 | LPA | 134 | 56.5 |
| P00747 | PLG | 15.6 | 6.52 |
| P02766 | TTR | 43 | 20 |
| P07225 | PROS1 | 20.6 | 14.4 |
| P02753 | RBP4 | 23.9 | 9.48 |
| P04278 | SHBG | 123 | 34.7 |
| P05543 | SERPINA7 | 35.9 | 18.3 |

**Table S6**: Correlation between clinical laboratory measurements and quantified protein abundances.

| Laboratory.parameter | UniProt.ID | HGNC.symbol | rho |
|---|---|---|---|
| Antithrombin (%) | P01008 | SERPINC1 | 0.366 |
| Albumin (g/dL) | P02768 | ALB | 0.32 |
| HDL (mg/dL) | P02647 | APOA1 | 0.408 |
| LDL (mg/dL) | P04114 | APOB | 0.787 |
| Triglycerides (mg/dL) | P02656 | APOC3 | 0.596 |
| Transferrin (mg/dL) | P02787 | TF | 0.686 |
| HGN (h/dL) | P69905 | HBA1 | 0.48 |

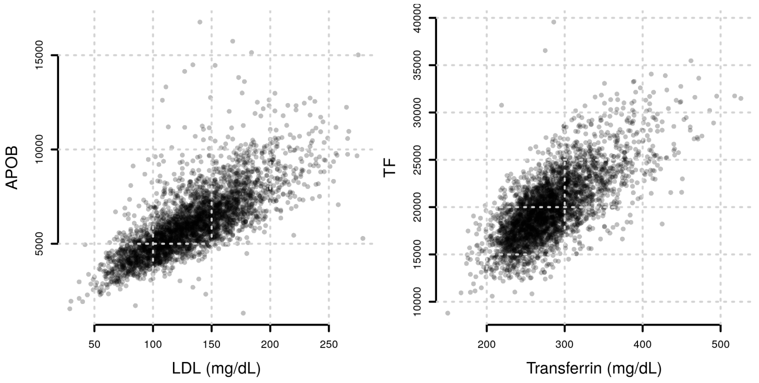

**Figure S150**: Correlation between quantified protein abundances and related diagnostic assays. Shown are MS-determind, relative protein abundances (normalized signal intensities) on the y-axis and related diagnostic assays on the x-axis.

#### Principal Component Analysis and influence of hormonal contraceptive use

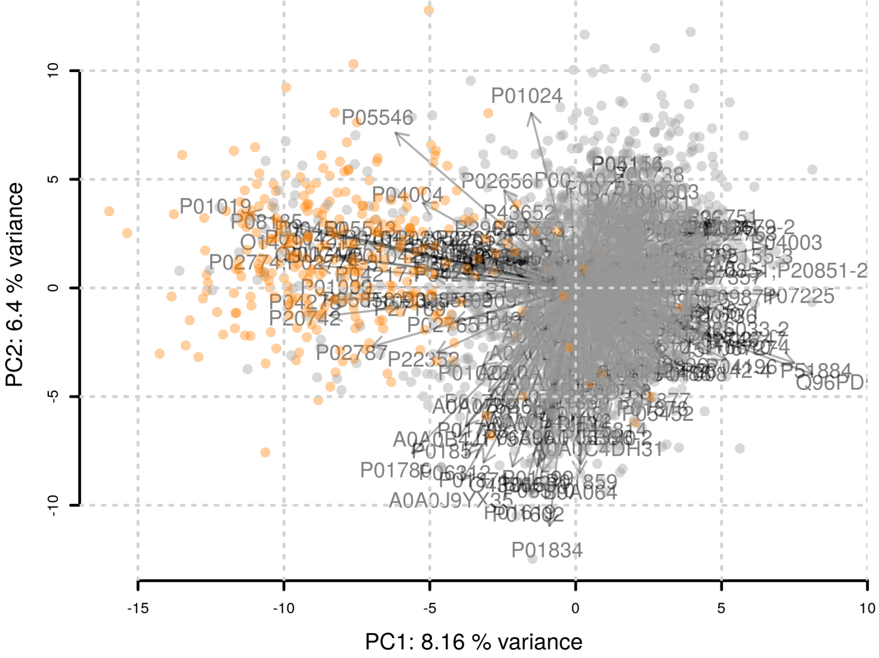

**Figure S151**: Principal component anaysis of the plasma proteome data. Data points are colored according to usage of hormonal contraceptives.

#### Previous hormonal contraceptive use

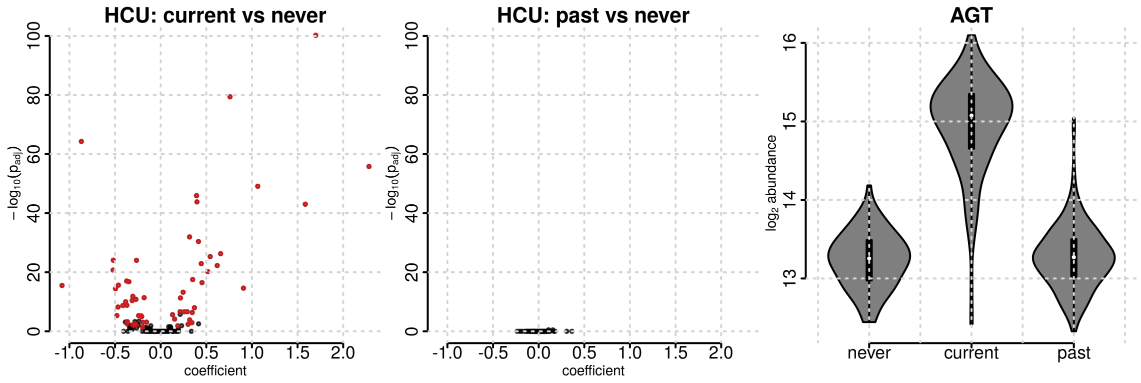

**Figure S152**: Impact of previous use of hormonal contraceptives on the plasma proteome. Left: results from the comparison between participants currently taking hormonal contraceptives and those that never took hormonal contraceptives. Middle: results from the comparison between participants that used hormonal contraceptives in the past and those that never took them. Proteins with significant differences in abundances are highlighted in red. Right: abundance of angiotensinogen (AGT) in the three hormonal contraceptive use (HCU) groups.

### Sex, age and body mass index associated proteins

#### Sex-associated plasma proteins

**Table S7**: Proteins with significant difference in abundance between female and male study participants. $coef$, $ES$ and $p_{adj}$: coefficient (representing the differential abundance in log2 scale), effect size and p-value adjusted for multiple hypothesis testing. Proteins are ordered by p-value.

| $UniProt$ | $Gene$ | $coef$ | $p_{adj}$ | $ES$ |
|---|---|---|---|---|
| P02753 | RBP4 | -0.306 | 3.11e-152 | -0.908 |
| P00450 | CP | 0.231 | 2.93e-120 | 0.668 |
| P80108 | GPLD1 | -0.306 | 3.14e-90 | -0.709 |
| P36955 | SERPINF1 | -0.202 | 2.51e-83 | -0.671 |
| P02766 | TTR | -0.387 | 1.28e-81 | -0.676 |
| P25311 | AZGP1 | -0.177 | 4e-80 | -0.662 |
| P69905 | HBA1 | -0.347 | 6.89e-78 | -0.667 |
| P02750 | LRG1 | 0.281 | 3.39e-77 | 0.647 |
| P68871 | HBB | -0.313 | 1.92e-74 | -0.654 |
| O75636 | FCN3 | -0.368 | 3.52e-70 | -0.616 |
| P04278 | SHBG | 0.633 | 5.19e-69 | 0.512 |
| P13671 | C6 | 0.199 | 3.2e-68 | 0.615 |
| P02748 | C9 | 0.199 | 4.41e-45 | 0.504 |
| P05090 | APOD | -0.191 | 1.14e-42 | -0.474 |
| P01871 | IGHM | 0.331 | 2.25e-33 | 0.436 |
| P01023 | A2M | 0.165 | 2.13e-29 | 0.398 |
| O43866 | CD5L | 0.286 | 2.24e-25 | 0.382 |
| P01861 | IGHG4 | -0.43 | 1.88e-22 | -0.37 |
| P01877 | IGHA2 | -0.209 | 2.22e-17 | -0.324 |
| P01876 | IGHA1 | -0.198 | 7.73e-15 | -0.3 |

| *UniProt* | *Gene* | *coef* | $p_{adj}$ | *ES* |
|---|---|---|---|---|
| P00739 | HPR | -0.177 | 5.15e-14 | -0.293 |
| P06727 | APOA4 | -0.117 | 3.34e-13 | -0.272 |

#### Age-associated plasma proteins

**Table S8**: Proteins significantly associated with participants' age. *coef*, *ES* and $p_{adj}$: coefficient (representing the log2 change in abundance in 10 years), effect size (for 10-year change) and p-value adjusted for multiple hypothesis testing. Proteins are ordered by p-value.

| *UniProt* | *Gene* | *coef* | $p_{adj}$ | *ES* |
|---|---|---|---|---|
| P35858;P35858-2 | IGFALS | -0.113 | 4.3e-137 | -0.255 |
| P04004 | VTN | -0.0574 | 2.75e-111 | -0.224 |
| P02787 | TF | -0.0454 | 6.94e-63 | -0.18 |
| P02765 | AHSG | -0.0505 | 7.35e-60 | -0.181 |
| P51884 | LUM | 0.0632 | 9.77e-55 | 0.162 |
| P01031 | C5 | 0.0364 | 2.18e-50 | 0.168 |
| P04114 | APOB | 0.0633 | 1.9e-43 | 0.156 |
| O14791;O14791-2;O14791-3 | APOL1 | -0.0684 | 4.71e-43 | -0.148 |
| P00734 | F2 | -0.0312 | 1.91e-42 | -0.157 |
| Q06033;Q06033-2 | ITIH3 | 0.0845 | 6.73e-42 | 0.147 |
| P01619 | IGKV3-20 | -0.0815 | 7.37e-41 | -0.153 |
| P01008 | SERPINC1 | -0.0273 | 1.21e-40 | -0.136 |
| P02774;P02774-3 | GC | -0.0277 | 7.68e-40 | -0.128 |
| P04003 | C4BPA | 0.0416 | 1.73e-38 | 0.137 |
| P02750 | LRG1 | 0.0599 | 1.12e-36 | 0.138 |
| P08697 | SERPINF2 | -0.039 | 5.28e-35 | -0.142 |
| P00742 | F10 | -0.044 | 1.13e-33 | -0.138 |
| P04217 | A1BG | 0.0308 | 1e-32 | 0.134 |
| P01602 | IGKV1-5 | -0.0734 | 3.72e-32 | -0.137 |
| P02748 | C9 | 0.0487 | 6.16e-28 | 0.124 |
| P00747 | PLG | -0.0259 | 1.09e-27 | -0.115 |
| P01703 | IGLV1-40 | -0.11 | 8.37e-27 | -0.125 |
| P01780 | IGHV3-7 | -0.0783 | 2.74e-26 | -0.122 |
| P00746 | CFD | 0.0677 | 3.15e-25 | 0.12 |
| P06312 | IGKV4-1 | -0.0687 | 3.06e-24 | -0.119 |
| P01871 | IGHM | -0.086 | 4.53e-23 | -0.113 |
| P02760 | AMBP | 0.0245 | 2.72e-22 | 0.113 |
| O43866 | CD5L | -0.0834 | 3.97e-22 | -0.111 |
| P06310 | IGKV2-30 | -0.0649 | 6.88e-22 | -0.112 |
| P27169 | PON1 | -0.0456 | 4.99e-19 | -0.105 |
| P02768 | ALB | -0.0202 | 9.83e-19 | -0.101 |
| P19827 | ITIH1 | -0.0209 | 6.46e-17 | -0.1 |
| P08185 | SERPINA6 | -0.0463 | 4.77e-16 | -0.0815 |
| P20851;P20851-2 | C4BPB | 0.0443 | 7.89e-16 | 0.0915 |
| A0A0J9YX35 | IGHV3-64D | -0.0504 | 1.96e-15 | -0.0952 |
| P05452 | CLEC3B | -0.0293 | 2.54e-15 | -0.0916 |
| P04217;P04217-2 | A1BG | 0.0242 | 4.8e-15 | 0.0922 |
| P01877 | IGHA2 | 0.0608 | 5.16e-15 | 0.0941 |
| P00748 | F12 | -0.0434 | 7.9e-15 | -0.0925 |
| P02790 | HPX | 0.0182 | 4.09e-14 | 0.0903 |

| *UniProt* | *Gene* | *coef* | $p_{adj}$ | *ES* |
|---|---|---|---|---|
| P80748 | IGLV3-21 | -0.0727 | 5.66e-14 | -0.0918 |
| P04196 | HRG | 0.032 | 2.12e-13 | 0.0822 |
| P01011 | SERPINA3 | 0.0247 | 2.83e-13 | 0.0885 |
| P00738 | HP | 0.0558 | 3.7e-13 | 0.0878 |
| P01705 | IGLV2-23 | -0.0744 | 5.4e-13 | -0.0886 |
| P02766 | TTR | -0.0475 | 9.76e-13 | -0.0829 |
| P10643 | C7 | 0.0347 | 2.09e-12 | 0.0853 |
| P01019 | AGT | -0.0406 | 2.11e-12 | -0.0615 |
| P06727 | APOA4 | 0.0349 | 5.57e-12 | 0.081 |
| P01876 | IGHA1 | 0.0545 | 2e-11 | 0.0827 |
| P02749 | APOH | 0.0338 | 3.91e-11 | 0.0807 |
| Q96PD5 | PGLYRP2 | 0.0258 | 4.78e-11 | 0.068 |
| Q9UGM5 | FETUB | -0.0616 | 5.65e-11 | -0.0713 |
| P01042;P01042-2 | KNG1 | -0.0152 | 5.18e-10 | -0.0731 |
| P01599 | IGKV1-17 | -0.0598 | 1.32e-09 | -0.077 |
| P05090 | APOD | 0.0284 | 2.2e-09 | 0.0703 |
| A0A0C4DH31 | IGHV1-18 | -0.0585 | 2.45e-08 | -0.0724 |
| A0A075B6I0 | IGLV8-61 | -0.0559 | 4.57e-08 | -0.0712 |
| P02656 | APOC3 | 0.0306 | 4.85e-08 | 0.0705 |
| P05156 | CFI | 0.019 | 1.07e-07 | 0.0662 |
| P08603 | CFH | 0.0144 | 1.85e-07 | 0.0627 |
| P22352 | GPX3 | -0.031 | 8.94e-07 | -0.0633 |
| P03952 | KLKB1 | -0.0203 | 1.63e-06 | -0.0635 |
| P23142;P23142-4 | FBLN1 | 0.0372 | 2.17e-06 | 0.0632 |
| P01042-2 | KNG1 | -0.0237 | 6.31e-06 | -0.0585 |
| A0A075B6J9 | IGLV2-18 | -0.061 | 8.46e-06 | -0.0615 |
| P25311 | AZGP1 | 0.0151 | 1.05e-05 | 0.0566 |
| A0A0B4J1V2 | IGHV2-26 | -0.0416 | 1.28e-05 | -0.0607 |
| A0A0B4J1Y9 | IGHV3-72 | -0.0514 | 1.33e-05 | -0.0605 |
| P01701 | IGLV1-51 | -0.0429 | 0.000169 | -0.0553 |
| P80108 | GPLD1 | -0.0213 | 0.000436 | -0.0493 |
| P09871 | C1S | 0.0139 | 0.000531 | 0.0517 |
| P0C0L4 | C4A | 0.0366 | 0.000636 | 0.0518 |
| A0A0A0MS15 | IGHV3-49 | -0.0294 | 0.000679 | -0.0521 |
| P02679;P02679-2 | FGG | 0.0392 | 0.000683 | 0.052 |
| P19652 | ORM2 | -0.021 | 0.00129 | -0.0482 |
| Q16610;Q16610-4 | ECM1 | -0.0241 | 0.00168 | -0.0486 |
| P02652 | APOA2 | -0.0106 | 0.00172 | -0.0482 |
| P23142 | FBLN1 | 0.0334 | 0.00239 | 0.0483 |
| P02675 | FGB | 0.0392 | 0.00245 | 0.0488 |
| Q9HCU4 | CELSR2 | 0.0543 | 0.00309 | 0.0482 |
| P01857 | IGHG1 | -0.0173 | 0.00389 | -0.0474 |
| P43652 | AFM | -0.0139 | 0.00515 | -0.045 |
| P02649 | APOE | 0.022 | 0.00765 | 0.0456 |
| P01023 | A2M | -0.0175 | 0.00899 | -0.0422 |
| P07225 | PROS1 | 0.0126 | 0.011 | 0.0409 |
| P10909;P10909-2;P10909-4;P10909-5 | CLU | -0.00702 | 0.0116 | -0.0442 |
| P05543 | SERPINA7 | -0.02 | 0.0167 | -0.0391 |
| P00450 | CP | 0.0111 | 0.0229 | 0.0323 |
| O75882-2 | ATRN | 0.0143 | 0.0271 | 0.0425 |
| P02671 | FGA | 0.034 | 0.0494 | 0.0408 |

The age association for the 2 most significant proteins is shown below.

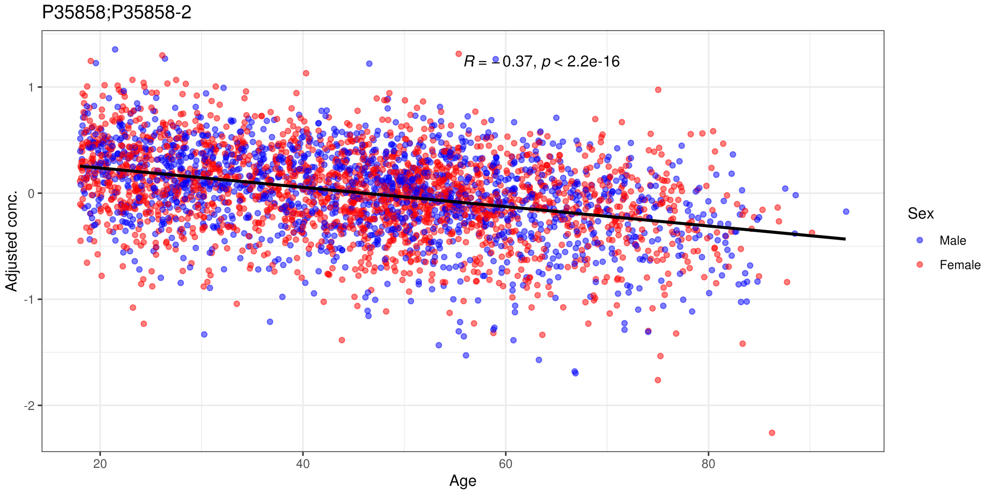

**Figure S153**: Age dependency of the protein IGFALS. Shown are sex, BMI, HCU and fasting status adjusted abundances against participants' age. The solid black line represents the linear regression line..

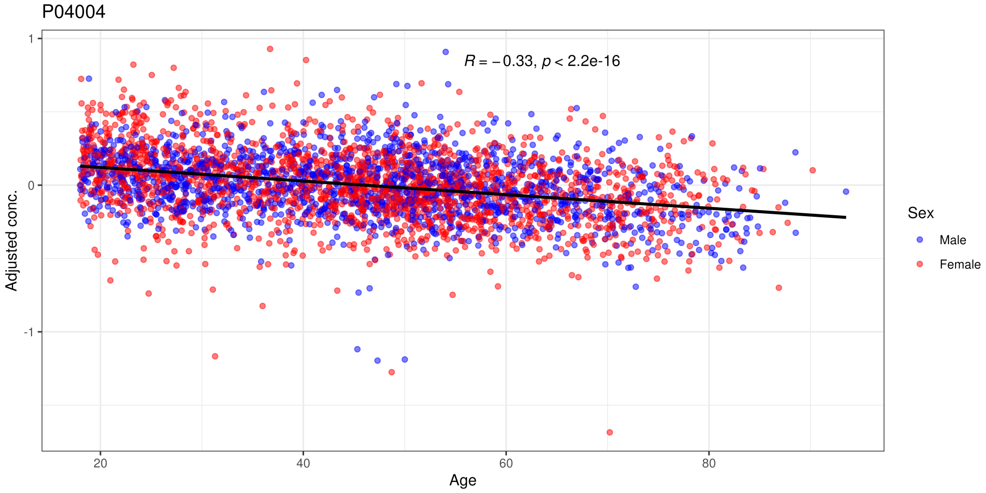

**Figure S154**: Age dependency of the protein VTN. Shown are sex, BMI, HCU and fasting status adjusted abundances against participants' age. The solid black line represents the linear regression line..

#### Body mass index associated plasma proteins

Proteins with significant differences in concentrations between body mass index (BMI) categories 1 (BMI < 18.5), 3 (25 <= BMI < 30) and 4 (BMI > 30) to the *normal* category 2 (18.5 <= BMI < 25) are listed in the tables below. Proteins with an adjusted p-value smaller than 0.05 and a difference in concentrations larger than the coefficient of variation in QC (study pool) samples are considered significant.

**Table S9**: Proteins with significant difference in abundance between participants with a BMI < 18.5 (BMI1) and participants with a BMI between 18.5 and 25 (BMI2; normal). $coef$, $ES$ and $p_{adj}$: coefficient (representing the differential abundance in log2 scale), effect size and p-value adjusted for multiple hypothesis testing. Proteins are ordered by p-value.

| *UniProt* | *Gene* | $coef$ | $p_{adj}$ | $ES$ |
|---|---|---|---|---|
| P08603 | CFH | -0.132 | 0.00215 | -0.578 |
| P01023 | A2M | 0.237 | 0.00405 | 0.571 |
| P02763 | ORM1 | -0.218 | 0.0323 | -0.511 |
| P01024 | C3 | -0.0961 | 0.0418 | -0.481 |

**Table S10**: Proteins with significant difference in abundance between participants with a BMI between 25 and 30 (BMI3) and participants with a BMI between 18.5 and 25 (BMI2; normal). $coef$, $ES$ and $p_{adj}$: coefficient (representing the differential abundance in log2 scale) effect size and p-value adjusted for multiple hypothesis testing. Proteins are ordered by p-value.

| *UniProt* | *Gene* | $coef$ | $p_{adj}$ | $ES$ |
|---|---|---|---|---|
| P01024 | C3 | 0.112 | 6.38e-52 | 0.561 |
| P01023 | A2M | -0.199 | 3.42e-36 | -0.48 |
| P05090 | APOD | -0.181 | 2.55e-32 | -0.448 |
| P06727 | APOA4 | -0.12 | 1.47e-11 | -0.277 |

**Table S11**: Proteins with significant difference in abundance between participants with a BMI > 30 (BMI4) and participants with a BMI between 18.5 and 25 (BMI2; normal). $coef$, $ES$ and $p_{adj}$: coefficient (representing the differential abundance in log2 scale) effect size and p-value adjusted for multiple hypothesis testing. Proteins are ordered by p-value.

| *UniProt* | *Gene* | $coef$ | $p_{adj}$ | $ES$ |
|---|---|---|---|---|
| P01024 | C3 | 0.234 | 1.62e-128 | 1.17 |
| P05090 | APOD | -0.401 | 1.16e-91 | -0.992 |
| P08603 | CFH | 0.213 | 7.13e-83 | 0.933 |
| P01008 | SERPINC1 | -0.151 | 6.26e-61 | -0.755 |
| P43652 | AFM | 0.247 | 7.59e-56 | 0.8 |
| P04004 | VTN | 0.179 | 4.34e-55 | 0.698 |
| P05156 | CFI | 0.219 | 6.13e-52 | 0.761 |
| P00751 | CFB | 0.167 | 7.25e-41 | 0.692 |
| O95445 | APOM | -0.23 | 2.04e-36 | -0.651 |
| P02774;P02774-3 | GC | -0.112 | 5.04e-32 | -0.515 |
| P01023 | A2M | -0.242 | 8.25e-32 | -0.583 |
| P25311 | AZGP1 | -0.151 | 3.62e-30 | -0.564 |
| P04278 | SHBG | -0.577 | 2.09e-29 | -0.466 |
| P06727 | APOA4 | -0.207 | 5.29e-21 | -0.48 |
| P05546 | SERPIND1 | 0.15 | 3.75e-16 | 0.423 |
| P02763 | ORM1 | 0.176 | 2.67e-15 | 0.411 |
| P00738 | HP | 0.245 | 1.8e-12 | 0.386 |
| O43866 | CD5L | -0.277 | 8.28e-12 | -0.37 |
| P19652 | ORM2 | 0.141 | 5.59e-09 | 0.324 |
| P01871 | IGHM | -0.244 | 7.78e-09 | -0.321 |

#### Proteins significantly associated with fasting status

**Table S12**: Proteins with significant difference in abundance between participants declared to not have fasted against those who did. $coef$, $ES$ and $p_{adj}$: coefficient (representing the differential abundance in log2 scale) effect size and p-value adjusted for multiple hypothesis testing. Proteins are ordered by p-value.

| $UniProt$ | $Gene$ | $coef$ | $p_{adj}$ | $ES$ |
|---|---|---|---|---|
| P06727 | APOA4 | 0.126 | 0.00158 | 0.291 |

### Plasma proteins associated with usage of hormonal contraceptives

A comparison of the results from the analysis on the full data set and on the subset of female participants below 40 years of age is shown below. Coefficients for associations with hormonal contraceptive use are highly similar between the two analyses and p-values highly related (same rank, but difference in the values due to the differences in statistical power from the two analyses).

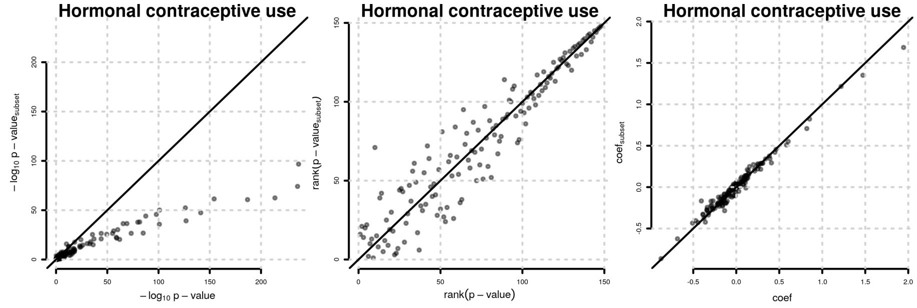

**Figure S155**: Comparison of results for association with hormonal contraceptive use from the analysis of the full data set (x-axis) and the analysis on the subset of female participants below 40 years of age (y axis). Shown are -log10 of p-values, ranks of p-values and coefficients.

#### Sensitivity analysis

To evaluate the influence of hormonal contraceptive use on results for age, sex and BMI associations, the analysis was repeated without adjustment for hormonal contraceptive use. Proteins that were found significantly associated with sex in this analysis, but were no longer significant after adjustment for hormonal contraceptive use are listed in the table below.

**Table S13**: Proteins significantly associated with sex in an analysis without adjusting for hormonal contraceptive use that were no longer significant after adjustment. $coef$ and $p$: coefficient and bonferroni adjusted p-value from the analysis without adjustment for hormonal contraceptive use. $coef_{adj}$, $p_{adj}$, $coef_{HCU}$ and $p_{HCU}$: coefficients and bonferroni adjusted p-values for sex association and hormoman contraceptive use association from the analysis with adjustment for hormonal contraceptive use.

| $Gene$ | $coef$ | $p$ | $coef_{adj}$ | $p_{adj}$ | $coef_{HCU}$ | $p_{HCU}$ |
|---|---|---|---|---|---|---|
| SERPINA7 | 0.308 | 5.29e-70 | 0.216 | 8.47e-36 | 0.583 | 2.56e-78 |
| PZP | 0.476 | 3.66e-63 | 0.347 | 9.28e-34 | 0.82 | 2.45e-57 |
| AGT | 0.35 | 1.89e-58 | 0.118 | 3.76e-10 | 1.47 | 0 |
| FETUB | 0.452 | 7.8e-54 | 0.26 | 1.74e-19 | 1.22 | 9.73e-125 |
| SERPINA6 | 0.274 | 1.16e-48 | 0.14 | 3.46e-14 | 0.852 | 7.89e-153 |

| *Gene* | $coef$ | $p$ | $coef_{adj}$ | $p_{adj}$ | $coef_{HCU}$ | $p_{HCU}$ |
|---|---|---|---|---|---|---|
| PGLYRP2 | -0.169 | 1.42e-38 | -0.061 | 1e-05 | -0.683 | 3.7e-212 |
| SERPINA1 | 0.135 | 6.47e-37 | 0.0734 | 3.55e-11 | 0.392 | 2e-96 |
| SERPING1 | -0.314 | 1.17e-20 | -0.176 | 5.86e-06 | -0.874 | 4.95e-48 |

Proteins significantly associated with age from an analysis without adjustment for hormonal contraceptive use that were no longer significant after adjustment for hormonal contraceptive use are listed in the table below.

**Table S14**: Proteins significantly associated with age in an analysis without adjusting for hormonal contraceptive use that were no longer significant after adjustment. $coef$ and $p$: coefficient and bonferroni adjusted p-value from the analysis without adjustment for hormonal contraceptive use. $coef_{adj}$, $p_{adj}$, $coef_{HCU}$ and $p_{HCU}$: coefficients and bonferroni adjusted p-values for age association and hormonal contraceptive use association from the analysis with adjustment for hormonal contraceptive use.

| *Gene* | $coef$ | $p$ | $coef_{adj}$ | $p_{adj}$ | $coef_{HCU}$ | $p_{HCU}$ |
|---|---|---|---|---|---|---|
| SERPINA1 | -0.0299 | 4.79e-19 | -0.00795 | 1 | 0.392 | 2e-96 |
| C6 | 0.0293 | 6.8e-16 | 0.0113 | 0.143 | -0.32 | 9.39e-55 |
| SHBG | -0.0965 | 1.8e-14 | 0.0126 | 1 | 1.95 | 2.06e-185 |
| PZP | -0.0676 | 1.91e-13 | -0.0216 | 1 | 0.82 | 2.45e-57 |
| SERPIND1 | -0.0299 | 3.27e-13 | -0.00828 | 1 | 0.385 | 4.97e-64 |
| SERPING1 | 0.073 | 1.47e-11 | 0.024 | 1 | -0.874 | 4.95e-48 |
| C3 | -0.0124 | 5.27e-08 | -0.00522 | 1 | 0.127 | 1.32e-24 |
| C1QC | 0.0207 | 1.87e-07 | 0.00875 | 1 | -0.213 | 6.2e-23 |
| HPR | -0.0365 | 2.35e-06 | -0.0207 | 0.323 | 0.281 | 1.03e-10 |
| ORM1 | 0.0247 | 4.01e-06 | 0.0037 | 1 | -0.375 | 1.26e-42 |
| C1QB | 0.0179 | 7.1e-05 | 0.00755 | 1 | -0.184 | 1.7e-15 |
| CD14 | 0.0321 | 9.31e-05 | 0.0139 | 1 | -0.325 | 1.45e-14 |
| C1R | 0.0158 | 0.000111 | 0.00616 | 1 | -0.172 | 8.53e-17 |
| FN1 | 0.0372 | 0.0319 | 0.0208 | 1 | -0.293 | 0.000248 |
| APOM | 0.0135 | 0.0414 | 0.00366 | 1 | -0.175 | 1.2e-12 |

### Plasma proteins associated with medication (ATC level 3)

Associations were evaluated for 27 ATC level 3 medications taken on a regular basis (at least twice per week) by more than 14 study participants. Significant associations are shown for each medication in the tables below. For 11 medications no significant association was identified.

**Table S15**: Proteins significantly associated with ATC level 3 medication *HORMONAL CONTRACEPTIVES FOR SYSTEMIC USE* (ATC3 G03A). $coef$ and $p_{adj}$: coefficient (representing the differential abundance in log2 scale) and p-value adjusted for multiple hypothesis testing. $ES$ effect size. Proteins are ordered by p-value.

| *UniProt* | *Gene* | $coef$ | $p_{adj}$ | $ES$ |
|---|---|---|---|---|
| P01019 | AGT | 1.46 | 0 | 2.21 |
| P00450 | CP | 0.589 | 3.01e-206 | 1.71 |
| Q96PD5 | PGLYRP2 | -0.657 | 4.3e-179 | -1.73 |
| P04278 | SHBG | 1.86 | 1.51e-157 | 1.51 |
| P08185 | SERPINA6 | 0.84 | 9e-135 | 1.48 |
| P02774;P02774-3 | GC | 0.309 | 3.46e-128 | 1.42 |

| *UniProt* | *Gene* | *coef* | $p_{adj}$ | *ES* |
|---|---|---|---|---|
| Q9UGM5 | FETUB | 1.22 | 1.15e-114 | 1.42 |
| P01008 | SERPINC1 | -0.273 | 4.4e-106 | -1.36 |
| P00747 | PLG | 0.299 | 6.25e-98 | 1.33 |
| P04196 | HRG | -0.496 | 6.92e-86 | -1.28 |
| P01009 | SERPINA1 | 0.383 | 1.4e-85 | 1.24 |
| P01042;P01042-2 | KNG1 | 0.254 | 1.32e-76 | 1.22 |
| P51884 | LUM | -0.449 | 8.51e-75 | -1.15 |
| P04004 | VTN | 0.28 | 2.26e-74 | 1.09 |
| P05543 | SERPINA7 | 0.57 | 4.04e-68 | 1.11 |
| P07225 | PROS1 | -0.333 | 5.97e-63 | -1.08 |
| P05546 | SERPIND1 | 0.391 | 8.15e-60 | 1.1 |
| P01042-2 | KNG1 | 0.438 | 1.21e-58 | 1.08 |
| O14791;O14791-2;O14791-3 | APOL1 | 0.468 | 1.6e-54 | 1.02 |
| P05452 | CLEC3B | -0.324 | 2.04e-51 | -1.01 |
| P13671 | C6 | -0.321 | 4.84e-50 | -0.992 |
| P04003 | C4BPA | -0.282 | 6.7e-49 | -0.929 |
| P05155;P05155-3 | SERPING1 | -0.878 | 3.34e-44 | -0.949 |
| P20742 | PZP | 0.752 | 4.72e-44 | 0.896 |
| P20851;P20851-2 | C4BPB | -0.445 | 1.43e-43 | -0.919 |
| P19652 | ORM2 | -0.377 | 1.03e-36 | -0.866 |
| P02763 | ORM1 | -0.359 | 1.03e-35 | -0.84 |
| P80108 | GPLD1 | 0.354 | 1.33e-35 | 0.822 |
| P02753 | RBP4 | 0.265 | 7.03e-35 | 0.789 |
| P06727 | APOA4 | -0.346 | 5.35e-33 | -0.802 |
| Q16610;Q16610-4 | ECM1 | -0.38 | 1.91e-27 | -0.765 |
| P07357 | C8A | -0.211 | 1.73e-25 | -0.744 |
| Q06033;Q06033-2 | ITIH3 | -0.388 | 2.56e-24 | -0.676 |
| P02768 | ALB | -0.134 | 2.02e-22 | -0.667 |
| P35858;P35858-2 | IGFALS | 0.266 | 4.24e-22 | 0.601 |
| P01024 | C3 | 0.123 | 8.19e-21 | 0.614 |
| P00748 | F12 | 0.306 | 2.49e-20 | 0.653 |
| P02747 | C1QC | -0.208 | 1.24e-19 | -0.65 |
| P02787 | TF | 0.134 | 3.76e-15 | 0.53 |
| P02790 | HPX | 0.111 | 2.46e-14 | 0.547 |
| P02746 | C1QB | -0.181 | 1.42e-13 | -0.549 |
| P02656 | APOC3 | 0.229 | 9.57e-13 | 0.529 |
| P00739 | HPR | 0.299 | 5.22e-11 | 0.494 |
| P27169 | PON1 | 0.2 | 7.97e-10 | 0.459 |
| P02748 | C9 | -0.166 | 1.33e-08 | -0.42 |
| O43866 | CD5L | -0.298 | 2.27e-07 | -0.398 |
| A0A075B6I0 | IGLV8-61 | -0.31 | 1.11e-06 | -0.395 |
| P02649 | APOE | -0.187 | 1.62e-06 | -0.388 |
| P01602 | IGKV1-5 | -0.196 | 6.85e-06 | -0.365 |
| P01871 | IGHM | -0.245 | 0.000116 | -0.323 |

**Table S16**: Proteins significantly associated with ATC level 3 medication *ANTIANDROGENS* (ATC3 G03H). $coef$ and $p_{adj}$: coefficient (representing the differential abundance in log2 scale) and p-value adjusted for multiple hypothesis testing. $ES$ effect size. Proteins are ordered by p-value.

| *UniProt* | *Gene* | $coef$ | $p_{adj}$ | $ES$ |
|---|---|---|---|---|
| P01019 | AGT | 1.68 | 5.77e-43 | 2.55 |
| P00450 | CP | 0.85 | 9.01e-36 | 2.46 |
| Q96PD5 | PGLYRP2 | -0.972 | 4.27e-32 | -2.57 |
| P04278 | SHBG | 2.78 | 2.76e-28 | 2.25 |
| P08185 | SERPINA6 | 1.09 | 6.52e-18 | 1.92 |
| P01008 | SERPINC1 | -0.383 | 3.64e-16 | -1.91 |
| P02774;P02774-3 | GC | 0.388 | 8.44e-16 | 1.78 |
| Q9UGM5 | FETUB | 1.56 | 2e-14 | 1.81 |
| P04196 | HRG | -0.646 | 5.69e-11 | -1.66 |
| P00747 | PLG | 0.361 | 7.85e-11 | 1.61 |
| P02763 | ORM1 | -0.72 | 1.75e-10 | -1.69 |
| P20742 | PZP | 1.33 | 5.1e-10 | 1.58 |
| P51884 | LUM | -0.597 | 7.85e-10 | -1.53 |
| P05155;P05155-3 | SERPING1 | -1.49 | 2.65e-09 | -1.62 |
| P01042;P01042-2 | KNG1 | 0.323 | 3.81e-09 | 1.55 |
| P05543 | SERPINA7 | 0.76 | 6.58e-09 | 1.48 |
| P04217;P04217-2 | A1BG | 0.406 | 2.36e-08 | 1.55 |
| P05452 | CLEC3B | -0.462 | 1.75e-07 | -1.44 |
| P01042-2 | KNG1 | 0.583 | 1.76e-07 | 1.44 |
| P07225 | PROS1 | -0.427 | 1.85e-07 | -1.39 |
| P20851;P20851-2 | C4BPB | -0.674 | 3.93e-07 | -1.39 |
| P04003 | C4BPA | -0.39 | 1.28e-06 | -1.28 |
| P01009 | SERPINA1 | 0.385 | 3.42e-06 | 1.24 |
| P04004 | VTN | 0.299 | 4.65e-06 | 1.17 |
| P13671 | C6 | -0.388 | 5.19e-05 | -1.2 |
| P02768 | ALB | -0.241 | 7.19e-05 | -1.19 |
| P19652 | ORM2 | -0.508 | 0.000176 | -1.17 |
| P05546 | SERPIND1 | 0.382 | 0.000959 | 1.07 |
| P00738 | HP | -0.677 | 0.00203 | -1.06 |
| Q16610;Q16610-4 | ECM1 | -0.523 | 0.00272 | -1.05 |
| P02747 | C1QC | -0.327 | 0.00503 | -1.02 |
| P01011 | SERPINA3 | -0.278 | 0.00741 | -0.996 |
| O14791;O14791-2;O14791-3 | APOL1 | 0.428 | 0.00828 | 0.929 |
| P02656 | APOC3 | 0.427 | 0.0104 | 0.985 |
| P00748 | F12 | 0.446 | 0.014 | 0.951 |
| P07358 | C8B | -0.345 | 0.0163 | -0.964 |
| P08571 | CD14 | -0.553 | 0.0324 | -0.922 |
| P00736 | C1R | -0.268 | 0.0447 | -0.897 |

**Table S17**: Proteins significantly associated with ATC level 3 medication *CONTRACEPTIVES FOR TOPICAL USE* (ATC3 G02B). $coef$ and $p_{adj}$: coefficient (representing the differential abundance in log2 scale) and p-value adjusted for multiple hypothesis testing. $ES$ effect size. Proteins are ordered by p-value.

| *UniProt* | *Gene* | $coef$ | $p_{adj}$ | $ES$ |
|---|---|---|---|---|
| P01019 | AGT | 0.685 | 4.22e-20 | 1.04 |
| P00450 | CP | 0.272 | 4.55e-10 | 0.788 |
| Q96PD5 | PGLYRP2 | -0.29 | 1.01e-07 | -0.766 |

| *UniProt* | *Gene* | *coef* | $p_{adj}$ | *ES* |
|---|---|---|---|---|
| P02774;P02774-3 | GC | 0.164 | 1.04e-07 | 0.756 |
| P08185 | SERPINA6 | 0.41 | 8.82e-07 | 0.722 |
| P00747 | PLG | 0.17 | 1.56e-06 | 0.759 |
| P80108 | GPLD1 | 0.339 | 1.94e-06 | 0.786 |
| P04278 | SHBG | 0.799 | 3.64e-06 | 0.647 |
| P04003 | C4BPA | -0.222 | 5.5e-06 | -0.731 |
| P05543 | SERPINA7 | 0.333 | 0.000177 | 0.65 |
| P01876 | IGHA1 | -0.446 | 0.000594 | -0.677 |
| P02760 | AMBP | -0.137 | 0.00208 | -0.631 |
| P01009 | SERPINA1 | 0.177 | 0.00245 | 0.57 |
| Q9UGM5 | FETUB | 0.479 | 0.00278 | 0.555 |
| P05546 | SERPIND1 | 0.213 | 0.00358 | 0.599 |
| P01008 | SERPINC1 | -0.104 | 0.00951 | -0.519 |
| P02765 | AHSG | 0.148 | 0.0233 | 0.533 |
| P02787 | TF | 0.123 | 0.0487 | 0.489 |
| P02753 | RBP4 | 0.162 | 0.0489 | 0.48 |

**Table S18**: Proteins significantly associated with ATC level 3 medication *ANTIINFLAMMATORY AND ANTIRHEUMATIC PRODUCTS, NON-STEROIDS* (ATC3 M01A). *coef* and $p_{adj}$: coefficient (representing the differential abundance in log2 scale) and p-value adjusted for multiple hypothesis testing. *ES* effect size. Proteins are ordered by p-value.

| *UniProt* | *Gene* | *coef* | $p_{adj}$ | *ES* |
|---|---|---|---|---|
| P02748 | C9 | 0.249 | 0.00066 | 0.632 |
| P01011 | SERPINA3 | 0.161 | 0.00752 | 0.579 |
| P02750 | LRG1 | 0.236 | 0.00804 | 0.542 |

**Table S19**: Proteins significantly associated with ATC level 3 medication *DOPAMINERGIC AGENTS* (ATC3 N04B). *coef* and $p_{adj}$: coefficient (representing the differential abundance in log2 scale) and p-value adjusted for multiple hypothesis testing. *ES* effect size. Proteins are ordered by p-value.

| *UniProt* | *Gene* | *coef* | $p_{adj}$ | *ES* |
|---|---|---|---|---|
| P02748 | C9 | 0.364 | 0.0157 | 0.921 |

**Table S20**: Proteins significantly associated with ATC level 3 medication *BLOOD GLUCOSE LOWERING DRUGS, EXCL. INSULINS* (ATC3 A10B). *coef* and $p_{adj}$: coefficient (representing the differential abundance in log2 scale) and p-value adjusted for multiple hypothesis testing. *ES* effect size. Proteins are ordered by p-value.

| *UniProt* | *Gene* | *coef* | $p_{adj}$ | *ES* |
|---|---|---|---|---|
| P06727 | APOA4 | 0.332 | 1.44e-06 | 0.769 |
| P02649 | APOE | -0.3 | 0.00166 | -0.623 |
| P05090 | APOD | -0.224 | 0.00331 | -0.556 |
| P02654 | APOC1 | -0.428 | 0.0207 | -0.539 |

**Table S21**: Proteins significantly associated with ATC level 3 medication *ANTIGOUT PREPARATIONS* (ATC3 M04A). $coef$ and $p_{adj}$: coefficient (representing the differential abundance in log2 scale) and p-value adjusted for multiple hypothesis testing. $ES$ effect size. Proteins are ordered by p-value.

| *UniProt* | *Gene* | $coef$ | $p_{adj}$ | $ES$ |
|---|---|---|---|---|
| P00734 | F2 | -0.137 | 0.00415 | -0.688 |
| P02763 | ORM1 | 0.258 | 0.032 | 0.604 |

**Table S22**: Proteins significantly associated with ATC level 3 medication *ANTITHROMBOTIC AGENTS* (ATC3 B01A). $coef$ and $p_{adj}$: coefficient (representing the differential abundance in log2 scale) and p-value adjusted for multiple hypothesis testing. $ES$ effect size. Proteins are ordered by p-value.

| *UniProt* | *Gene* | $coef$ | $p_{adj}$ | $ES$ |
|---|---|---|---|---|
| P00734 | F2 | -0.136 | 6.69e-15 | -0.684 |
| P01023 | A2M | 0.224 | 8.09e-10 | 0.54 |

**Table S23**: Proteins significantly associated with ATC level 3 medication *BETA BLOCKING AGENTS* (ATC3 C07A). $coef$ and $p_{adj}$: coefficient (representing the differential abundance in log2 scale) and p-value adjusted for multiple hypothesis testing. $ES$ effect size. Proteins are ordered by p-value.

| *UniProt* | *Gene* | $coef$ | $p_{adj}$ | $ES$ |
|---|---|---|---|---|
| P00734 | F2 | -0.106 | 3.84e-07 | -0.532 |

**Table S24**: Proteins significantly associated with ATC level 3 medication *DRUGS USED IN BENIGN PROSTATIC HYPERTROPHY* (ATC3 G04C). $coef$ and $p_{adj}$: coefficient (representing the differential abundance in log2 scale) and p-value adjusted for multiple hypothesis testing. $ES$ effect size. Proteins are ordered by p-value.

| *UniProt* | *Gene* | $coef$ | $p_{adj}$ | $ES$ |
|---|---|---|---|---|
| P04278 | SHBG | 0.647 | 0.00171 | 0.523 |
| A0A0J9YX35 | IGHV3-64D | 0.342 | 0.00259 | 0.647 |
| P01023 | A2M | 0.246 | 0.00371 | 0.593 |
| P02753 | RBP4 | -0.173 | 0.0273 | -0.515 |

**Table S25**: Proteins significantly associated with ATC level 3 medication *ANTIEPILEPTICS* (ATC3 N03A). $coef$ and $p_{adj}$: coefficient (representing the differential abundance in log2 scale) and p-value adjusted for multiple hypothesis testing. $ES$ effect size. Proteins are ordered by p-value.

| *UniProt* | *Gene* | $coef$ | $p_{adj}$ | $ES$ |
|---|---|---|---|---|
| P80108 | GPLD1 | 0.452 | 8.27e-09 | 1.05 |

**Table S26**: Proteins significantly associated with ATC level 3 medication *ADRENERGICS, INHALANTS* (ATC3 R03A). $coef$ and $p_{adj}$: coefficient (representing the differential abundance in log2 scale) and p-value adjusted for multiple hypothesis testing. $ES$ effect size. Proteins are ordered by p-value.

| *UniProt* | *Gene* | $coef$ | $p_{adj}$ | $ES$ |
|---|---|---|---|---|
| P00488 | F13A1 | -0.483 | 0.0186 | -0.719 |

**Table S27**: Proteins significantly associated with ATC level 3 medication *CALCIUM* (ATC3 A12A). $coef$ and $p_{adj}$: coefficient (representing the differential abundance in log2 scale) and p-value adjusted for multiple hypothesis testing. $ES$ effect size. Proteins are ordered by p-value.

| *UniProt* | *Gene* | $coef$ | $p_{adj}$ | $ES$ |
|---|---|---|---|---|
| P01859 | IGHG2 | -0.253 | 0.00388 | -0.536 |
| P06727 | APOA4 | 0.185 | 0.0469 | 0.43 |

**Table S28**: Proteins significantly associated with ATC level 3 medication *LIPID MODIFYING AGENTS, PLAIN* (ATC3 C10A). $coef$ and $p_{adj}$: coefficient (representing the differential abundance in log2 scale) and p-value adjusted for multiple hypothesis testing. $ES$ effect size. Proteins are ordered by p-value.

| *UniProt* | *Gene* | $coef$ | $p_{adj}$ | $ES$ |
|---|---|---|---|---|
| P04114 | APOB | -0.264 | 1.36e-12 | -0.649 |

**Table S29**: Proteins significantly associated with ATC level 3 medication *ANGIOTENSIN II ANTAGONISTS, COMBINATIONS* (ATC3 C09D). $coef$ and $p_{adj}$: coefficient (representing the differential abundance in log2 scale) and p-value adjusted for multiple hypothesis testing. $ES$ effect size. Proteins are ordered by p-value.

| *UniProt* | *Gene* | $coef$ | $p_{adj}$ | $ES$ |
|---|---|---|---|---|
| P01024 | C3 | 0.0692 | 0.0322 | 0.346 |

**Table S30**: Proteins significantly associated with ATC level 3 medication *ANGIOTENSIN II ANTAGONISTS, PLAIN* (ATC3 C09C). $coef$ and $p_{adj}$: coefficient (representing the differential abundance in log2 scale) and p-value adjusted for multiple hypothesis testing. $ES$ effect size. Proteins are ordered by p-value.

| *UniProt* | *Gene* | $coef$ | $p_{adj}$ | $ES$ |
|---|---|---|---|---|
| P01602 | IGKV1-5 | 0.237 | 0.0284 | 0.441 |

#### Plasma proteins associated with medication (ATC level 4)

Associations were evaluated for 27 ATC level 4 medications taken on a regular basis (at least twice per week) by more than 14 study participants. The considered medications, numbers of participants and significant proteins are shown in the table below.

**Table S31**: Overview of association results for ATC level 4 medications in the CHRIS study subset. Only medications taken on a regular basis by more than 14 of the in total 3,632 study participants were considered. Columns *Participants* and *Proteins* list the number of participants taking the medication and number of significantly associated proteins.

| ATC | Name | Participants | Proteins |
|---|---|---|---|
| H03AA | Thyroid hormones | 262 | 0 |
| G03AA, G03FA | Progestogens and estrogens, fixed combinations | 260 | 52 |
| B01AC | Platelet aggregation inhibitors excl. heparin | 211 | 1 |
| C10AA | HMG CoA reductase inhibitors | 193 | 1 |
| C09AA | ACE inhibitors, plain | 156 | 0 |
| C07AB | Beta blocking agents, selective | 135 | 0 |
| C09DA | Angiotensin II antagonists and diuretics | 109 | 0 |
| N06AB | Selective serotonin reuptake inhibitors | 94 | 0 |
| A02BC | Proton pump inhibitors | 92 | 0 |
| C08CA | Dihydropyridine derivatives | 91 | 0 |
| C09CA | Angiotensin II antagonists, plain | 74 | 1 |
| A12AX | Calcium, combinations with vitamin D and/or other drugs | 62 | 1 |
| C09BA | ACE inhibitors and diuretics | 59 | 0 |
| N05BA, N05CD | Benzodiazepine derivatives | 51 | 0 |
| G04CA, C02CA | Alpha-adrenoreceptor antagonists | 48 | 5 |
| A10BA | Biguanides | 43 | 4 |
| N06AX | Other antidepressants | 41 | 0 |
| B01AA | Vitamin K antagonists | 37 | 14 |
| M04AA | Preparations inhibiting uric acid production | 36 | 0 |
| G03AB, G03FB | Progestogens and estrogens, sequential preparations | 34 | 36 |
| R03AK | Adrenergics in combination with corticosteroids or other drugs, excl. anticholinergics | 27 | 1 |
| G02BA | Intrauterine contraceptives | 25 | 0 |
| M01AE | Propionic acid derivatives | 24 | 0 |
| S01ED | Beta blocking agents | 21 | 0 |
| G02BB | Intravaginal contraceptives | 21 | 35 |
| N03AX | Other antiepileptics | 21 | 0 |
| H02AB, R03BA | Glucocorticoids | 18 | 2 |
| M01AB | Acetic acid derivatives and related substances | 16 | 0 |
| G03HB | Antiandrogens and estrogens | 16 | 38 |

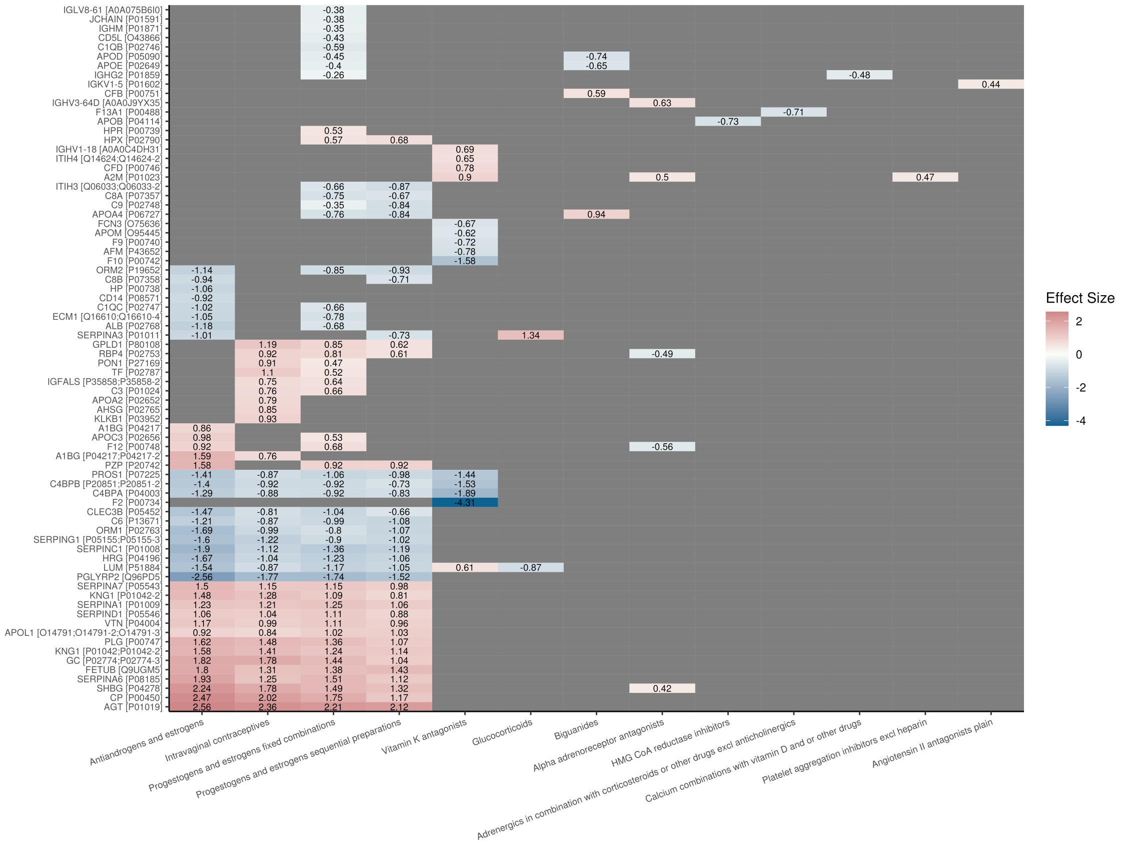

**Figure S156**: Significant associations between proteins (rows) and ATC level 4 medications (columns). Effect sizes (as a number and color coded) are only shown for significant associations.

**Table S32**: Proteins significantly associated with ATC level 4 medication *Progestogens and estrogens, fixed combinations* (ATC4 G03AA, G03FA). $coef$ and $p_{adj}$: coefficient (representing the differential abundance in log2 scale) and p-value adjusted for multiple hypothesis testing. $ES$ effect size. Proteins are ordered by p-value.

| $UniProt$ | $Gene$ | $coef$ | $p_{adj}$ | $ES$ |
|---|---|---|---|---|
| P01019 | AGT | 1.46 | 0 | 2.21 |
| P00450 | CP | 0.604 | 5.58e-212 | 1.75 |
| Q96PD5 | PGLYRP2 | -0.658 | 4.86e-174 | -1.74 |
| P04278 | SHBG | 1.84 | 4.15e-149 | 1.49 |
| P08185 | SERPINA6 | 0.86 | 6.93e-135 | 1.51 |
| P02774;P02774-3 | GC | 0.314 | 6.19e-127 | 1.44 |
| Q9UGM5 | FETUB | 1.2 | 1.16e-104 | 1.38 |
| P01008 | SERPINC1 | -0.273 | 4.67e-101 | -1.36 |
| P00747 | PLG | 0.304 | 7.55e-97 | 1.36 |
| P01009 | SERPINA1 | 0.386 | 2.42e-82 | 1.25 |
| P04196 | HRG | -0.479 | 4.06e-76 | -1.23 |
| P01042;P01042-2 | KNG1 | 0.257 | 3.53e-75 | 1.24 |
| P51884 | LUM | -0.456 | 3.69e-74 | -1.17 |
| P04004 | VTN | 0.285 | 1.71e-73 | 1.11 |
| P05543 | SERPINA7 | 0.592 | 4.32e-70 | 1.15 |
| P07225 | PROS1 | -0.326 | 1.93e-58 | -1.06 |

| *UniProt* | *Gene* | *coef* | $p_{adj}$ | *ES* |
|---|---|---|---|---|
| P05546 | SERPIND1 | 0.395 | 5.03e-58 | 1.11 |
| P01042-2 | KNG1 | 0.441 | 1.25e-56 | 1.09 |
| O14791;O14791-2;O14791-3 | APOL1 | 0.471 | 1.34e-52 | 1.02 |
| P05452 | CLEC3B | -0.333 | 7.97e-52 | -1.04 |
| P13671 | C6 | -0.321 | 1.11e-47 | -0.993 |
| P04003 | C4BPA | -0.279 | 7.19e-47 | -0.918 |
| P20742 | PZP | 0.771 | 5.44e-44 | 0.917 |
| P20851;P20851-2 | C4BPB | -0.445 | 1.64e-42 | -0.92 |
| P05155;P05155-3 | SERPING1 | -0.834 | 5.03e-38 | -0.901 |
| P80108 | GPLD1 | 0.367 | 7.78e-36 | 0.851 |
| P02753 | RBP4 | 0.274 | 3.2e-35 | 0.813 |
| P19652 | ORM2 | -0.369 | 1.77e-33 | -0.849 |
| P02763 | ORM1 | -0.342 | 4.68e-31 | -0.801 |
| P06727 | APOA4 | -0.328 | 4.31e-28 | -0.76 |
| Q16610;Q16610-4 | ECM1 | -0.386 | 7.84e-27 | -0.777 |
| P07357 | C8A | -0.213 | 1.19e-24 | -0.751 |
| P35858;P35858-2 | IGFALS | 0.284 | 6.38e-24 | 0.641 |
| P01024 | C3 | 0.132 | 4.03e-23 | 0.661 |
| Q06033;Q06033-2 | ITIH3 | -0.38 | 4.24e-22 | -0.662 |
| P02768 | ALB | -0.137 | 5.28e-22 | -0.68 |
| P00748 | F12 | 0.317 | 1.44e-20 | 0.676 |
| P02747 | C1QC | -0.212 | 1.7e-19 | -0.665 |
| P02746 | C1QB | -0.196 | 3.81e-15 | -0.594 |
| P02790 | HPX | 0.115 | 1.08e-14 | 0.568 |
| P02787 | TF | 0.13 | 1.57e-13 | 0.516 |
| P00739 | HPR | 0.32 | 5.35e-12 | 0.529 |
| P02656 | APOC3 | 0.229 | 5.58e-12 | 0.527 |
| P05090 | APOD | -0.18 | 6.41e-10 | -0.445 |
| P27169 | PON1 | 0.204 | 1.31e-09 | 0.467 |
| O43866 | CD5L | -0.32 | 4.16e-08 | -0.427 |
| P02649 | APOE | -0.194 | 1.25e-06 | -0.403 |
| A0A075B6I0 | IGLV8-61 | -0.299 | 8.51e-06 | -0.381 |
| P01591 | JCHAIN | -0.233 | 8.91e-06 | -0.376 |
| P02748 | C9 | -0.139 | 1.72e-05 | -0.353 |
| P01871 | IGHM | -0.264 | 3.54e-05 | -0.347 |
| P01859 | IGHG2 | -0.124 | 0.0273 | -0.263 |

**Table S33**: Proteins significantly associated with ATC level 4 medication *Antiandrogens and estrogens* (ATC4 G03HB). *coef* and $p_{adj}$: coefficient (representing the differential abundance in log2 scale) and p-value adjusted for multiple hypothesis testing. *ES* effect size. Proteins are ordered by p-value.

| *UniProt* | *Gene* | *coef* | $p_{adj}$ | *ES* |
|---|---|---|---|---|
| P01019 | AGT | 1.69 | 7.78e-46 | 2.56 |
| P00450 | CP | 0.855 | 1.26e-37 | 2.47 |
| Q96PD5 | PGLYRP2 | -0.968 | 1.31e-32 | -2.56 |
| P04278 | SHBG | 2.76 | 1.47e-28 | 2.24 |
| P08185 | SERPINA6 | 1.1 | 1.91e-18 | 1.93 |
| P02774;P02774-3 | GC | 0.396 | 6.8e-17 | 1.82 |
| P01008 | SERPINC1 | -0.38 | 4.52e-16 | -1.9 |
| Q9UGM5 | FETUB | 1.56 | 1.66e-14 | 1.8 |
| P00747 | PLG | 0.364 | 4.01e-11 | 1.62 |

| $UniProt$ | $Gene$ | $coef$ | $p_{adj}$ | $ES$ |
|---|---|---|---|---|
| P04196 | HRG | -0.647 | 4.9e-11 | -1.67 |
| P02763 | ORM1 | -0.72 | 1.18e-10 | -1.69 |
| P51884 | LUM | -0.602 | 3.31e-10 | -1.54 |
| P20742 | PZP | 1.33 | 3.94e-10 | 1.58 |
| P01042;P01042-2 | KNG1 | 0.329 | 1.11e-09 | 1.58 |
| P05543 | SERPINA7 | 0.769 | 2.9e-09 | 1.5 |
| P05155;P05155-3 | SERPING1 | -1.48 | 2.96e-09 | -1.6 |
| P04217;P04217-2 | A1BG | 0.417 | 5.9e-09 | 1.59 |
| P01042-2 | KNG1 | 0.599 | 4.97e-08 | 1.48 |
| P07225 | PROS1 | -0.433 | 6.2e-08 | -1.41 |
| P05452 | CLEC3B | -0.47 | 7.16e-08 | -1.47 |
| P20851;P20851-2 | C4BPB | -0.676 | 1.98e-07 | -1.4 |
| P04003 | C4BPA | -0.393 | 4.83e-07 | -1.29 |
| P04004 | VTN | 0.3 | 3.32e-06 | 1.17 |
| P01009 | SERPINA1 | 0.382 | 3.81e-06 | 1.23 |
| P13671 | C6 | -0.39 | 3.91e-05 | -1.21 |
| P02768 | ALB | -0.237 | 1e-04 | -1.18 |
| P19652 | ORM2 | -0.495 | 0.000299 | -1.14 |
| P05546 | SERPIND1 | 0.375 | 0.00127 | 1.06 |
| P00738 | HP | -0.675 | 0.00198 | -1.06 |
| Q16610;Q16610-4 | ECM1 | -0.521 | 0.00276 | -1.05 |
| P01011 | SERPINA3 | -0.282 | 0.00505 | -1.01 |
| P02747 | C1QC | -0.325 | 0.00519 | -1.02 |
| O14791;O14791-2;O14791-3 | APOL1 | 0.425 | 0.00856 | 0.922 |
| P02656 | APOC3 | 0.427 | 0.00995 | 0.984 |
| P00748 | F12 | 0.434 | 0.021 | 0.925 |
| P07358 | C8B | -0.337 | 0.0216 | -0.943 |
| P08571 | CD14 | -0.552 | 0.0311 | -0.921 |
| P04217 | A1BG | 0.197 | 0.0475 | 0.861 |

**Table S34**: Proteins significantly associated with ATC level 4 medication *Progestogens and estrogens, sequential preparations* (ATC4 G03AB, G03FB). $coef$ and $p_{adj}$: coefficient (representing the differential abundance in log2 scale) and p-value adjusted for multiple hypothesis testing. $ES$ effect size. Proteins are ordered by p-value.

| $UniProt$ | $Gene$ | $coef$ | $p_{adj}$ | $ES$ |
|---|---|---|---|---|
| P01019 | AGT | 1.4 | 8.3e-65 | 2.12 |
| Q96PD5 | PGLYRP2 | -0.578 | 3.1e-24 | -1.52 |
| P04278 | SHBG | 1.63 | 8.28e-21 | 1.32 |
| Q9UGM5 | FETUB | 1.23 | 3.74e-19 | 1.43 |
| P00450 | CP | 0.405 | 1.29e-17 | 1.17 |
| P01008 | SERPINC1 | -0.239 | 3.58e-13 | -1.19 |
| P08185 | SERPINA6 | 0.638 | 7.15e-13 | 1.12 |
| P02774;P02774-3 | GC | 0.226 | 2.54e-11 | 1.04 |
| P01042;P01042-2 | KNG1 | 0.236 | 1.78e-10 | 1.14 |
| P00747 | PLG | 0.241 | 4.11e-10 | 1.07 |
| P01009 | SERPINA1 | 0.327 | 8.83e-10 | 1.06 |
| P51884 | LUM | -0.409 | 8.99e-10 | -1.05 |
| P04196 | HRG | -0.412 | 3.13e-09 | -1.06 |
| P13671 | C6 | -0.35 | 3.93e-09 | -1.08 |
| P04004 | VTN | 0.247 | 4.56e-09 | 0.965 |

| *UniProt* | *Gene* | *coef* | $p_{adj}$ | *ES* |
|---|---|---|---|---|
| P02763 | ORM1 | -0.455 | 8.83e-09 | -1.07 |
| O14791;O14791-2;O14791-3 | APOL1 | 0.475 | 1.3e-08 | 1.03 |
| P05543 | SERPINA7 | 0.504 | 3.11e-08 | 0.983 |
| P07225 | PROS1 | -0.301 | 5.71e-08 | -0.978 |
| P05155;P05155-3 | SERPING1 | -0.942 | 1.12e-07 | -1.02 |
| P20742 | PZP | 0.771 | 6.29e-07 | 0.918 |
| P19652 | ORM2 | -0.404 | 3.34e-06 | -0.928 |
| P04003 | C4BPA | -0.253 | 5.77e-06 | -0.832 |
| Q06033;Q06033-2 | ITIH3 | -0.5 | 6.69e-06 | -0.872 |
| P05546 | SERPIND1 | 0.313 | 1.22e-05 | 0.881 |
| P06727 | APOA4 | -0.362 | 3.39e-05 | -0.84 |
| P02748 | C9 | -0.33 | 5.02e-05 | -0.835 |
| P01042-2 | KNG1 | 0.326 | 0.000117 | 0.806 |
| P20851;P20851-2 | C4BPB | -0.352 | 0.000722 | -0.728 |
| P01011 | SERPINA3 | -0.205 | 0.00201 | -0.734 |
| P07358 | C8B | -0.253 | 0.00562 | -0.708 |
| P02790 | HPX | 0.138 | 0.00628 | 0.683 |
| P05452 | CLEC3B | -0.212 | 0.00702 | -0.663 |
| P02753 | RBP4 | 0.205 | 0.012 | 0.61 |
| P07357 | C8A | -0.189 | 0.0138 | -0.669 |
| P80108 | GPLD1 | 0.269 | 0.0154 | 0.623 |

**Table S35**: Proteins significantly associated with ATC level 4 medication *Intravaginal contraceptives* (ATC4 G02BB). *coef* and $p_{adj}$: coefficient (representing the differential abundance in log2 scale) and p-value adjusted for multiple hypothesis testing. *ES* effect size. Proteins are ordered by p-value.

| *UniProt* | *Gene* | *coef* | $p_{adj}$ | *ES* |
|---|---|---|---|---|
| P01019 | AGT | 1.56 | 1.9e-50 | 2.36 |
| P00450 | CP | 0.698 | 1.22e-32 | 2.02 |
| P04278 | SHBG | 2.2 | 1.61e-23 | 1.78 |
| P02774;P02774-3 | GC | 0.388 | 4.43e-21 | 1.78 |
| Q96PD5 | PGLYRP2 | -0.67 | 3.19e-20 | -1.77 |
| P00747 | PLG | 0.331 | 4.95e-12 | 1.48 |
| P01042;P01042-2 | KNG1 | 0.293 | 4.85e-10 | 1.41 |
| Q9UGM5 | FETUB | 1.13 | 7.51e-10 | 1.31 |
| P08185 | SERPINA6 | 0.711 | 7.69e-10 | 1.25 |
| P01009 | SERPINA1 | 0.376 | 6.31e-08 | 1.21 |
| P01042-2 | KNG1 | 0.519 | 8.48e-08 | 1.28 |
| P01008 | SERPINC1 | -0.225 | 6.09e-07 | -1.12 |
| P05543 | SERPINA7 | 0.591 | 6.14e-07 | 1.15 |
| P05155;P05155-3 | SERPING1 | -1.13 | 8.33e-07 | -1.22 |
| P80108 | GPLD1 | 0.512 | 8.72e-07 | 1.19 |
| P02787 | TF | 0.278 | 5.2e-06 | 1.1 |
| P04004 | VTN | 0.254 | 1.09e-05 | 0.989 |
| P04196 | HRG | -0.404 | 3.21e-05 | -1.04 |
| P05546 | SERPIND1 | 0.37 | 8.61e-05 | 1.04 |
| P02763 | ORM1 | -0.425 | 0.000207 | -0.995 |
| P02753 | RBP4 | 0.309 | 0.000439 | 0.917 |
| P04003 | C4BPA | -0.266 | 0.000717 | -0.876 |
| P20851;P20851-2 | C4BPB | -0.444 | 0.000796 | -0.918 |
| P51884 | LUM | -0.337 | 0.00104 | -0.866 |

| *UniProt* | *Gene* | *coef* | $p_{adj}$ | *ES* |
|---|---|---|---|---|
| P07225 | PROS1 | -0.267 | 0.00165 | -0.868 |
| P03952 | KLKB1 | 0.297 | 0.00219 | 0.93 |
| P27169 | PON1 | 0.395 | 0.00309 | 0.906 |
| P13671 | C6 | -0.281 | 0.00346 | -0.869 |
| O14791;O14791-2;O14791-3 | APOL1 | 0.386 | 0.00453 | 0.838 |
| P02765 | AHSG | 0.237 | 0.0058 | 0.851 |
| P35858;P35858-2 | IGFALS | 0.33 | 0.0104 | 0.746 |
| P05452 | CLEC3B | -0.259 | 0.0132 | -0.81 |
| P01024 | C3 | 0.151 | 0.0177 | 0.758 |
| P02652 | APOA2 | 0.172 | 0.0312 | 0.786 |
| P04217;P04217-2 | A1BG | 0.199 | 0.0456 | 0.759 |

**Table S36**: Proteins significantly associated with ATC level 4 medication *Vitamin K antagonists* (ATC4 B01AA). *coef* and $p_{adj}$: coefficient (representing the differential abundance in log2 scale) and p-value adjusted for multiple hypothesis testing. *ES* effect size. Proteins are ordered by p-value.

| *UniProt* | *Gene* | *coef* | $p_{adj}$ | *ES* |
|---|---|---|---|---|
| P00734 | F2 | -0.857 | 2.35e-164 | -4.31 |
| P04003 | C4BPA | -0.573 | 3.01e-33 | -1.89 |
| P20851;P20851-2 | C4BPB | -0.74 | 1.12e-19 | -1.53 |
| P00742 | F10 | -0.504 | 1.28e-19 | -1.58 |
| P07225 | PROS1 | -0.442 | 4e-18 | -1.44 |
| P01023 | A2M | 0.373 | 1.91e-06 | 0.897 |
| P43652 | AFM | -0.239 | 0.000378 | -0.776 |
| P00746 | CFD | 0.439 | 0.000418 | 0.781 |
| O75636 | FCN3 | -0.4 | 0.00366 | -0.669 |
| P00740 | F9 | -0.584 | 0.00379 | -0.724 |
| P51884 | LUM | 0.238 | 0.00796 | 0.61 |
| A0A0C4DH31 | IGHV1-18 | 0.555 | 0.00957 | 0.688 |
| Q14624;Q14624-2 | ITIH4 | 0.141 | 0.0158 | 0.652 |
| O95445 | APOM | -0.217 | 0.0289 | -0.616 |

**Table S37**: Proteins significantly associated with ATC level 4 medication *Alpha-adrenoreceptor antagonists* (ATC4 G04CA, C02CA). *coef* and $p_{adj}$: coefficient (representing the differential abundance in log2 scale) and p-value adjusted for multiple hypothesis testing. *ES* effect size. Proteins are ordered by p-value.

| *UniProt* | *Gene* | *coef* | $p_{adj}$ | *ES* |
|---|---|---|---|---|
| A0A0J9YX35 | IGHV3-64D | 0.335 | 0.00302 | 0.634 |
| P00748 | F12 | -0.264 | 0.0203 | -0.562 |
| P02753 | RBP4 | -0.166 | 0.0416 | -0.493 |
| P04278 | SHBG | 0.524 | 0.042 | 0.424 |
| P01023 | A2M | 0.209 | 0.0433 | 0.503 |

**Table S38**: Proteins significantly associated with ATC level 4 medication *Biguanides* (ATC4 A10BA). $coef$ and $p_{adj}$: coefficient (representing the differential abundance in log2 scale) and p-value adjusted for multiple hypothesis testing. $ES$ effect size. Proteins are ordered by p-value.

| $UniProt$ | $Gene$ | $coef$ | $p_{adj}$ | $ES$ |
|---|---|---|---|---|
| P06727 | APOA4 | 0.404 | 1.77e-07 | 0.938 |
| P05090 | APOD | -0.299 | 0.000112 | -0.741 |
| P02649 | APOE | -0.314 | 0.00918 | -0.653 |
| P00751 | CFB | 0.141 | 0.0349 | 0.586 |

**Table S39**: Proteins significantly associated with ATC level 4 medication *Glucocorticoids* (ATC4 H02AB, R03BA). $coef$ and $p_{adj}$: coefficient (representing the differential abundance in log2 scale) and p-value adjusted for multiple hypothesis testing. $ES$ effect size. Proteins are ordered by p-value.

| $UniProt$ | $Gene$ | $coef$ | $p_{adj}$ | $ES$ |
|---|---|---|---|---|
| P01011 | SERPINA3 | 0.374 | 1.67e-06 | 1.34 |
| P51884 | LUM | -0.341 | 0.00509 | -0.875 |

**Table S40**: Proteins significantly associated with ATC level 4 medication *Platelet aggregation inhibitors excl. heparin* (ATC4 B01AC). $coef$ and $p_{adj}$: coefficient (representing the differential abundance in log2 scale) and p-value adjusted for multiple hypothesis testing. $ES$ effect size. Proteins are ordered by p-value.

| $UniProt$ | $Gene$ | $coef$ | $p_{adj}$ | $ES$ |
|---|---|---|---|---|
| P01023 | A2M | 0.194 | 1.69e-06 | 0.467 |

**Table S41**: Proteins significantly associated with ATC level 4 medication *HMG CoA reductase inhibitors* (ATC4 C10AA). $coef$ and $p_{adj}$: coefficient (representing the differential abundance in log2 scale) and p-value adjusted for multiple hypothesis testing. $ES$ effect size. Proteins are ordered by p-value.

| $UniProt$ | $Gene$ | $coef$ | $p_{adj}$ | $ES$ |
|---|---|---|---|---|
| P04114 | APOB | -0.295 | 4.97e-15 | -0.726 |

**Table S42**: Proteins significantly associated with ATC level 4 medication *Angiotensin II antagonists, plain* (ATC4 C09CA). $coef$ and $p_{adj}$: coefficient (representing the differential abundance in log2 scale) and p-value adjusted for multiple hypothesis testing. $ES$ effect size. Proteins are ordered by p-value.

| $UniProt$ | $Gene$ | $coef$ | $p_{adj}$ | $ES$ |
|---|---|---|---|---|
| P01602 | IGKV1-5 | 0.237 | 0.028 | 0.441 |

**Table S43**: Proteins significantly associated with ATC level 4 medication *Calcium, combinations with vitamin D and/or other drugs* (ATC4 A12AX). *coef* and $p_{adj}$: coefficient (representing the differential abundance in log2 scale) and p-value adjusted for multiple hypothesis testing. *ES* effect size. Proteins are ordered by p-value.

| *UniProt* | *Gene* | *coef* | $p_{adj}$ | *ES* |
|---|---|---|---|---|
| P01859 | IGHG2 | -0.224 | 0.0443 | -0.476 |

**Table S44**: Proteins significantly associated with ATC level 4 medication *Adrenergics in combination with corticosteroids or other drugs, excl. anticholinergics* (ATC4 R03AK). *coef* and $p_{adj}$: coefficient (representing the differential abundance in log2 scale) and p-value adjusted for multiple hypothesis testing. *ES* effect size. Proteins are ordered by p-value.

| *UniProt* | *Gene* | *coef* | $p_{adj}$ | *ES* |
|---|---|---|---|---|
| P00488 | F13A1 | -0.474 | 0.0412 | -0.707 |

**Table S45** (external spreadsheet, xlsx format): Comparison with results from literature.

### Extended Material and Methods

**Table S46**: Set up for gradient separation (left) and wash and equilibration (right).

| Gradient pump | | | | Regeneration pump | | | |
|---|---|---|---|---|---|---|---|
| Time | %A | %B | FR [ml/mn] | Time | %A | %B | FR [ml/mn] |
| 0 | 97 | 3 | 0.8 | 0 | 97 | 3 | 0.8 |
| 0.7 | 20 | 80 | 1 | 5 | 65 | 35 | 0.8 |
| 1.5 | 20 | 80 | 1 | 5.05 | 97 | 3 | 1 |
| 2 | 97 | 3 | 1 | 5.75 | 97 | 3 | 1 |
| 5 | 97 | 3 | 1 | 5.8 | 97 | 3 | 0.8 |
| 5.2 | 97 | 3 | 0.8 | | | | |
